# Global projections of lives saved from COVID-19 with universal mask use

**DOI:** 10.1101/2020.10.08.20209510

**Affiliations:** Institute for Health Metrics and Evaluation, University of Washington, Seattle, WA, United States; Department of Health Metrics Sciences, School of Medicine, University of Washington, Seattle, WA, United States; Department of Global Health, University of Washington, Seattle, WA, United States; Department of Political Science, University of Washington, Seattle, WA, United States; Center for Statistics and the Social Sciences, University of Washington, Seattle, WA, United States; Department of Applied Mathematics, University of Washington, Seattle, WA, United States; Big Data Institute, University of Oxford, Oxford, United Kingdom; Department of Environmental and Occupational Health Sciences, University of Washington, Seattle, WA, United States; Henry M Jackson School of International Studies, University of Washington, Seattle, WA, United States; Division of Cardiology, University of Washington, Seattle, WA, United States; Department of Epidemiology, Johns Hopkins University, Baltimore, MD, United States; School of Population and Public Health, University of British Columbia, Vancouver, BC, Canada

## Abstract

**BACKGROUND:** Social distancing mandates (SDM) have reduced health impacts from COVID-19 but also resulted in economic downturns that have led many nations to relax SDM. Until deployment of an efficacious and equitable vaccine, intervention options to reduce COVID-19 mortality and minimize restrictive SDM are sought by society.

**METHODS:** A susceptible-exposed-infectious-recovered (SEIR) deterministic transmission model was parameterized with data on reported deaths, cases, and select covariates to predict infections and deaths from COVID-19 through March 01, 2021. We explore three scenarios: a “non-adaptive” scenario where neither mask use or SDM adapt to changing conditions, a “reference” where current national levels of mask use are maintained and SDM reintroduced when deaths rise, and an increase in mask use to 95% coverage levels (“universal mask”). We reviewed published studies to set priors on the magnitude of reduction in transmission through increasing mask use.

**RESULTS:** Mask use was estimated at 59.0% of people globally on October 19, 2020. Universal mask use could avert 733,310 deaths (95% UI 385,981 to 1,107,759) between October 27, 2020 and March 01, 2021, the difference between the predicted 2.95 million deaths (95% UI 2.70 to 3.35) in the reference scenario and 2.22 million deaths (95% UI 2.00 to 2.45) in the universal mask scenario over this time period.

**CONCLUSIONS:** The cumulative toll of the COVID-19 pandemic could be substantially reduced by the universal adoption of masks before the availability of a vaccine. This low-cost, low-barrier policy, whether customary or mandated, has enormous health benefits with presumed marginal economic costs.

---

COVID-19 has spread to all regions of the world and as of October 25, 2020 nearly 43 million cases and 1,1953,034 deaths have been reported globally.^1^ In March 2020, nearly all nations put in place various social distancing mandates (SDM) (Figure 1) that have contributed to reducing the effects of COVID-19.^2–5^ The economic downturns associated with these measures subsequently led most nations to progressively relax SDM. As COVID-19 transmission and deaths rise globally and rebound in many countries,^1^ policymakers are keen to identify policy options to reduce COVID-19 mortality without re-imposing strict SDM before the widespread implantation of a vaccine.

**Figure 1.**
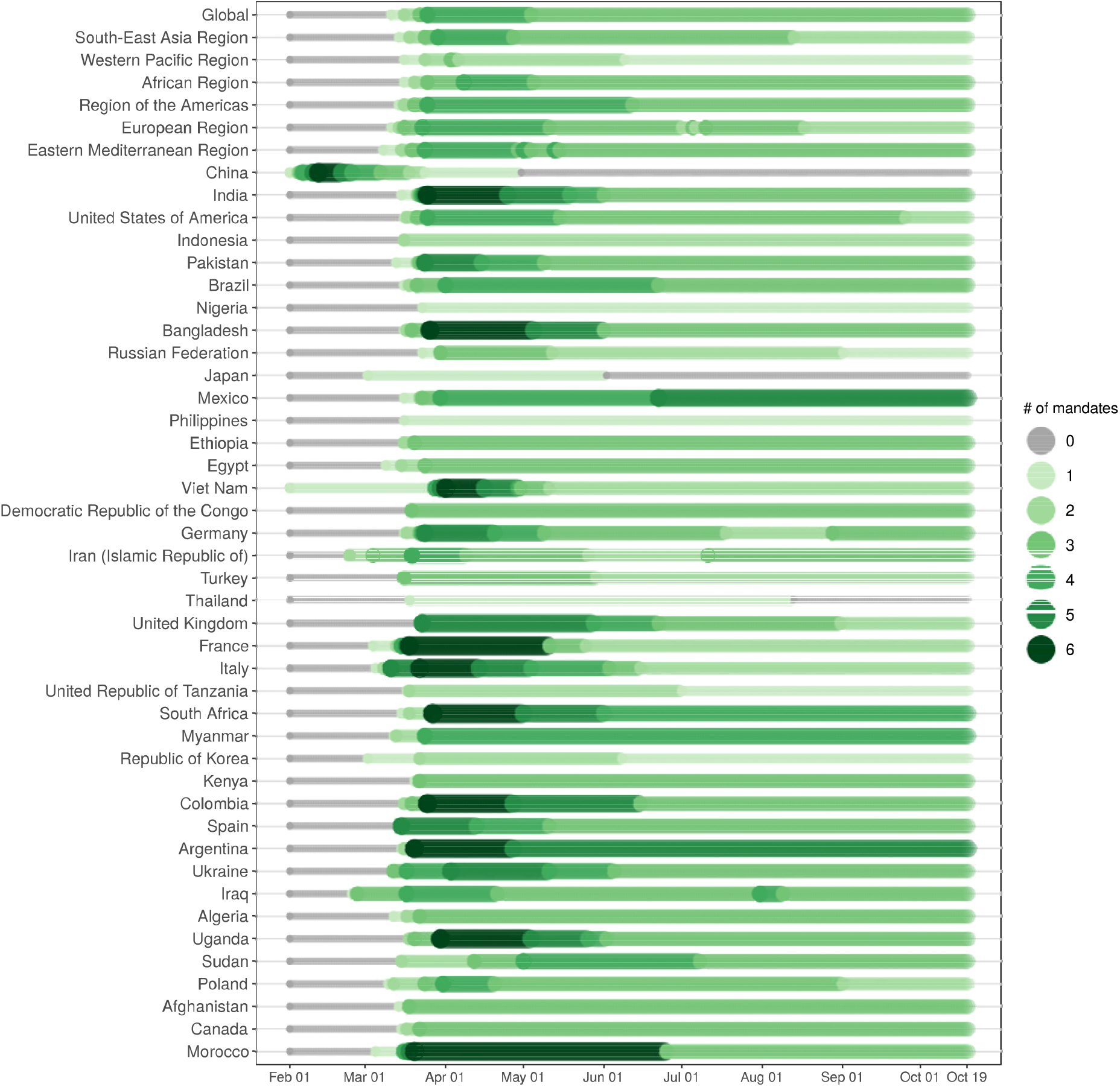
Number of social distancing mandates by WHO region and selected countries on a timeline starting February 01, 2020 through October 19, 2020.

Despite welcome news on vaccine development and efficacy of some candidates,^6,7^ safety, regulatory, manufacturing and distribution hurdles all need to be overcome before there will be an impact at scale.^8^ One existing and attractive strategy is the wearing of masks in public spaces or when physical distancing may not be possible. Several systematic reviews have suggested that cloth and other non-medical masks worn by the general public can reduce transmission.^9,10^ Moreover, simple non-medical masks are affordable and equitably accessible. Consequently, a growing number of national and local governments have recommended mask use, with more than 150 countries now mandating mask use.^11^ In nations such as the United States (US) and Brazil, mask use has become politicized, with widely varying views on the ethics, legality, and enforcement of mask use and mask use mandates.^12,13^ Evidence of the individual benefits and population health impacts of increased mask use is still extremely pertinent to these debates.

Masks reduce the transmission of respiratory pathogens *via* a physical barrier; the force of exhalation, coughing, or sneezing leads to impaction and interception of viral droplets (> 5µm in diameter) and aerosols (≤ 5 µm in diameter) onto the fibers of the mask.^14–16^ Given greater impaction efficiency for droplets, masks are most effective in blocking outward exhalation,^17^ although evidence from studies of air pollution aerosol indicates some protection of non-medical masks in blocking a small fraction of aerosol inhalation.^18^ Thus mask wearing is effective in intercepting particle emissions from the infected and reducing large particle transmission to the uninfected.^19^

In this paper, we use online survey data to assess current levels and trends in use of masks at the national level for 194 countries and quantify the benefits of universal mask use (here defined as 95% of individuals always wearing a mask when outside their home) on COVID-19 mortality in these countries using a COVID-19 transmission dynamics model. We then provide global and national estimates of lives saved through universal mask use between October 27, 2020 and March 01, 2021.

## Methods

Our analysis is comprised of two main components: an analysis of survey data on the levels and trends in mask use; and a deterministic transmission dynamics model with categories for susceptible, exposed, infected and recovered (SEIR) model of COVID-19 incidence and death. We explore three scenarios in the SEIR model: 1) a “non-adaptive” where mask use and SDM remain constant 2) a plausible “reference scenario” where SDMs are reinstated at a threshold level of daily deaths and current levels of mask use maintained and 3) a “universal mask” scenario based on the adoption of universal mask use at 95% coverage.

We used four main sources of data on self-reported use of masks to estimate past and current mask use globally, and make forecasts for future use: Facebook Global Symptom Survey,^20^ Facebook US Symptom Survey^21^, PREMISE surveys,^22^ and YouGov COVID-19 Behaviour Tracker surveys.^23^ Pooling estimates on the prevalence of self-reported mask use from these survey series provides a time series on trends in mask use in almost all countries. Between April 23, 2020 and October 19, 2020, Facebook has surveyed 30.86 million users from 198 countries and territories using an instrument with multiple items on behaviors related to COVID-19, including mask use. The YouGov surveys cover 29 countries and represent interviews with approximately 726,500 individuals since March 01, 2020 up until October 24, 2020. For the US, we used data collected through PREMISE and Facebook to inform the trend in mask use. There were 273,424 total PREMISE responses representing all 50 states and the District of Columbia with responses collected between April 21 and October 26. Facebook symptom surveys in the US started asking about mask use in the beginning of September (data range from September 8 to October 20) and included 1,813,000 respondents. Levels of mask use as reported by PREMISE are on average 30% lower across the USA compared to data from Facebook and YouGov, but other data sources do not provide a time trend in mask use. Therefore, we use the time trend implied by the PREMISE data and apply it to the level of mask use reported in the Facebook data. Details on the specific questions and responses used from each survey are identified in the Supplementary Appendix.

Mask use for each location was estimated using a spline-based smoothing process. For locations without data on mask use, we used national level estimates or regional estimates (see Supplementary Appendix for additional details).

### COVID-19 SEIR model construction

The IHME COVID-19 prediction model has been described in detail elsewhere.^24^ Briefly, we construct an SEIR model for each location we model; the Supplementary Appendix shows the basic health states included in the model and specific transition parameters. The critical driver of the epidemic is the rate at which susceptible individuals become infected in each location which is represented as:

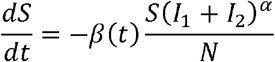

Where β_t_ is the transmission parameter for time period *t, α* represents a mixing coefficient to account for imperfect mixing within each location, S is the fraction of each location’s population that is susceptible, and I_1_ and I_2_ is the fraction that are infectious. Effective R_t_, the number of new infections caused by each case is a simple monotonic transformation of β_t_ and the fraction of the population that is susceptible. We use an efficient algorithm to directly estimate β_t_ in the past – see Supplementary Appendix for details.

The following covariates are included in the model: population density measured as the share of the population living in areas with more than 2,500 individuals per square kilometer, the fraction of the population living below 100 meters above sea level, smoking prevalence, particulate matter air pollution (PM2.5 population-weighted annual average concentration), mobility measured using cell phone apps, mask use, COVID-19 testing *per capita*, and pneumonia seasonality.^24^ To avoid over-estimating the effects of pneumonia seasonality and mask use we used constraints on each in the regression – see Supplementary Appendix for details. Specifically, for mask use, we did not let the regression estimate an effect size larger than what was consistent with the mask use meta-analysis of the individual level effect (Supplementary Appendix). To capture uncertainty in the input data, model parameters and regression coefficients linking β_jt_ to covariates, we generated 1,000 models for each location – see Supplementary Appendix for details.

We evaluated out of sample predictive validity for this modeling approach by holding out the last twelve weeks of data and compared predictions from the held-out data to what occurred; median absolute percent error for cumulative deaths at six weeks was 11% and twelve weeks was 24%.^25^ We also compared this model to other COVID-19 prediction models that make their estimates publicly available; overall, we find that our model is one of the best performing models at 12-weeks out-of-sample.^25^

### Scenario construction

We used the set of 1,000 SEIR models for each location to generate three scenarios: a plausible reference scenario, a universal mask use scenario, and a scenario wherein SDM continued to be lifted. In the plausible reference scenario key drivers such as mobility and testing *per capita* evolve according to past trends – see Supplementary Appendix for details. In the universal mask use scenario, we additionally assume that mask mandates and other campaigns lead to scale-up of mask use to 95% within 7 days of enactment; this represents the highest reported level of mask use (in Victoria, Australia Singapore) across the three survey platforms during the COVID-19 pandemic up to mid-October; other locations including Chile, Peru, and Spain have also approached this 95% coverage level from previous very low levels of coverage. The “universal mask” scenario assumes this level of mask use can be achieved through the adoption of mask use around the world. Both the reference and universal mask use scenarios additionally assume that SDM will be re-instated for 6 weeks if the daily death rate per location reaches 8 per million people per day.^26^ Finally, we estimate the trajectory of the epidemic in each location if SDM are not reinstated in response to changing pandemic conditions (“non-adaptive scenario”).

Our study follows the Guidelines for Accurate and Transparent Health Estimate Reporting. Data were not obtained from subjects for this study; we used pre-existing datasets identified through online searches, through outreach to institutions that hold relevant data such as ministries of health, or through individual collaborator reference and identification. This study was approved by University of Washington’s Human Subjects Division Study ID: STUDY00009060. Author contributions, including the identification of who analyzed data, are supplied in the Supplementary Appendix.

## Results

Figure 2 shows the relative risk of viral respiratory infection among users of different types of facemasks; additional details and results for this analysis are provided in the Supplementary Appendix. We found that non-medical masks among the general population reduced risk of viral respiratory infection by 40% (Relative risk 0.6, 95% CI 0.46 to 0.78).

**Figure 2.**
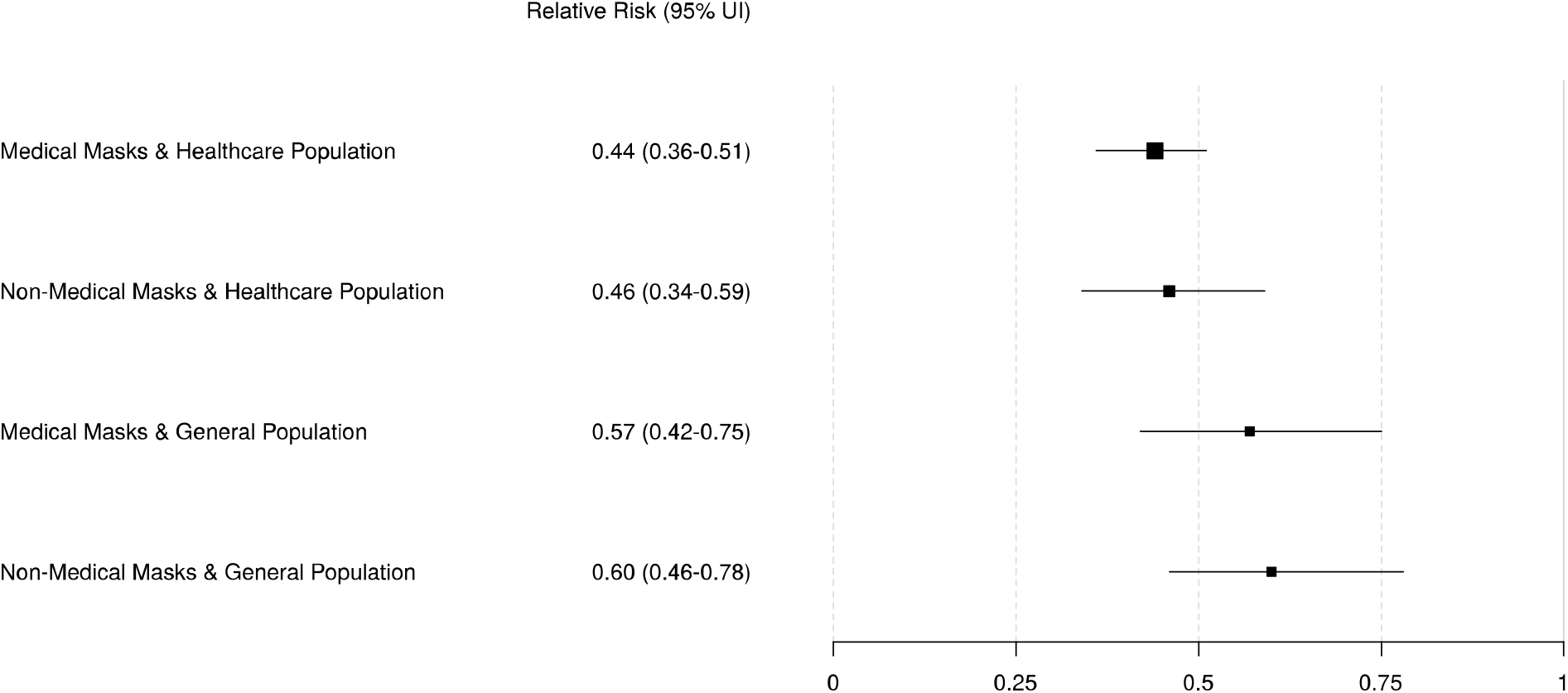
Relative risk of viral respiratory infection among users of different types of facemasks. This analysis includes only variables addressing facemask type (non-medical versus medical) and population type of mask user (general population versus healthcare setting). The size of the box is proportional to the precision of the estimate, based on number of observations, with more precise studies having larger boxes.

Based on survey data collected through smartphone hosted questionnaires, Figure 3 shows a map of mask use by location as of October 19, 2020, the last date of fully observed data in the model. Mask use is high in several locations that were at the forefront of the COVID-19 peak in the spring of 2020 including Italy, Spain, and most parts of Latin America (*i*.*e*. Chile, and Panama). The highest national mask use on October 19, 2020 was in Spain (93.9%), followed by Puerto Rico (92.6%), and Panama (92.5%). The lowest rates are seen in Northern Europe (Sweden, Norway, and Denmark < 4%) and East Africa (United Republic of Tanzania 2.1%). On October 19, women were more likely to report always wearing a mask (64% women, 58% men), with pronounced sex differences in sub-Saharan Africa, Central and Eastern Europe and Central Asia (See Supplementary Appendix). Mask use was highest among people who live in cities and towns (about 60%) and lowest in rural communities (less than 50%; see Supplementary Appendix). People over 65 years (56.4%) and young adults 18-24 (56.7%) were the least likely age groups to report always wearing a mask (other adult age groups were over 60% at the global level; see Supplementary Appendix).

**Figure 3.**
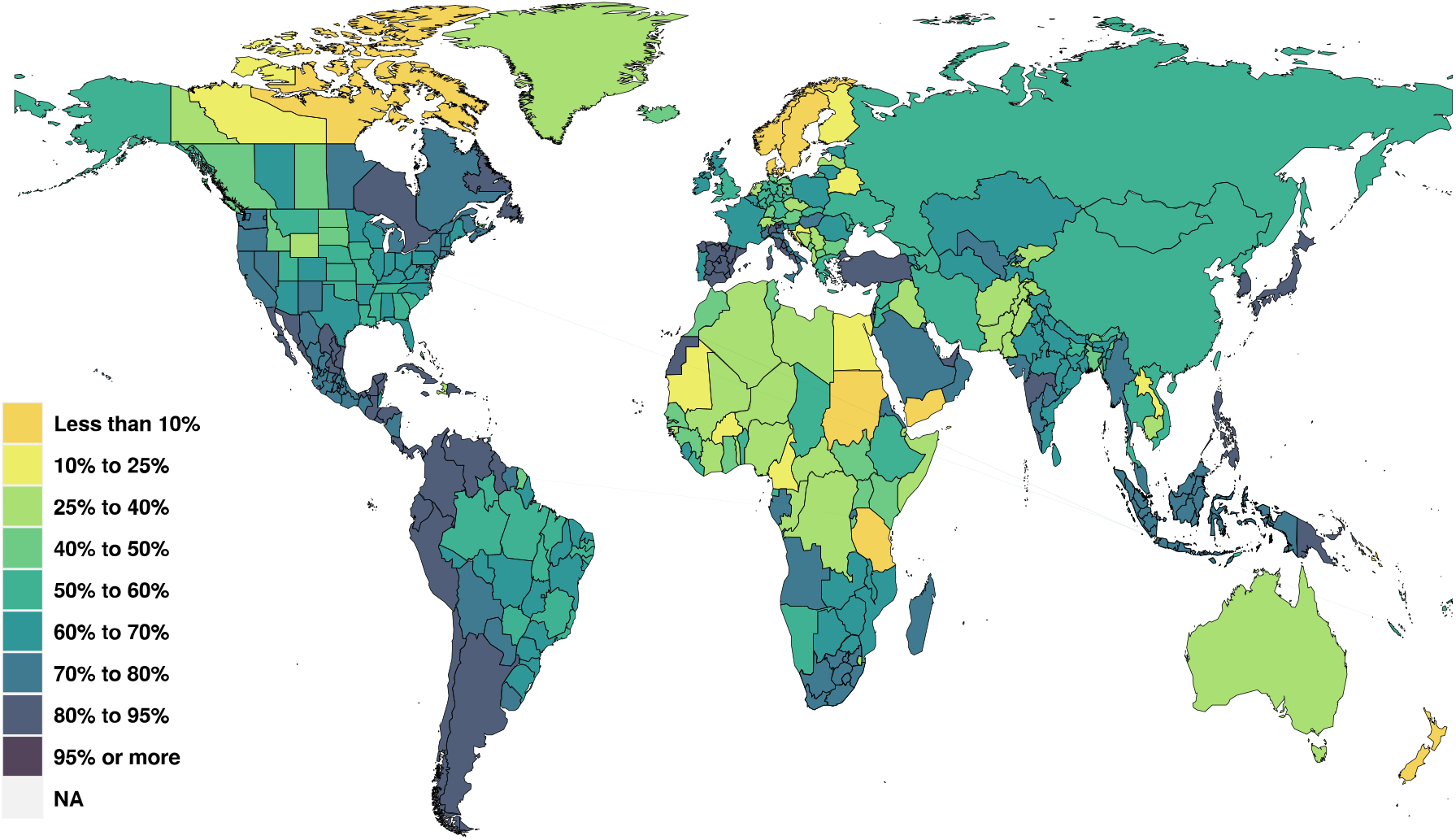
Proportion of the population that self-report always wearing a facemask when outside the home on October 19, 2020.

Figure 4 shows the global and WHO regional trends in mask use since the beginning of the epidemic. Mask use data start only in the beginning of April so the rapid expansion from likely a very low baseline pre-COVID-19 outside of East Asia most probably occurred in March. Mask use was estimated at 59.0% of people globally on October 19, 2020, ranging from 36.0% in the Eastern Mediterranean Region to 72.2% in the Region of the Americas (Figure 4). Mask use has increased in some countries where mandates have been put into place such as Belgium, Ireland and the Ethiopia (SI Figure 7). Mask use has declined in some settings in parallel with declining death rates such as Poland, Czechia, and Italy (SI Figure 7).

**Figure 4.**
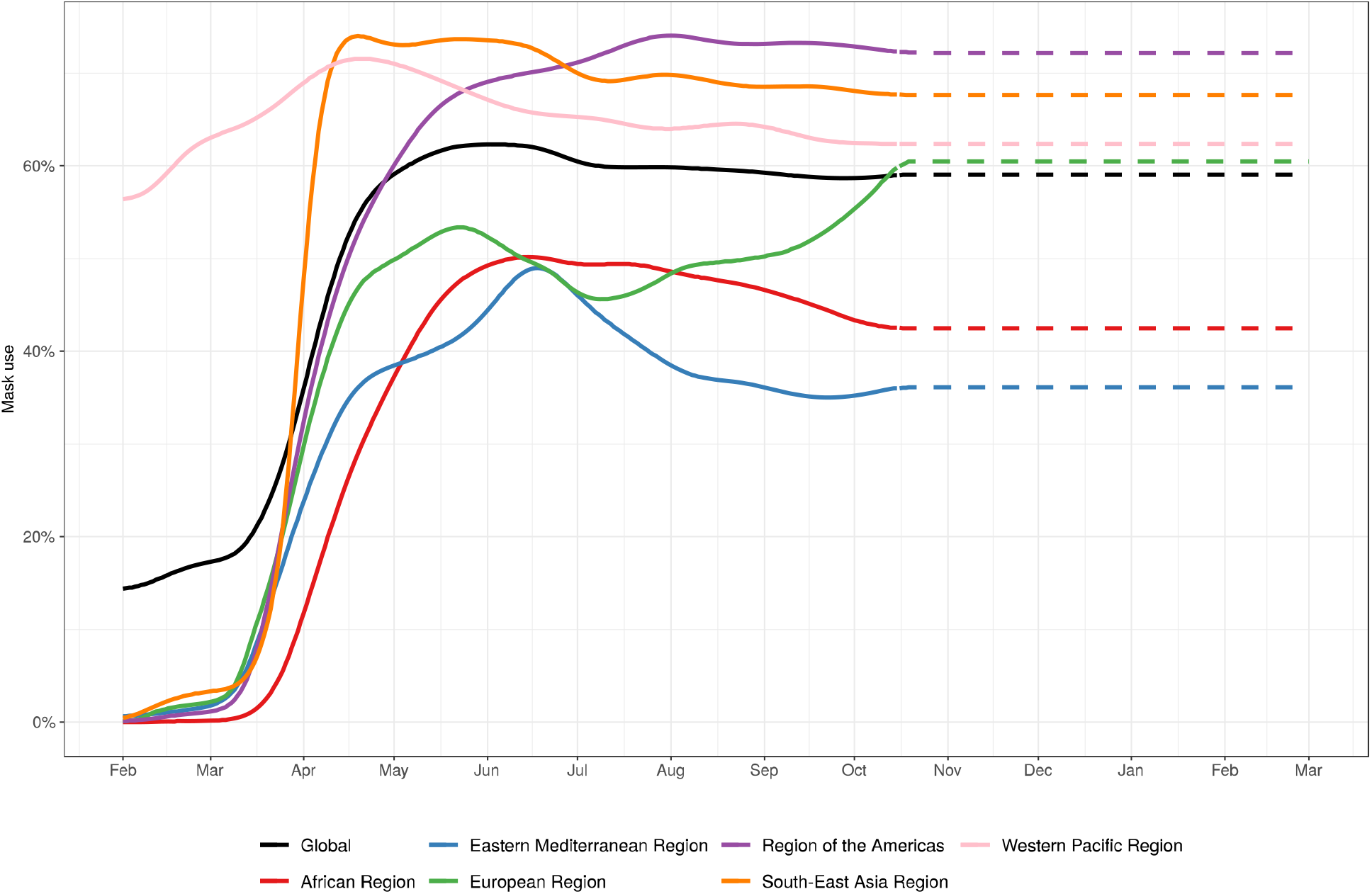
Proportion of the population that self-report always wearing a facemask when outside the home by WHO region between February 1, 2020 and March 01, 2021. Values after the last date of observed survey data are dashed lines and horizontal projections from the last observed values.

Global projections of COVID-19 deaths with and without adoption of universal mask use are shown in Figure 5. In our reference scenario, we expect daily COVID-19 deaths to increase in November through December leading to as many as 2.95 million deaths (95% UI 2.70 million to 3.35 million) by March 01, 2021. Uncertainty in daily deaths and cumulative deaths increases over time such that there is considerable uncertainty in global deaths by March 01, 2021. In contrast, the universal mask use scenario leads to a mean estimate of 2.22 million deaths (95% UI 2.00 million to 2.45 million) by March 01, 2021. The difference in mortality between the reference and universal mask use scenario suggests that 0.73 million lives (95% UI 0.39 million to 1.11 million) could be saved over this time period if 95% of people were to always wear masks when outside their home. Table 1 provides estimates by location of the expected deaths in the reference and universal mask use scenario along with the number of deaths saved through universal mask use. The countries where universal mask use would have the largest effect are populous nations where mask use is currently relatively low such as India (104,301 fewer deaths in universal mask scenario, 95% UI 60,679 to 151,873 deaths), the Germany (90,005 fewer deaths; 95% UI 27,524 to 190,010 deaths), and United States of America (82,150 fewer deaths; 95% UI 46,079 to 116,120 deaths). The greatest magnitude difference in mortality rate occurred in the Czechia (12,419 fewer deaths, 117 deaths per 100,000), Germany (90,005 fewer deaths, 106 deaths per 100,000) and Croatia (3,879 fewer deaths, 91 deaths per 100,000).

**Table 1.** The difference in cumulative deaths between the universal mask use scenario and the reference (current mask use) scenario globally and by country (and first administrative subnational units where applicable) within WHO regions, on March 01, 2021.

| Location | Lives saved (95% UI) | Percent difference in deaths (95% UI) | Cumulative deaths in reference scenario (95% UI) |
| --- | --- | --- | --- |
| Global | 733,310 (385,981 to 1,107,759) | -24.6% (-34.6 to -14.0%) | 2,954,098 (2,698,550 to 3,351,659) |
| African Region | 31,730 (14,284 to 58,110) | -41.7% (-58.2 to -24.5%) | 73,614 (54,785 to 101,214) |
| Algeria | 10,282 (4,196 to 20,655) | -73.3% (-87.5 to -54.7%) | 13,508 (7,358 to 24,041) |
| Angola | 335 (82 to 1,130) | -25.2% (-45.7 to -11.6%) | 1,171 (638 to 2,578) |
| Benin | 140 (0 to 1,119) | -35.2% (-89.8 to -0.7%) | 199 (42 to 1,274) |
| Botswana | 14 (0 to 97) | -15.0% (-59.0 to -0.8%) | 47 (23 to 196) |
| Burkina Faso | 236 (1 to 1,567) | -44.0% (-94.0 to -2.1%) | 321 (70 to 1,698) |
| Cabo Verde | 144 (66 to 237) | -37.0% (-52.7 to -21.0%) | 387 (248 to 488) |
| Cameroon | 33 (2 to 135) | -6.6% (-23.8 to -0.6%) | 463 (427 to 573) |
| Central African Republic | 40 (1 to 271) | -23.7% (-77.0 to -2.0%) | 107 (64 to 349) |
| Chad | 160 (7 to 791) | -38.9% (-80.3 to -6.8%) | 300 (109 to 1,047) |
| Comoros | 0 (0 to 0) | -0.8% (-1.1 to -0.5%) | 8 (8 to 8) |
| Congo | 154 (9 to 1,044) | -34.5% (-85.8 to -7.2%) | 284 (119 to 1,299) |
| Côte d'Ivoire | 436 (3 to 3,393) | -42.1% (-95.1 to -2.0%) | 577 (127 to 3,584) |
| Democratic Republic of the Congo | 443 (80 to 2,037) | -38.4% (-78.1 to -15.6%) | 918 (509 to 2,634) |
| Equatorial Guinea | 3 (1 to 8) | -3.4% (-9.1 to -1.2%) | 87 (85 to 93) |
| Eswatini | 44 (2 to 217) | -20.1% (-60.5 to -1.7%) | 174 (121 to 357) |
| Ethiopia | 3,077 (1,214 to 7,835) | -52.2% (-75.0 to -34.6%) | 5,587 (3,377 to 10,504) |
| Gabon | 5 (0 to 39) | -5.2% (-35.5 to -0.1%) | 63 (54 to 107) |
| Gambia | 98 (4 to 425) | -31.2% (-72.6 to -3.1%) | 236 (126 to 603) |
| Ghana | 307 (75 to 1,195) | -33.3% (-67.0 to -14.4%) | 810 (513 to 1,796) |
| Guinea | 408 (13 to 2,148) | -60.5% (-93.3 to -14.8%) | 519 (89 to 2,311) |
| Guinea-Bissau | 177 (3 to 823) | -52.9% (-91.2 to -6.2%) | 242 (49 to 911) |
| Kenya | 9,914 (3,128 to 19,964) | -68.5% (-86.7 to -45.8%) | 13,905 (5,902 to 24,512) |
| Lesotho | 33 (0 to 265) | -19.5% (-80.7 to -0.2%) | 81 (44 to 322) |
| Liberia | 2 (0 to 10) | -2.7% (-11.0 to -0.4%) | 85 (82 to 95) |
| Madagascar | 5 (1 to 17) | -1.9% (-6.1 to -0.5%) | 261 (249 to 290) |
| Malawi | 4 (1 to 14) | -2.1% (-6.8 to -0.5%) | 196 (188 to 212) |
| Mali | 66 (5 to 342) | -23.5% (-67.8 to -3.3%) | 210 (141 to 495) |
| Mauritania | 54 (5 to 213) | -20.9% (-55.9 to -3.2%) | 221 (170 to 383) |
| Mauritius | 0 (0 to 0) | -0.3% (-0.5 to -0.2%) | 11 (11 to 11) |
| Mozambique | 55 (8 to 286) | -19.8% (-49.9 to -6.6%) | 217 (124 to 589) |
| Namibia | 18 (2 to 64) | -9.6% (-26.6 to -1.3%) | 172 (142 to 245) |
| Niger | 80 (2 to 759) | -25.8% (-89.6 to -3.1%) | 154 (73 to 856) |
| Nigeria | 266 (33 to 958) | -16.0% (-44.0 to -2.8%) | 1,455 (1,191 to 2,207) |
| Rwanda | 43 (0 to 316) | -17.6% (-55.7 to -0.7%) | 126 (39 to 637) |
| Sao Tome and Principe | 1 (0 to 5) | -6.4% (-23.7 to -1.0%) | 17 (16 to 22) |
| Senegal | 150 (69 to 337) | -27.2% (-46.6 to -15.4%) | 532 (439 to 726) |
| Sierra Leone | 44 (1 to 297) | -21.0% (-71.7 to -1.3%) | 132 (76 to 413) |
| South Africa | 1,457 (180 to 3,935) | -5.5% (-12.8 to -0.9%) | 25,084 (20,625 to 32,461) |
| South Sudan | 234 (0 to 1,977) | -39.0% (-92.8 to -0.3%) | 316 (55 to 2,098) |
| Togo | 193 (31 to 799) | -57.8% (-88.1 to -29.9%) | 285 (101 to 921) |
| Uganda | 1,992 (75 to 10,752) | -75.4% (-97.0 to -38.4%) | 2,261 (193 to 11,088) |
| United Republic of Tanzania | 477 (178 to 1,360) | -39.5% (-65.9 to -22.0%) | 1,116 (780 to 2,062) |
| Zambia | 9 (1 to 40) | -2.3% (-9.5 to -0.2%) | 367 (344 to 434) |
| Zimbabwe | 94 (7 to 527) | -16.5% (-54.7 to -2.7%) | 402 (268 to 963) |
| Eastern Mediterranean Region | 85,430 (37,549 to 151,164) | -33.8% (-51.8 to -16.6%) | 248,219 (219,986 to 303,019) |
| Afghanistan | 2,864 (599 to 10,488) | -52.2% (-82.9 to -26.2%) | 4,735 (2,250 to 12,893) |
| Bahrain | 382 (132 to 628) | -36.9% (-52.6 to -21.6%) | 1,013 (575 to 1,271) |
| Djibouti | 128 (0 to 778) | -28.6% (-87.9 to -0.2%) | 215 (62 to 1,014) |
| Egypt | 8,687 (3,941 to 22,233) | -51.3% (-75.3 to -34.4%) | 16,057 (11,295 to 29,593) |
| Iran (Islamic Republic of) | 10,794 (0 to 25,342) | -12.7% (-29.9 to 0.0%) | 86,083 (81,032 to 91,487) |
| Iraq | 14,273 (9,139 to 20,470) | -47.1% (-59.0 to -33.2%) | 30,080 (25,712 to 35,048) |
| Jordan | 468 (0 to 3,241) | -6.1% (-42.7 to 0.0%) | 7,126 (6,072 to 8,383) |
| Kuwait | 822 (280 to 1,399) | -30.2% (-47.6 to -9.9%) | 2,719 (1,723 to 3,372) |
| Lebanon | 1,540 (61 to 2,425) | -45.6% (-69.5 to -1.5%) | 3,434 (1,841 to 4,080) |
| Libya | 2,618 (1,318 to 3,400) | -55.2% (-71.0 to -26.5%) | 4,765 (3,697 to 5,575) |
| Morocco | 7,351 (1,887 to 13,297) | -30.5% (-55.6 to -7.7%) | 24,183 (22,868 to 25,973) |
| Oman | 450 (276 to 715) | -18.4% (-26.7 to -11.5%) | 2,436 (2,074 to 2,840) |
| Pakistan | 23,141 (5,635 to 56,754) | -64.7% (-86.0 to -38.9%) | 32,756 (14,367 to 66,719) |
| Azad Jammu & Kashmir | 1,187 (388 to 1,951) | -55.1% (-87.9 to -17.8%) | 2,182 (1,795 to 2,753) |
| Balochistan | 959 (43 to 4,380) | -65.1% (-94.6 to -21.0%) | 1,181 (204 to 4,776) |
| Gilgit-Baltistan | 338 (0 to 852) | -51.4% (-87.0 to -0.5%) | 546 (91 to 1,186) |
| Islamabad Capital Territory | 440 (41 to 761) | -39.8% (-67.3 to -3.4%) | 1,117 (875 to 1,273) |
| Khyber Pakhtunkhwa | 1,384 (172 to 5,646) | -41.4% (-80.1 to -11.8%) | 2,752 (1,475 to 6,989) |
| Punjab | 17,106 (2,067 to 47,426) | -76.1% (-94.0 to -44.9%) | 20,339 (4,713 to 51,175) |
| Sindh | 1,726 (375 to 5,214) | -33.2% (-63.4 to -12.0%) | 4,639 (3,104 to 8,439) |
| Qatar | 428 (138 to 882) | -41.0% (-62.9 to -19.9%) | 1,022 (503 to 1,675) |
| Saudi Arabia | 2,001 (1,060 to 3,395) | -20.2% (-30.6 to -12.2%) | 9,759 (8,037 to 11,583) |
| Somalia | 227 (4 to 1,886) | -34.1% (-90.4 to -3.7%) | 357 (108 to 2,204) |
| Sudan | 1,671 (2 to 18,812) | -23.6% (-94.6 to -0.2%) | 2,539 (837 to 19,741) |
| Syrian Arab Republic | 2,634 (1,015 to 5,748) | -71.9% (-88.8 to -50.9%) | 3,536 (1,765 to 6,905) |
| Tunisia | 4,212 (229 to 8,396) | -35.9% (-67.3 to -2.4%) | 11,619 (7,962 to 17,998) |
| United Arab Emirates | 735 (115 to 1,737) | -22.9% (-41.8 to -7.5%) | 3,186 (1,007 to 5,329) |
| Yemen | 6 (1 to 16) | -1.0% (-2.6 to -0.2%) | 600 (593 to 613) |
| European Region | 355,754 (175,846 to 583,089) | -31.1% (-44.2 to -17.6%) | 1,126,561 (960,340 to 1,421,258) |
| Albania | 1,238 (701 to 1,666) | -53.3% (-68.3 to -28.8%) | 2,328 (2,074 to 2,719) |
| Andorra | 7 (1 to 22) | -5.9% (-15.6 to -1.5%) | 113 (91 to 150) |
| Armenia | 1,539 (834 to 2,548) | -29.0% (-43.1 to -17.0%) | 5,288 (4,120 to 7,104) |
| Austria | 4,442 (1,472 to 8,797) | -44.1% (-72.9 to -18.9%) | 10,005 (6,122 to 16,164) |
| Azerbaijan | 3,320 (698 to 7,080) | -32.9% (-54.4 to -9.3%) | 9,738 (6,631 to 15,271) |
| Belarus | 4,835 (3,902 to 5,739) | -76.8% (-82.3 to -66.9%) | 6,283 (5,552 to 7,070) |
| Belgium | 6,473 (1,641 to 12,748) | -23.4% (-38.0 to -8.0%) | 26,692 (19,781 to 34,565) |
| Bosnia and Herzegovina | 982 (140 to 1,883) | -28.8% (-48.0 to -4.8%) | 3,343 (2,669 to 4,733) |
| Bulgaria | 4,847 (1,108 to 10,913) | -43.6% (-68.2 to -14.8%) | 10,559 (7,051 to 16,914) |
| Croatia | 3,879 (1,001 to 8,887) | -55.1% (-76.8 to -24.0%) | 6,700 (3,853 to 12,411) |
| Cyprus | 134 (1 to 727) | -42.9% (-86.8 to -3.7%) | 203 (31 to 1,010) |
| Czechia | 12,419 (6,969 to 20,681) | -49.4% (-67.0 to -30.5%) | 25,242 (16,055 to 37,207) |
| Denmark | 3,975 (1,998 to 7,339) | -76.1% (-87.3 to -61.1%) | 5,142 (2,988 to 8,576) |
| Estonia | 138 (3 to 548) | -37.7% (-73.8 to -3.2%) | 279 (83 to 812) |
| Finland | 501 (108 to 2,032) | -47.6% (-81.3 to -22.4%) | 902 (478 to 2,592) |
| France | 19,695 (9,480 to 39,305) | -20.1% (-31.6 to -11.5%) | 95,640 (78,657 to 130,050) |
| Georgia | 900 (0 to 2,099) | -29.3% (-61.8 to 0.0%) | 2,991 (1,950 to 4,610) |
| Germany | 90,005 (27,524 to 190,010) | -60.6% (-79.6 to -38.4%) | 140,720 (69,488 to 257,049) |
| Baden-Württemberg | 21,001 (5,002 to 55,334) | -61.6% (-82.6 to -35.4%) | 31,229 (12,664 to 72,101) |
| Bavaria | 12,983 (3,551 to 34,630) | -52.2% (-76.4 to -26.2%) | 22,977 (11,657 to 47,058) |
| Berlin | 2,914 (424 to 8,629) | -70.7% (-89.8 to -46.8%) | 3,934 (798 to 10,620) |
| Brandenburg | 1,831 (32 to 8,711) | -60.4% (-86.7 to -13.2%) | 2,647 (249 to 11,638) |
| Bremen | 145 (6 to 607) | -45.2% (-82.7 to -8.0%) | 241 (75 to 765) |
| Hamburg | 851 (81 to 2,629) | -55.8% (-81.8 to -20.1%) | 1,340 (398 to 3,477) |
| Hesse | 17,888 (3,016 to 41,332) | -61.7% (-82.3 to -37.2%) | 27,695 (7,661 to 62,642) |
| Lower Saxony | 4,273 (1,359 to 9,749) | -69.0% (-86.0 to -48.1%) | 5,963 (2,632 to 11,982) |
| Mecklenburg-Vorpommern | 81 (2 to 665) | -41.2% (-90.6 to -7.6%) | 117 (28 to 709) |
| North Rhine-Westphalia | 20,416 (5,689 to 59,418) | -58.8% (-81.3 to -31.4%) | 32,632 (13,171 to 76,786) |
| Rhineland-Palatinate | 1,521 (362 to 4,208) | -68.1% (-87.1 to -44.0%) | 2,104 (796 to 5,067) |
| Saarland | 149 (0 to 1,136) | -19.1% (-72.4 to -0.3%) | 383 (179 to 1,673) |
| Saxony-Anhalt | 614 (54 to 2,109) | -64.3% (-89.2 to -32.6%) | 863 (165 to 2,819) |
| Saxony | 4,009 (1,168 to 12,366) | -59.8% (-83.4 to -33.6%) | 6,476 (2,043 to 17,251) |
| Schleswig-Holstein | 327 (11 to 2,345) | -38.3% (-85.7 to -5.9%) | 550 (180 to 2,870) |
| Thuringia | 999 (91 to 3,715) | -58.3% (-82.5 to -25.9%) | 1,569 (339 to 5,288) |
| Greece | 1,011 (0 to 3,191) | -12.9% (-35.8 to 0.0%) | 7,312 (6,175 to 9,524) |
| Hungary | 2,624 (822 to 4,433) | -24.0% (-37.5 to -9.7%) | 10,760 (8,287 to 13,921) |
| Iceland | 55 (2 to 174) | -54.1% (-87.2 to -11.2%) | 81 (16 to 232) |
| Ireland | 3,278 (881 to 9,273) | -31.9% (-53.1 to -14.1%) | 9,425 (5,793 to 17,793) |
| Israel | 1,181 (413 to 2,446) | -14.5% (-32.3 to -4.6%) | 8,489 (5,100 to 10,696) |
| Italy | 21,491 (7,538 to 51,432) | -16.7% (-28.2 to -8.4%) | 121,821 (87,133 to 202,584) |
| Piemonte | 3,147 (839 to 8,351) | -21.9% (-37.8 to -9.7%) | 13,292 (8,542 to 24,854) |
| Valle d'Aosta | 25 (0 to 179) | -7.5% (-31.5 to 0.0%) | 231 (154 to 548) |
| Liguria | 568 (72 to 2,261) | -10.4% (-25.9 to -2.8%) | 4,412 (2,493 to 10,417) |
| Lombardia | 4,858 (1,126 to 14,130) | -12.1% (-24.6 to -4.4%) | 36,233 (25,042 to 64,454) |
| Provincia autonoma di Bolzano | 632 (98 to 1,300) | -18.9% (-33.0 to -7.7%) | 3,535 (815 to 7,097) |
| Provincia autonoma di Trento | 520 (60 to 1,272) | -21.4% (-36.0 to -6.8%) | 2,190 (873 to 4,190) |
| Veneto | 1,261 (279 to 3,881) | -15.8% (-32.3 to -5.0%) | 7,282 (5,378 to 12,730) |
| Friuli-Venezia Giulia | 882 (151 to 2,509) | -31.9% (-52.8 to -12.2%) | 2,509 (1,251 to 4,920) |
| Emilia-Romagna | 2,266 (387 to 7,448) | -16.3% (-33.4 to -5.1%) | 12,118 (7,682 to 24,550) |
| Toscana | 2,034 (432 to 6,234) | -20.4% (-35.9 to -9.4%) | 9,031 (4,229 to 22,009) |
| Umbria | 73 (0 to 243) | -15.4% (-39.1 to 0.0%) | 503 (156 to 1,145) |
| Marche | 590 (73 to 2,368) | -14.8% (-32.6 to -3.7%) | 3,293 (1,893 to 8,644) |
| Lazio | 1,295 (445 to 2,646) | -16.2% (-26.7 to -7.5%) | 7,705 (5,695 to 10,650) |
| Abruzzo | 230 (49 to 887) | -12.1% (-27.3 to -3.7%) | 1,684 (1,243 to 3,438) |
| Molise | 61 (0 to 341) | -23.6% (-67.7 to -0.2%) | 157 (27 to 656) |
| Campania | 1,192 (268 to 3,067) | -14.6% (-25.8 to -5.9%) | 7,527 (4,324 to 13,315) |
| Puglia | 414 (0 to 1,110) | -12.3% (-34.2 to 0.0%) | 3,295 (2,731 to 4,438) |
| Basilicata | 66 (0 to 219) | -20.4% (-49.9 to 0.0%) | 305 (60 to 898) |
| Calabria | 69 (0 to 463) | -14.9% (-51.2 to -0.4%) | 263 (108 to 1,200) |
| Sicilia | 1,133 (340 to 2,938) | -20.0% (-35.3 to -8.5%) | 5,300 (3,748 to 8,716) |
| Sardegna | 175 (0 to 425) | -19.8% (-43.8 to 0.0%) | 953 (499 to 1,248) |
| Kazakhstan | 2,232 (759 to 6,582) | -29.5% (-56.6 to -15.3%) | 6,905 (4,790 to 12,017) |
| Kyrgyzstan | 2,509 (868 to 4,459) | -41.6% (-67.7 to -16.4%) | 6,094 (3,078 to 8,935) |
| Latvia | 429 (27 to 1,435) | -61.3% (-88.9 to -24.0%) | 600 (113 to 1,908) |
| Lithuania | 682 (63 to 1,389) | -47.7% (-76.6 to -13.8%) | 1,427 (301 to 2,654) |
| Luxembourg | 115 (0 to 266) | -25.9% (-54.4 to 0.0%) | 418 (176 to 600) |
| Malta | 257 (90 to 677) | -45.9% (-68.5 to -23.6%) | 555 (230 to 1,216) |
| Montenegro | 156 (10 to 318) | -23.1% (-43.9 to -1.6%) | 666 (519 to 793) |
| Netherlands | 11,984 (7,206 to 18,654) | -47.6% (-62.0 to -29.4%) | 25,101 (19,286 to 33,479) |
| North Macedonia | 498 (0 to 1,257) | -19.5% (-40.1 to 0.0%) | 2,452 (2,034 to 3,410) |
| Norway | 176 (15 to 776) | -29.4% (-71.8 to -5.0%) | 469 (299 to 1,077) |
| Poland | 8,040 (89 to 17,561) | -20.9% (-47.3 to -0.3%) | 37,870 (30,568 to 49,559) |
| Portugal | 3,303 (1,420 to 6,491) | -24.4% (-39.1 to -12.8%) | 13,136 (10,891 to 17,051) |
| Republic of Moldova | 464 (0 to 1,424) | -10.7% (-34.9 to 0.0%) | 4,345 (3,965 to 4,861) |
| Romania | 944 (0 to 3,070) | -4.5% (-14.4 to 0.0%) | 20,120 (18,198 to 22,376) |
| Russian Federation | 38,820 (8,738 to 66,611) | -32.1% (-56.1 to -7.1%) | 121,841 (114,758 to 128,944) |
| San Marino | 2 (1 to 8) | -4.9% (-15.7 to -1.4%) | 46 (44 to 53) |
| Serbia | 3,702 (1,484 to 6,084) | -70.3% (-83.7 to -50.3%) | 5,192 (2,663 to 7,883) |
| Slovakia | 1,046 (0 to 2,502) | -38.7% (-67.6 to 0.0%) | 2,745 (968 to 4,544) |
| Slovenia | 466 (0 to 1,367) | -22.6% (-53.9 to 0.0%) | 1,900 (1,275 to 3,751) |
| Spain | 401 (111 to 681) | -0.5% (-0.9 to -0.2%) | 73,697 (69,330 to 78,890) |
| Andalucia | 68 (0 to 185) | -1.4% (-3.4 to 0.0%) | 4,728 (2,814 to 7,435) |
| Aragon | 5 (0 to 37) | -0.2% (-1.5 to 0.0%) | 2,268 (1,929 to 2,825) |
| Cantabria | 2 (0 to 13) | -0.4% (-2.0 to 0.0%) | 612 (554 to 757) |
| Castilla-La Mancha | 7 (0 to 34) | -0.2% (-0.8 to 0.0%) | 4,110 (3,480 to 4,899) |
| Community of Madrid | 7 (0 to 87) | -0.1% (-0.5 to 0.0%) | 15,531 (14,414 to 16,430) |
| Extremadura | 3 (0 to 11) | -0.3% (-0.8 to 0.0%) | 1,281 (1,117 to 1,529) |
| Balearic Islands | 13 (0 to 33) | -1.3% (-3.5 to 0.0%) | 1,030 (664 to 1,490) |
| Canary Islands | 28 (6 to 59) | -2.1% (-4.3 to -0.4%) | 1,438 (867 to 2,311) |
| Asturias | 56 (14 to 109) | -1.6% (-3.2 to -0.6%) | 3,646 (1,781 to 5,919) |
| Murcia | 31 (0 to 78) | -1.7% (-3.7 to 0.0%) | 1,830 (1,414 to 2,560) |
| Castile and León | 25 (0 to 50) | -0.5% (-0.9 to 0.0%) | 4,513 (3,708 to 6,148) |
| Catalonia | 86 (0 to 198) | -0.4% (-0.9 to 0.0%) | 19,392 (17,893 to 22,364) |
| Ceuta | 1 (0 to 5) | -1.2% (-3.6 to 0.0%) | 135 (23 to 743) |
| Navarre | 8 (0 to 21) | -0.6% (-1.6 to 0.0%) | 1,221 (1,030 to 1,451) |
| Valencian Community | 23 (0 to 113) | -0.5% (-2.4 to 0.0%) | 4,800 (4,169 to 5,908) |
| Galicia | 17 (0 to 46) | -0.6% (-1.6 to 0.0%) | 2,808 (2,360 to 3,575) |
| Melilla | 3 (0 to 10) | -1.5% (-4.1 to -0.3%) | 296 (16 to 648) |
| Basque Country | 16 (0 to 57) | -0.5% (-1.6 to 0.0%) | 3,421 (2,514 to 4,011) |
| La Rioja | 0 (0 to 4) | 0.0% (-0.6 to 0.0%) | 637 (572 to 684) |
| Sweden | 7,124 (4,052 to 12,595) | -51.4% (-66.3 to -38.5%) | 13,563 (10,576 to 18,974) |
| Switzerland | 5,195 (694 to 18,400) | -39.7% (-69.2 to -8.0%) | 11,734 (7,465 to 26,782) |
| Tajikistan | 108 (19 to 386) | -37.8% (-70.3 to -14.4%) | 244 (125 to 553) |
| Turkey | 9,793 (0 to 17,435) | -18.2% (-31.7 to 0.0%) | 53,601 (47,235 to 57,687) |
| Ukraine | 12,641 (4,410 to 22,207) | -31.0% (-56.9 to -10.7%) | 41,226 (36,013 to 46,789) |
| United Kingdom | 54,917 (25,955 to 100,855) | -31.7% (-47.1 to -18.2%) | 168,540 (136,449 to 217,547) |
| Northern Ireland | 1,059 (342 to 2,336) | -29.7% (-46.6 to -12.8%) | 3,427 (2,431 to 5,443) |
| Scotland | 4,625 (1,714 to 8,669) | -30.4% (-45.9 to -16.5%) | 14,658 (10,126 to 19,836) |
| Wales | 1,485 (573 to 2,944) | -22.5% (-36.3 to -10.3%) | 6,440 (4,996 to 8,171) |
| England | 47,749 (21,849 to 89,092) | -32.2% (-48.1 to -18.1%) | 144,016 (116,422 to 190,937) |
| Uzbekistan | 780 (313 to 1,852) | -37.2% (-57.1 to -22.3%) | 2,018 (1,233 to 3,598) |
| Region of the Americas | 123,358 (70,931 to 174,474) | -11.8% (-16.4 to -6.9%) | 1,045,057 (987,186 to 1,098,209) |
| Argentina | 647 (275 to 1,646) | -1.4% (-3.5 to -0.6%) | 45,614 (41,519 to 51,590) |
| Bahamas | 79 (30 to 165) | -24.3% (-40.0 to -12.0%) | 313 (221 to 434) |
| Barbados | 1 (1 to 2) | -14.3% (-20.7 to -9.0%) | 10 (10 to 10) |
| Belize | 69 (12 to 165) | -33.9% (-56.8 to -13.7%) | 187 (85 to 316) |
| Bolivia (Plurinational State of) | 90 (48 to 145) | -1.0% (-1.5 to -0.5%) | 9,330 (9,092 to 9,633) |
| Brazil | 9,707 (6,097 to 14,524) | -5.2% (-7.5 to -3.4%) | 186,751 (178,214 to 195,088) |
| Acre | 13 (5 to 25) | -1.7% (-3.2 to -0.7%) | 731 (711 to 760) |
| Alagoas | 175 (81 to 291) | -6.7% (-10.6 to -3.4%) | 2,603 (2,436 to 2,762) |
| Amazonas | 618 (0 to 1,758) | -8.9% (-23.4 to 0.0%) | 6,636 (5,887 to 7,669) |
| Amapá | 42 (7 to 125) | -4.8% (-12.8 to -1.0%) | 840 (768 to 988) |
| Bahia | 421 (213 to 799) | -4.7% (-8.1 to -2.5%) | 8,929 (8,394 to 9,774) |
| Ceará | 201 (40 to 575) | -2.0% (-5.6 to -0.4%) | 9,753 (9,418 to 10,516) |
| Distrito Federal | 177 (100 to 314) | -3.9% (-6.5 to -2.3%) | 4,470 (4,200 to 4,813) |
| Espírito Santo | 286 (131 to 546) | -6.2% (-11.0 to -3.1%) | 4,570 (4,184 to 5,073) |
| Goiás | 416 (79 to 955) | -5.6% (-11.1 to -1.3%) | 7,208 (6,026 to 8,706) |
| Maranhão | 414 (202 to 675) | -8.6% (-13.3 to -4.6%) | 4,791 (4,412 to 5,167) |
| Minas Gerais | 827 (267 to 1,741) | -7.2% (-13.6 to -2.7%) | 11,146 (9,752 to 12,863) |
| Mato Grosso do Sul | 87 (24 to 197) | -4.5% (-9.1 to -1.4%) | 1,874 (1,673 to 2,175) |
| Mato Grosso | 203 (90 to 403) | -4.6% (-8.4 to -2.2%) | 4,383 (4,081 to 4,812) |
| Pará | 119 (20 to 433) | -1.7% (-5.8 to -0.3%) | 7,018 (6,796 to 7,587) |
| Paraíba | 452 (197 to 1,013) | -10.9% (-20.7 to -5.4%) | 4,049 (3,571 to 4,868) |
| Paraná | 456 (174 to 1,096) | -6.6% (-14.2 to -2.9%) | 6,683 (5,851 to 7,966) |
| Pernambuco | 406 (59 to 1,125) | -4.1% (-10.4 to -0.7%) | 9,532 (8,757 to 10,807) |
| Piauí | 467 (212 to 975) | -13.5% (-24.3 to -7.1%) | 3,378 (2,886 to 4,217) |
| Rio de Janeiro | 1,218 (417 to 2,673) | -5.0% (-10.1 to -1.9%) | 23,867 (21,613 to 26,942) |
| Rio Grande do Norte | 297 (51 to 862) | -9.1% (-22.4 to -1.9%) | 3,096 (2,686 to 3,910) |
| Rondônia | 105 (40 to 236) | -6.0% (-12.6 to -2.5%) | 1,690 (1,561 to 1,922) |
| Roraima | 82 (38 to 147) | -9.0% (-14.8 to -4.7%) | 906 (797 to 1,017) |
| Rio Grande do Sul | 633 (150 to 1,844) | -7.6% (-18.2 to -2.4%) | 7,821 (6,365 to 10,286) |
| Santa Catarina | 135 (37 to 431) | -3.6% (-10.3 to -1.1%) | 3,570 (3,230 to 4,236) |
| Sergipe | 195 (98 to 376) | -7.0% (-12.6 to -3.7%) | 2,742 (2,529 to 3,063) |
| São Paulo | 1,018 (285 to 2,212) | -2.3% (-4.8 to -0.7%) | 42,779 (40,303 to 46,440) |
| Tocantins | 240 (96 to 468) | -13.9% (-23.5 to -6.7%) | 1,683 (1,352 to 2,063) |
| Canada | 11,350 (4,457 to 23,687) | -28.0% (-42.0 to -15.0%) | 39,829 (22,778 to 65,957) |
| Alberta | 2,730 (1,089 to 7,455) | -59.6% (-79.0 to -36.2%) | 4,551 (2,039 to 11,060) |
| British Columbia | 160 (35 to 440) | -30.8% (-55.1 to -11.0%) | 473 (315 to 818) |
| Manitoba | 1,098 (17 to 7,163) | -52.8% (-87.3 to -12.7%) | 2,945 (68 to 18,524) |
| New Brunswick | 430 (1 to 2,341) | -32.8% (-69.5 to -4.8%) | 1,951 (11 to 10,535) |
| Nova Scotia | 2 (0 to 10) | -2.6% (-11.1 to -0.3%) | 70 (66 to 83) |
| Ontario | 3,593 (1,185 to 9,776) | -23.9% (-39.7 to -10.9%) | 14,395 (6,536 to 27,097) |
| Quebec | 3,262 (1,054 to 8,067) | -20.3% (-36.2 to -8.3%) | 15,333 (10,506 to 22,457) |
| Saskatchewan | 76 (0 to 660) | -32.6% (-91.4 to -0.9%) | 111 (25 to 783) |
| Chile | 107 (46 to 216) | -0.6% (-1.2 to -0.3%) | 16,635 (15,636 to 17,923) |
| Colombia | 2,757 (1,480 to 4,992) | -5.9% (-9.7 to -3.4%) | 46,438 (41,385 to 52,554) |
| Costa Rica | 377 (162 to 698) | -11.1% (-17.8 to -5.9%) | 3,361 (2,463 to 4,291) |
| Cuba | 240 (14 to 1,751) | -21.5% (-53.3 to -5.8%) | 721 (230 to 3,515) |
| Dominican Republic | 40 (5 to 117) | -1.6% (-4.5 to -0.2%) | 2,396 (2,244 to 2,648) |
| Ecuador | 51 (23 to 111) | -0.2% (-0.5 to -0.1%) | 20,713 (20,426 to 21,129) |
| El Salvador | 213 (0 to 827) | -7.5% (-22.9 to 0.0%) | 3,289 (1,250 to 4,763) |
| Guatemala | 315 (84 to 949) | -4.6% (-9.9 to -1.8%) | 6,388 (4,575 to 10,444) |
| Guyana | 14 (2 to 35) | -5.0% (-10.5 to -1.3%) | 247 (161 to 391) |
| Haiti | 31 (1 to 152) | -9.8% (-38.0 to -0.6%) | 268 (232 to 406) |
| Honduras | 308 (64 to 781) | -7.5% (-15.8 to -2.0%) | 3,867 (2,988 to 5,238) |
| Jamaica | 44 (7 to 119) | -8.8% (-17.2 to -2.7%) | 455 (273 to 795) |
| Mexico | 13,148 (6,521 to 21,395) | -9.0% (-14.2 to -4.3%) | 145,982 (131,662 to 156,820) |
| Aguascalientes | 71 (25 to 164) | -5.3% (-11.4 to -1.7%) | 1,373 (990 to 1,758) |
| Baja California | 364 (0 to 698) | -6.2% (-11.4 to 0.0%) | 5,829 (4,980 to 6,534) |
| Baja California Sur | 24 (0 to 89) | -3.0% (-10.2 to 0.0%) | 689 (574 to 928) |
| Campeche | 6 (0 to 19) | -0.7% (-2.0 to 0.0%) | 894 (859 to 951) |
| Coahuila | 149 (0 to 543) | -3.4% (-12.6 to 0.0%) | 4,143 (3,550 to 4,756) |
| Colima | 77 (40 to 126) | -5.8% (-9.3 to -2.8%) | 1,331 (1,144 to 1,577) |
| Chiapas | 1,078 (1 to 3,232) | -27.5% (-56.3 to -0.1%) | 3,114 (1,076 to 7,188) |
| Chihuahua | 362 (0 to 657) | -7.2% (-14.0 to 0.0%) | 5,037 (3,953 to 6,363) |
| Mexico City | 699 (325 to 1,240) | -3.6% (-6.2 to -1.7%) | 19,428 (18,260 to 20,727) |
| Durango | 203 (40 to 472) | -14.1% (-27.2 to -4.2%) | 1,362 (899 to 1,942) |
| Guanajuato | 987 (162 to 2,063) | -16.4% (-29.3 to -4.3%) | 5,744 (3,748 to 7,597) |
| Guerrero | 618 (197 to 1,165) | -16.4% (-27.9 to -7.2%) | 3,661 (2,677 to 4,534) |
| Hidalgo | 439 (210 to 729) | -11.3% (-17.8 to -5.8%) | 3,872 (3,225 to 4,439) |
| Jalisco | 817 (0 to 2,301) | -10.1% (-24.4 to 0.0%) | 8,210 (5,528 to 10,438) |
| México | 1,762 (0 to 4,433) | -10.2% (-21.3 to 0.0%) | 16,867 (12,214 to 22,609) |
| Michoacán de Ocampo | 604 (0 to 1,269) | -12.6% (-24.7 to 0.0%) | 4,890 (3,551 to 5,681) |
| Morelos | 365 (121 to 675) | -17.4% (-29.3 to -7.2%) | 2,051 (1,492 to 2,533) |
| Nayarit | 163 (0 to 384) | -10.3% (-23.0 to 0.0%) | 1,587 (1,200 to 1,841) |
| Nuevo León | 344 (0 to 1,240) | -4.8% (-17.2 to 0.0%) | 7,096 (6,168 to 8,036) |
| Oaxaca | 638 (170 to 1,199) | -17.3% (-30.5 to -5.7%) | 3,664 (2,192 to 4,718) |
| Puebla | 983 (253 to 1,940) | -13.0% (-22.9 to -4.6%) | 7,329 (5,469 to 9,045) |
| Querétaro | 248 (0 to 552) | -10.5% (-22.1 to 0.0%) | 2,374 (1,708 to 2,819) |
| Quintana Roo | 68 (0 to 170) | -2.6% (-6.1 to 0.0%) | 2,625 (2,339 to 2,996) |
| San Luis Potosí | 245 (0 to 597) | -6.7% (-15.4 to 0.0%) | 3,610 (3,099 to 4,137) |
| Sinaloa | 294 (117 to 536) | -6.1% (-10.5 to -2.8%) | 4,722 (4,082 to 5,397) |
| Sonora | 315 (0 to 671) | -7.0% (-14.0 to 0.0%) | 4,522 (3,485 to 5,169) |
| Tabasco | 66 (18 to 145) | -2.0% (-4.1 to -0.6%) | 3,298 (3,097 to 3,592) |
| Tamaulipas | 117 (12 to 340) | -3.5% (-9.0 to -0.4%) | 3,135 (2,676 to 4,023) |
| Tlaxcala | 157 (0 to 339) | -8.4% (-17.6 to 0.0%) | 1,886 (1,492 to 2,163) |
| Veracruz de Ignacio de la Llave | 584 (148 to 1,448) | -8.4% (-18.2 to -2.7%) | 6,583 (5,409 to 8,342) |
| Yucatán | 84 (0 to 233) | -2.6% (-7.2 to 0.0%) | 3,211 (2,794 to 3,789) |
| Zacatecas | 216 (0 to 429) | -11.6% (-21.8 to 0.0%) | 1,848 (1,458 to 2,162) |
| Nicaragua | 725 (116 to 2,418) | -53.2% (-79.2 to -28.2%) | 1,205 (386 to 3,431) |
| Panama | 119 (58 to 213) | -2.5% (-4.4 to -1.3%) | 4,692 (4,046 to 5,403) |
| Paraguay | 195 (93 to 383) | -8.3% (-13.8 to -4.6%) | 2,313 (1,946 to 2,880) |
| Peru | 75 (12 to 241) | -0.2% (-0.5 to 0.0%) | 44,830 (43,857 to 46,523) |
| Suriname | 51 (3 to 191) | -19.7% (-48.4 to -2.5%) | 212 (120 to 463) |
| Trinidad and Tobago | 35 (11 to 88) | -14.8% (-28.2 to -6.6%) | 221 (161 to 325) |
| United States of America | 82,150 (46,079 to 116,120) | -18.0% (-25.1 to -10.1%) | 456,375 (424,449 to 489,680) |
| Alabama | 1,641 (702 to 3,007) | -22.5% (-38.9 to -10.3%) | 7,327 (4,609 to 9,522) |
| Alaska | 93 (9 to 321) | -36.6% (-69.0 to -9.8%) | 211 (95 to 500) |
| Arizona | 1,572 (0 to 3,036) | -15.6% (-28.7 to 0.0%) | 10,123 (7,786 to 13,035) |
| Arkansas | 432 (0 to 1,267) | -10.6% (-31.7 to 0.0%) | 4,075 (3,476 to 4,659) |
| California | 10,847 (4,969 to 17,611) | -26.1% (-39.3 to -12.8%) | 41,492 (28,290 to 51,411) |
| Colorado | 1,693 (432 to 3,131) | -30.6% (-49.4 to -12.7%) | 5,444 (2,936 to 7,494) |
| Connecticut | 1,653 (280 to 4,913) | -16.5% (-36.0 to -3.8%) | 9,144 (7,209 to 14,005) |
| Delaware | 161 (17 to 318) | -13.2% (-25.9 to -1.2%) | 1,243 (954 to 1,445) |
| District of Columbia | 20 (0 to 98) | -2.7% (-12.1 to 0.0%) | 687 (643 to 841) |
| Florida | 5,402 (0 to 9,274) | -18.6% (-30.8 to 0.0%) | 29,050 (22,259 to 31,933) |
| Georgia | 3,221 (33 to 5,226) | -20.9% (-34.0 to -0.2%) | 15,497 (13,905 to 17,171) |
| Hawaii | 251 (40 to 591) | -29.2% (-49.3 to -10.8%) | 787 (355 to 1,350) |
| Idaho | 900 (67 to 2,399) | -34.7% (-57.4 to -3.4%) | 2,472 (1,624 to 4,497) |
| Illinois | 1,924 (0 to 4,846) | -9.1% (-22.1 to 0.0%) | 20,807 (19,124 to 23,122) |
| Indiana | 2,125 (719 to 3,871) | -19.5% (-33.1 to -7.4%) | 10,772 (9,489 to 12,811) |
| Iowa | 1,161 (163 to 2,245) | -24.8% (-44.6 to -3.9%) | 4,653 (3,753 to 5,818) |
| Kansas | 1,156 (296 to 2,380) | -27.7% (-47.2 to -8.4%) | 4,110 (2,946 to 5,782) |
| Kentucky | 721 (0 to 1,978) | -13.2% (-33.2 to 0.0%) | 5,110 (4,158 to 7,037) |
| Louisiana | 1,054 (0 to 2,111) | -11.1% (-22.9 to 0.0%) | 9,502 (8,901 to 10,445) |
| Maine | 95 (4 to 455) | -25.6% (-64.5 to -2.8%) | 282 (156 to 722) |
| Maryland | 1,492 (656 to 2,362) | -18.4% (-29.9 to -7.9%) | 8,142 (6,899 to 9,614) |
| Massachusetts | 830 (0 to 2,556) | -5.2% (-14.9 to 0.0%) | 15,397 (14,247 to 17,332) |
| Michigan | 1,370 (0 to 3,559) | -8.7% (-20.6 to 0.0%) | 15,329 (13,874 to 17,904) |
| Minnesota | 693 (0 to 1,794) | -10.1% (-27.1 to 0.0%) | 6,771 (6,163 to 7,876) |
| Mississippi | 714 (0 to 1,402) | -13.2% (-26.3 to 0.0%) | 5,418 (4,972 to 5,807) |
| Missouri | 1,725 (0 to 2,988) | -25.1% (-43.0 to 0.0%) | 6,938 (5,043 to 9,405) |
| Montana | 854 (158 to 2,189) | -36.6% (-59.2 to -12.6%) | 2,215 (1,078 to 4,738) |
| Nebraska | 785 (53 to 1,883) | -29.0% (-51.0 to -2.5%) | 2,586 (1,927 to 4,174) |
| Nevada | 805 (351 to 1,352) | -21.7% (-34.7 to -8.2%) | 3,752 (2,655 to 4,826) |
| New Hampshire | 693 (42 to 2,253) | -36.3% (-61.2 to -7.5%) | 1,645 (547 to 4,040) |
| New Jersey | 2,797 (0 to 6,941) | -11.5% (-22.4 to 0.0%) | 23,654 (19,210 to 30,875) |
| New Mexico | 935 (288 to 2,209) | -22.5% (-38.8 to -9.4%) | 3,919 (2,913 to 5,939) |
| New York | 2,001 (248 to 5,895) | -5.1% (-13.9 to -0.7%) | 37,152 (34,014 to 42,900) |
| North Carolina | 2,917 (0 to 7,239) | -19.5% (-41.8 to 0.0%) | 14,354 (11,699 to 20,333) |
| North Dakota | 168 (48 to 346) | -17.5% (-32.4 to -6.5%) | 920 (687 to 1,236) |
| Ohio | 3,354 (0 to 9,408) | -23.0% (-41.3 to 0.0%) | 14,065 (7,702 to 23,896) |
| Oklahoma | 822 (0 to 1,788) | -20.0% (-45.7 to 0.0%) | 4,118 (3,528 to 5,099) |
| Oregon | 1,111 (311 to 2,044) | -35.6% (-57.2 to -17.4%) | 3,138 (1,280 to 4,764) |
| Pennsylvania | 4,686 (1,664 to 9,809) | -21.3% (-37.4 to -8.4%) | 21,574 (17,997 to 27,933) |
| Rhode Island | 267 (0 to 774) | -10.7% (-25.9 to 0.0%) | 2,276 (1,885 to 3,114) |
| South Carolina | 840 (0 to 2,267) | -11.1% (-30.3 to 0.0%) | 7,491 (6,669 to 8,838) |
| South Dakota | 244 (0 to 489) | -22.9% (-41.0 to 0.0%) | 1,038 (766 to 1,449) |
| Tennessee | 1,741 (0 to 3,224) | -22.7% (-40.9 to 0.0%) | 7,782 (4,942 to 10,050) |
| Texas | 6,967 (453 to 11,797) | -19.5% (-33.5 to -1.2%) | 36,092 (32,097 to 37,875) |
| Utah | 897 (254 to 1,552) | -34.4% (-59.8 to -11.9%) | 2,689 (1,086 to 3,713) |
| Vermont | 4 (1 to 12) | -5.6% (-14.4 to -1.8%) | 69 (65 to 79) |
| Virginia | 1,831 (9 to 3,684) | -20.8% (-40.3 to -0.1%) | 8,992 (5,102 to 11,915) |
| Washington | 1,895 (933 to 3,085) | -29.0% (-43.8 to -15.5%) | 6,501 (4,972 to 7,792) |
| West Virginia | 564 (85 to 977) | -35.2% (-53.7 to -5.1%) | 1,623 (951 to 2,516) |
| Wisconsin | 1,849 (269 to 3,439) | -23.9% (-41.7 to -4.1%) | 7,566 (6,106 to 9,363) |
| Wyoming | 176 (14 to 611) | -41.7% (-69.8 to -13.9%) | 377 (100 to 1,137) |
| Uruguay | 6 (1 to 20) | -7.0% (-19.9 to -1.3%) | 72 (58 to 104) |
| Venezuela (Bolivarian Republic of) | 414 (183 to 846) | -17.3% (-28.9 to -9.0%) | 2,342 (1,859 to 3,101) |
| South-East Asia Region | 121,638 (72,475 to 176,995) | -30.5% (-41.6 to -19.6%) | 397,289 (345,640 to 470,913) |
| Bangladesh | 4,423 (2,249 to 8,156) | -36.6% (-53.0 to -24.0%) | 11,781 (8,953 to 16,187) |
| India | 104,301 (60,679 to 151,873) | -31.2% (-42.3 to -19.9%) | 332,258 (285,297 to 396,931) |
| Andhra Pradesh | 2,252 (1,192 to 4,009) | -20.6% (-32.1 to -12.6%) | 10,764 (8,870 to 13,079) |
| Arunachal Pradesh | 375 (70 to 732) | -52.2% (-84.7 to -20.0%) | 769 (128 to 1,221) |
| Assam | 1,267 (187 to 3,910) | -39.8% (-65.4 to -14.2%) | 2,841 (1,283 to 6,191) |
| Bihar | 9,069 (1,081 to 42,104) | -66.8% (-92.6 to -39.9%) | 11,687 (2,663 to 45,700) |
| Chhattisgarh | 5,160 (2,652 to 9,277) | -48.8% (-66.5 to -30.3%) | 10,467 (6,633 to 15,253) |
| Delhi | 2,888 (1,541 to 4,581) | -20.3% (-30.9 to -11.2%) | 14,187 (12,621 to 16,034) |
| Goa | 174 (80 to 310) | -15.2% (-24.3 to -8.3%) | 1,130 (864 to 1,381) |
| Gujarat | 2,757 (1,036 to 6,313) | -28.2% (-47.2 to -14.2%) | 9,329 (6,559 to 13,680) |
| Haryana | 1,516 (139 to 4,338) | -27.8% (-52.0 to -6.4%) | 4,691 (2,138 to 8,873) |
| Himachal Pradesh | 1,114 (320 to 2,443) | -46.3% (-67.1 to -27.7%) | 2,326 (973 to 4,217) |
| Jammu & Kashmir and Ladakh | 1,103 (457 to 2,290) | -31.3% (-47.9 to -18.7%) | 3,400 (2,345 to 5,008) |
| Jharkhand | 1,338 (401 to 3,242) | -43.4% (-65.8 to -25.2%) | 2,897 (1,555 to 5,266) |
| Karnataka | 3,937 (1,279 to 7,934) | -18.6% (-30.9 to -8.6%) | 20,426 (14,613 to 27,609) |
| Kerala | 4,013 (1,446 to 7,893) | -28.4% (-45.1 to -15.3%) | 13,783 (7,303 to 20,098) |
| Madhya Pradesh | 4,728 (1,433 to 12,022) | -42.0% (-65.9 to -23.2%) | 10,447 (5,617 to 20,250) |
| Maharashtra | 9,055 (4,453 to 16,138) | -12.2% (-19.5 to -7.0%) | 73,657 (60,033 to 87,612) |
| Manipur | 612 (165 to 1,056) | -22.6% (-38.0 to -7.4%) | 2,666 (2,045 to 3,286) |
| Meghalaya | 384 (20 to 879) | -22.3% (-59.2 to -1.2%) | 1,785 (1,270 to 2,299) |
| Nagaland | 561 (206 to 926) | -60.3% (-86.1 to -25.5%) | 967 (343 to 1,558) |
| Odisha | 8,536 (2,962 to 16,109) | -55.8% (-76.7 to -35.0%) | 14,773 (7,170 to 23,726) |
| Punjab | 1,197 (343 to 3,245) | -17.5% (-36.1 to -6.9%) | 6,362 (4,996 to 9,327) |
| Rajasthan | 8,632 (3,132 to 19,753) | -52.5% (-74.0 to -31.6%) | 15,556 (8,841 to 28,199) |
| Sikkim | 101 (44 to 170) | -17.2% (-41.5 to -8.6%) | 601 (376 to 778) |
| Tamil Nadu | 4,778 (1,321 to 10,649) | -23.5% (-41.4 to -9.5%) | 19,249 (13,975 to 26,437) |
| Telangana | 1,076 (379 to 2,393) | -31.0% (-50.4 to -16.5%) | 3,298 (2,177 to 5,038) |
| Tripura | 410 (186 to 816) | -40.9% (-59.7 to -25.8%) | 972 (684 to 1,420) |
| Uttar Pradesh | 4,598 (2,482 to 9,857) | -31.2% (-48.4 to -21.1%) | 14,325 (10,929 to 21,680) |
| Uttarakhand | 1,662 (276 to 3,515) | -36.3% (-58.4 to -15.7%) | 4,316 (1,654 to 6,787) |
| West Bengal | 21,010 (11,694 to 31,455) | -38.6% (-58.0 to -20.1%) | 54,583 (44,352 to 60,015) |
| Indonesia | 2,518 (1,335 to 4,881) | -11.1% (-18.7 to -6.4%) | 22,434 (19,590 to 26,666) |
| Maldives | 20 (1 to 92) | -15.7% (-41.7 to -2.2%) | 91 (45 to 271) |
| Myanmar | 6,451 (2,609 to 13,658) | -44.6% (-64.6 to -26.3%) | 14,178 (7,621 to 23,573) |
| Nepal | 3,623 (0 to 9,217) | -23.7% (-67.8 to 0.0%) | 16,041 (11,297 to 19,520) |
| Sri Lanka | 268 (0 to 3,487) | -29.9% (-81.0 to -1.2%) | 405 (16 to 4,377) |
| Thailand | 34 (6 to 168) | -27.0% (-68.3 to -8.5%) | 101 (68 to 247) |
| Western Pacific Region | 14,031 (3,207 to 35,638) | -21.8% (-43.3 to -9.0%) | 58,890 (33,758 to 100,123) |
| Australia | 72 (32 to 148) | -5.4% (-10.3 to -2.6%) | 1,317 (1,231 to 1,457) |
| China | 1,467 (41 to 18,772) | -10.7% (-72.2 to -0.8%) | 6,680 (4,819 to 25,720) |
| Hong Kong Special Administrative Region of China | 166 (1 to 1,355) | -16.1% (-52.7 to -0.5%) | 542 (109 to 3,707) |
| Beijing | 32 (1 to 211) | -42.0% (-86.7 to -6.2%) | 49 (11 to 243) |
| Chongqing | 4 (1 to 16) | -27.2% (-65.2 to -10.1%) | 12 (9 to 26) |
| Guangdong | 29 (3 to 127) | -53.8% (-87.6 to -20.8%) | 45 (15 to 151) |
| Hainan | 7 (1 to 54) | -28.5% (-80.7 to -11.9%) | 16 (9 to 70) |
| Hebei | 8 (2 to 27) | -43.0% (-72.9 to -21.8%) | 17 (11 to 37) |
| Heilongjiang | 1 (0 to 2) | -5.7% (-11.5 to -2.2%) | 15 (15 to 17) |
| Hubei | 1,175 (3 to 18,652) | -6.5% (-72.7 to -0.1%) | 5,915 (4,565 to 25,385) |
| Shandong | 35 (4 to 157) | -58.8% (-89.4 to -27.4%) | 49 (15 to 178) |
| Shanghai | 10 (2 to 35) | -43.2% (-76.3 to -19.6%) | 20 (12 to 47) |
| Japan | 6,373 (1,226 to 19,648) | -22.9% (-39.7 to -10.6%) | 24,607 (10,311 to 61,165) |
| Malaysia | 3,232 (318 to 9,074) | -47.3% (-71.2 to -25.3%) | 6,227 (1,108 to 16,112) |
| New Zealand | 1 (0 to 6) | -2.5% (-19.4 to -0.1%) | 27 (26 to 33) |
| Papua New Guinea | 0 (0 to 1) | -1.8% (-9.5 to -0.1%) | 9 (8 to 11) |
| Philippines | 2,233 (285 to 6,228) | -12.0% (-25.1 to -3.1%) | 16,708 (9,036 to 29,294) |
| Republic of Korea | 415 (73 to 1,664) | -12.1% (-23.7 to -5.1%) | 2,969 (1,240 to 8,514) |
| Singapore | 6 (0 to 40) | -7.6% (-27.4 to -0.5%) | 52 (29 to 156) |
| Viet Nam | 231 (5 to 1,757) | -52.7% (-94.3 to -10.3%) | 294 (47 to 1,903) |

**Figure 5.**
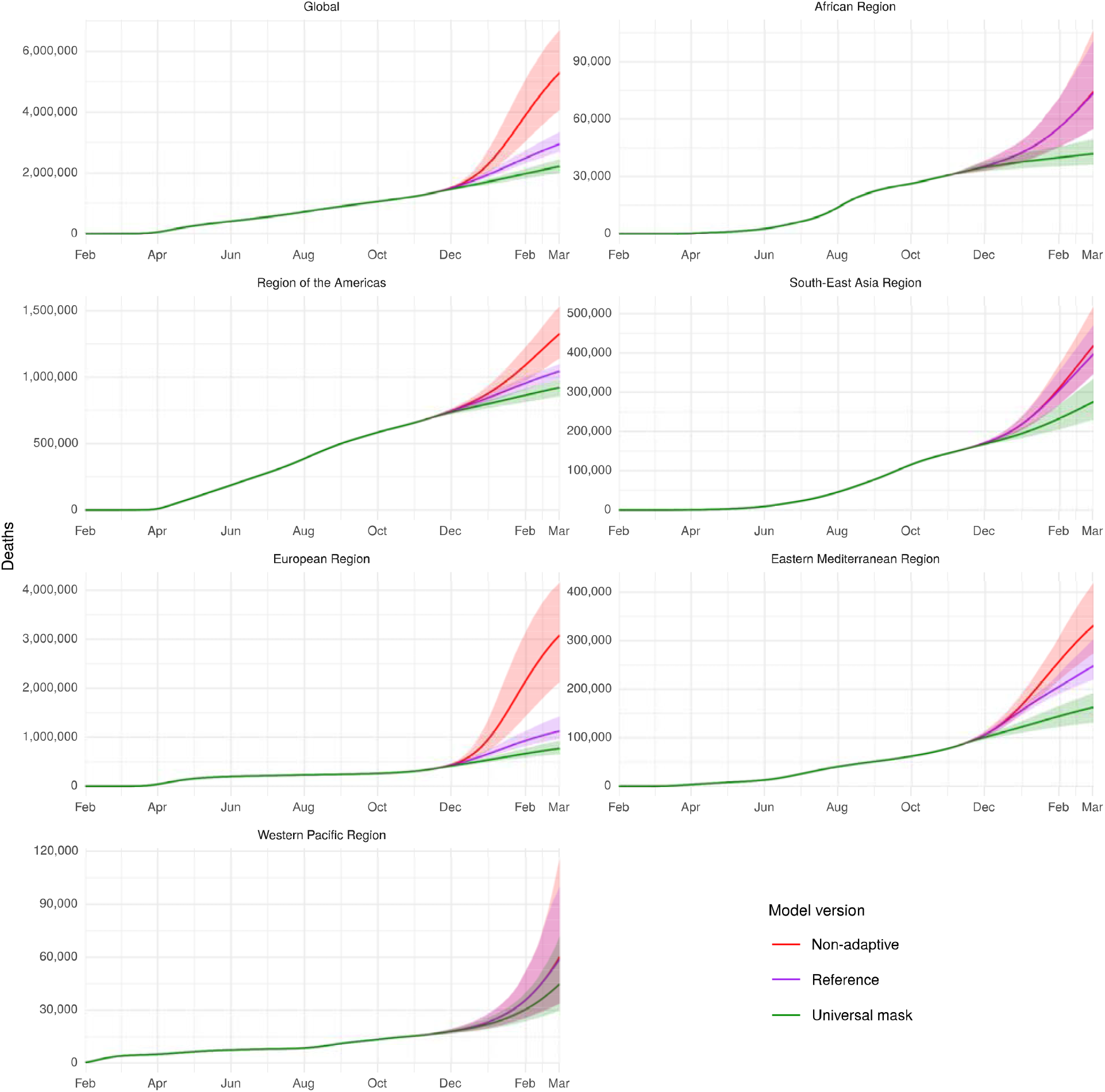
Projected global deaths due to COVID-19 up to March 01, 2021 in the scenario for current projections of mask use, the non-adaptive scenario, and the scenario with 95% coverage of mask use. Projections are shown for the world and for the six WHO regions.

Figure 6 shows a map of the percent reduction in expected deaths from the reference scenario compared to the mask use scenario in the deaths from October 27, 2020 to March 01, 2021. The largest percent reduction in expected deaths occurred in Belarus (76.8% difference, 95% UI 66.9 to 82.3%), Denmark (76.1% difference, 95% UI 61.1 to 87.3%), and Uganda (75.4% difference, 95% UI 38.4 to 97.0%). There are large variations in the number of lives saved globally, regionally, nationally, and subnationally and these are detailed in full in Table 1.

**Figure 6.**
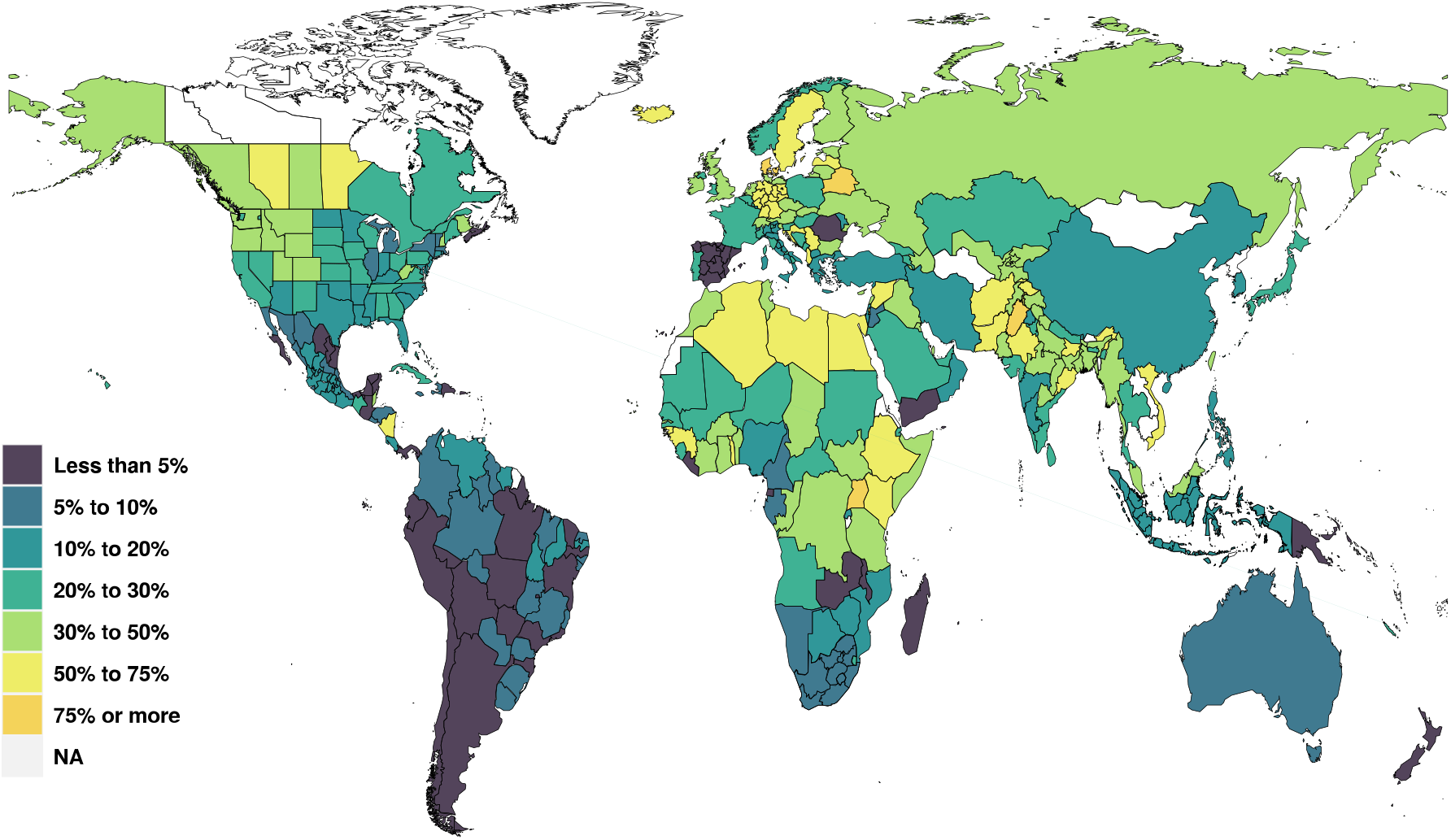
Percent reduction in cumulative deaths on March 01, 2021 in the universal mask use scenario and the reference scenario. Areas have “NA” either because of no available data or because the population size, cases or deaths are so small that the SEIR models do not run.

## Discussion

Given global mask use is currently at 59.0%, increasing mask use to 95% could decrease cumulative COVID-19 deaths by March 01,2021 by 0.73 million deaths (95% UI 0.39 to 1.11 million). This represents a 24.8% reduction in the number of deaths expected from October 27, 2020 to March 01, 2021. Our models also show that in many settings, increased mask use could delay the need re-imposition of SDM; in addition to the lives saved, a delay in re-imposition of social mandates might also be accompanied by substantial economic benefits, although this was not specifically modelled.

The published studies of SARS-CoV-2 included in our analysis of mask effectiveness (see Supplementary Appendix for details) demonstrated reductions in relative risk of 30% - 100%, with the one study of non-medical mask use amongst the general population, indicating a reduction of 42% for any mask use and 70% for consistent mask use.^27^ Mask effectiveness is also supported by additional evidence from case studies^28–30^ and from laboratory studies that report on the efficacy of masks in reducing exhalation of both aerosols and droplets by those infected with SARS CoV-2.^31^ Based on all types of available evidence, it seems wise to encourage mask use throughout the world as the benefits can be substantial with low to zero contraindications. Guidance for specific materials, handling of face coverings and other considerations is rapidly evolving as additional evidence emerges.^32^

Given that the cost of masks is very low, mask mandates and/or the promotion of mask use seems prudent, as the risk of adopting these policies is minimal and the potential benefits very large. In recognition of this, over the last few months we have seen the number of countries and territories with mask mandates in place increase substantially. Nevertheless, there remains reluctance to adopt mask use and to impose mask mandates. In some settings, the current epidemiological context means that mask wearing is not viewed as a necessary part of control, such as in Norway where the Norwegian Institute of Public Health determined that, given their current low prevalence, “200,000 people would need to wear facemasks to prevent one new infection per week”.^33^ In other settings however, despite increasing cases, public sentiment towards mask wearing hinders universal use. Past messages from some governments have not encouraged mask use and may have actually discouraged mask use.^34–36^ Early on, WHO stated “*the wide use of masks by healthy people in the community setting is not supported by current evidence and carries uncertainties and critical risks*”^37^ and only changed their official position on June 5^th^ to encourage mask use.^38^ For those decision-makers who are concerned with the economic effects of SDM, mask use provides a low-cost strategy to reduce the risk of a further round of SDM and the associated unemployment and economic downturn. Mask use scaled up rapidly in some locations following mandated use. For example, at the start of July, mask use in Victoria state Australia was less than 10% but following a mask mandate in that month, mask use exceeded 90% in early August.

While the effective R, the number of new infections created by a single infection under the auspices of control, can potentially be reduced by one-third through universal mask use compared to a reference scenario, mask use alone will likely be insufficient to control the epidemic in many locations. Even with universal mask use, we expect the global death toll due to COVID-19 to reach as high as 1.71 million deaths by the end of the year, and many more in 2021, assuming an efficacious vaccine is not widely deployed in the interim. Despite encouraging news of candidates,^6,7^ safety procedures, regulatory approval, and the problems of manufacturing and distribution at scale will likely delay the potential for widespread impact.^8^ Countries will have to consider other policy strategies to reduce transmission, including increasing testing, contact tracing, and isolation, along with “smart mandates”, that target SDM or restrictions to particular subgroups of the population, such as specific age-groups or local communities for short periods of time. A central policy challenge for many countries is understanding which of these mandates targeting strategies makes the most sense in a given context and at what level of COVID-19 transmission. This is the focus of our ongoing work.

The findings of this study should be interpreted while taking into account its limitations (further elaborated in the Supplementary Appendix): (i) the number of studies on general mask use in the population remains low; (ii) all the assumption of any type of SEIR model are also applicable here;^24^ prediction accuracy is highly spatially variable and compound into the future;^25^ (iv) death reporting globally is subject to regionally and nationally specific errors and biases for which we cannot fully control; (v) models is sensitive to death trend in last 7-14 days and (vi) self-reported mask use by surveys will also have an set of errors and biases that are not completely known.

Universal mask use can save many lives and avoid or delay the need for re-imposition of SDM. This will contribute to ameliorating the negative effects of COVID-19 on national economies. Until an affordable and effective vaccine becomes universally available, we find encouraging or mandating mask use is the least disruptive policy option available to countries currently experiences resurgences in infections and deaths. Simple face coverings are cheap and effective; one of the few available interventions that is widely available to everyone. Countries with currently low mask use will need to determine the optimal balance between encouraging the use of masks through advocacy and information about their benefits, and governance of a compulsory use associated with penalties for non-compliance.^39^

## Data Availability

Data are available to researchers and where not freely available, information on how to access the data is provided.

## Code Availability Statement

All code used for these analyses is publicly available online (http://github.com/ihmeuw/).

## Online content

Results for each scenario are accessible through a visualization tool at http://covid19.healthdata.org. The estimates presented in this tool will be iteratively updated as new data are incorporated and will ultimately supersede the results in this paper.

## Acknowledgements

We thank the various Departments of Health and frontline health professionals who are not only responding to this epidemic daily, but also provide the necessary data to inform this work – IHME wishes to warmly acknowledge the support of these and others who have made our COVID-19 estimation efforts possible (http://www.healthdata.org/covid/acknowledgements). This work was supported in part by the Bill & Melinda Gates Foundation, Bloomberg Philanthropies, as well as funding from the state of Washington and the National Science Foundation (2031096). AD Flaxman was supported in part by funding from the National Science Foundation (award DMS-1839116). We also extend a note of particular thanks to John Stanton and Julie Nordstrom for their generous support. We are grateful to Professor Wei Huang from Peking University for helping us extract information from scientific papers in Chinese.

