## Supplemental Material for "Global projections of lives saved from COVID-19 with universal mask use"

This appendix provides further methodological detail and additional results for “Global projections of potential lives saved from COVID-19 through universal mask use.”

Portions of this appendix have been reproduced or adapted from the supplementary information to Reiner et al.^1^ A reference has been provided at the top of each section that includes reproduced text.

### Author Contributions

CJL Murray, E Gakidou, SS Lim, RC Reiner, and DM Pigott conceived and planned the study.

DM Piggot obtained, extracted, processed, and curated input data for the SEIR model; E Frame extracted and analysed all data on the effectiveness of masks. SS Lim, A Carter, E Castro, L Woyczynski, H Lescinsky, and CE Troeger constructed covariate data layers. E Frame, A Deen, RM Barber, RC Reiner, RJD Sorenson, JK Collins, L Xu, E Linebarger, AY Aravkin, NJ Henry, X Dai, SS Lim, A Carter, L Woyczynski, H Lescinsky, and CE Troeger wrote the computer code and designed the statistical analyses.

E Frame, CE Troeger, and R Ma designed and prepared figures, conducted data quality checks, and verified estimates.

E Gakidou, M Brauer, CJL Murray, T Vos, LB Marczak and SI Hay wrote the first draft of the manuscript, and all authors contributed to subsequent revisions.

All authors provided intellectual input into aspects of this study.

### List of Investigators

| **First Name** | **Last Name** |
| --- | --- |
| Emmanuela | Gakidou |
| David M. | Pigott |
| Christopher E. | Troeger |
| Erin N. | Hulland |
| Robert C. | Reiner Jr |
| Christopher | Adolph |
| Aleksandr Y. | Aravkin |
| Steven D. | Bachmeier |
| Bree | Bang-Jensen |
| Ryan M. | Barber |
| Catherine | Bisignano |
| Sabina | Bloom |
| Austin | Carter |
| Emma | Castro |
| Suman | Chakrabarti |
| Jhilik | Chattopadhyay |
| Christine | Chen |
| Rebecca M. | Cogen |
| James K. | Collins |
| Emily | Combs |
| Natalie Maria | Cormier |
| Xiaochen | Dai |
| William James | Dangel |
| Amanda | Deen |
| Samuel B. | Ewald |
| Abraham D. | Flaxman |
| Nancy | Fullman |
| Gaorui | Guo |
| Kathryn | Hackman |
| Jiawei | He |
| Nathaniel J. | Henry |
| Casey | Johanns |
| Samantha Leigh | Larson |
| Alice | Lazzar-Atwood |
| Kate E. | LeGrand |
| Haley | Lescinsky |
| Stephen S. | Lim |
| Emily | Linebarger |
| Rafael | Lozano |
| Rui | Ma |
| Beatrice | Magistro |
| Johan C. | Månsson |
| Laurie B. | Marczak |
| Molly K. | Miller-Petrie |
| Ali H. | Mokdad |
| Meghan D. | Mooney |
| Christopher M. | Odell |
| James K. | O'Halloran |
| Samuel M. | Ostroff |
| Maja | Pasovic |
| Disha J. | Patel |
| Louise | Penberthy |
| Rebecca E. | Ramshaw |
| Gregory A. | Roth |
| David H. | Shaw |
| Brittney S. | Sheena |
| Aleksei | Sholokhov |
| Reed J. D. | Sorensen |
| Gianna | Sparks |
| Emma Elizabeth | Spurlock |
| Michelle L. | Subart |
| Ally | Walker |
| Alexandrea | Watson |
| Catherine A. | Welgan |
| Kirsten E. | Wiens |
| Lauren | Woyczynski |
| Liming | Xu |
| Simon I. | Hay |
| Michael | Brauer |
| Theo | Vos |
| Christopher J. L. | Murray |

### Guidelines for Accurate and Transparent Health Estimates Reporting (GATHER)

This study complies with the Guidelines for Accurate and Transparent Health Estimates Reporting (GATHER) recommendations.^2^ We have documented the steps in our analytical procedures and detailed the data sources used. See Table S4 for the GATHER checklist.

### Section 1. Estimating the effectiveness of masks in preventing transmission

#### Section 1.1. Overview

We performed a meta-regression of 63 observations from 40 studies of the effectiveness of masks in preventing the transmission of respiratory virus infections (Table S1). One study of mask use for COVID-19 in the general population was included,^3^ while the others were identified through two published meta-analyses.^4,5^ The studies varied in setting (general population versus healthcare), type of mask (which we dichotomized into medical-grade masks, including surgical and N95 masks, and non-medical masks, including cloth masks), comparator group (no mask use or “occasional” mask use), type of diagnosis (clinical or laboratory), country of study (dichotomized into Asian and non-Asian countries), and type of respiratory virus (SARS-CoV-1 or 2 versus H_1_N_1_, influenza, or other respiratory viruses); details of all included studies^3,6–44^ can be found in Table S2. From the identified papers, we extracted all relevant observations that assessed mask effectiveness, allowing for multiple observations per study based on variations in mask type, virus studied, or comparison group. Four studies identified from the two meta-analyses were excluded from our analysis because relative risk was not available,^45,46^ because we were unable to extract mask use from general PPE use,^47^ or because the comparison group was of less protective masks rather than no or infrequent mask use.^48^ In order to derive the most relevant pooled estimate for the effect of mask use on preventing the spread of COVID-19 in populations, we performed a meta-regression of all 63 observations and their characteristics to predict the effect of non-medical mask use in a community setting to prevent laboratory-confirmed SARS-CoV-2 compared to no use of masks. For studies with zero counts in the numerator, we used a continuity correction of 0.001 to estimate the relative risk (RR) in order to ensure inclusion in our analyses; sensitivity analyses considering other continuity corrections produced highly consistent results. To generate summary estimates, we used a custom Bayesian mixed-effects meta-regression tool (MR-BRT, “meta-regression—Bayesian, regularized, trimmed”)^49,50^ which accounted for between-study heterogeneity in the width of the uncertainty interval (see Supplementary Information to Reiner et al.^1^ for more details).

#### Section 1.2. Data Extraction

In total, 63 rows of data from 40 unique publications were extracted (see Figure S1); one article^24^ had an unclear comparison group and is thus missing the ControlGroup variable. Given that it did not have complete data, it was removed from analyses, other than from a sensitivity analysis, resulting in 62 rows of data from 39 unique publications. All publications were traced and translated, when published not in English.

The following variables were of particular interest for the data extraction:

- GeneralPop: Type of population using the mask (general population (1) versus healthcare population (0))
- SE_Asia: Country of study (SE Asian countries (1) vs non-SE Asian countries (0))
- OtherMask: Type of mask (paper / cloth or non-descript masks (1) versus surgical/medical masks and N95 masks (0))
- NoMaskControl: Type of control group (no use (1) versus infrequent use (0))
- Dx: Disease (SARS-CoV 1 or 2 (1) versus H1N1 / influenza/ other respiratory pathogens (0))
- ClinicalDx: Type of diagnosis (clinical (1) versus laboratory (0))
- Study_type: Study type (case control versus clustered randomized controlled trial (RCT) and cohort)

Studies of general population use (13 observations of 63 total) included cluster randomized trials of household member mask use living with an infected individual, cohort analyses of close contacts of infected individuals, and case-control analyses of mask use prior to infection amongst the general population or of secondary infections amongst household members of an infected individual. A total of 19 observations were of non-medical masks, with eight of these specifically assessing use by the general population. The meta-regression suggested the benefits of non-medical masks in the general population to be a 40% (95% uncertainty interval [UI] 20 to 54) reduction in transmission. The benefits of wearing surgical/ medical/N95 masks in the general population were slightly larger, a 43% reduction (95% UI 23 to 59) in transmission. Even larger reductions in transmission were estimated for non-medical (54% [95% UI 40 to 64] and surgical/medical/N95 (56% [95% UI 48 to 64]) mask use amongst healthcare worker populations.

We extracted all relevant data to form 2x2 tables and calculated the RRs and corresponding (log-transformed) Standard Errors for use in our MR-BRT analysis.

IHME’s custom meta-regression tool, MR-BRT, is a trimmed, constrained mixed-effects model that easily facilitates formulating and solving common linear and nonlinear mixed effects models. MR-BRT is open source, and its core computational kernel uses the mixed effects package LimeTr (<https://github.com/zhengp0/limetr>) and the spline package XSpline (<https://github.com/zhengp0/xspline>). A technical report of the statistical models and algorithmic features underlying MR-BRT was published previously.^49^

In order to account for between-study heterogeneity, we included a gamma term to reflect this heterogeneity in the uncertainty interval. MR-BRT also relaxes the assumption that there is one “true” point estimate across all studies and instead allows for variation in the point estimate.

#### Section 1.3. MR-BRT Analysis

Following data extraction, we used MR-BRT to do a meta-regression of the data, allowing for between-study heterogeneity using the gamma term and random effects. The following models were constructed:

- Intercept-only model (shown in Figure S2)
- All univariate associations
- Multivariate models
- Sub-analyses for key variables of interest (population and mask type) (Figure S3)

Sensitivity analyses were also performed to test other models and methods, including:

- Model specifications using different continuity corrections (0.5 versus 0.001) (Figure S4)
- Fixed-effects only model (Figure S5)
- Odds ratios versus RRs (Figure S6)
- Included estimates and confidence intervals from 1 study without full data—intercept only (Figure S7)
- Reported confidence interval versus calculated confidence interval for all studies (Figure S8), and
- Results omitting any clustered RCTs (Figure S9)

All presented analyses use the continuity correction of 0.001, present RRs rather than ORs, and include random-effects with the Gamma term. Sensitivity analyses revealed no major differences in results using a continuity correction of 0.001 versus 0.5, nor using RRs versus ORs.

### Section 2. Trends in the proportion of the population that report using a mask by select demographic characteristics of respondents and by country

#### Section 2.1. Overview

We used three main sources of data on self-reported use of masks: the Facebook Global Symptom Survey,^51^ the Facebook US Symptom Survey,^52^ the PREMISE surveys,^53^ and the YouGov COVID-19 Behaviour Tracker surveys.^54^ Between April 23 and October 19, 2020, Facebook surveyed 30.86 million Facebook users from 198 countries and territories using an instrument with multiple items on behaviors related to COVID-19, including mask use. The YouGov surveys covered 29 countries and interviewed around 726,500 individuals between March 1 and October 24, 2020.

For the US, we used data collected through PREMISE and Facebook to inform the trends in mask use. There were 273,424 total PREMISE responses representing all 50 states and the District of Columbia with responses collected between April 21 and October 25, 2020. Facebook symptom surveys in the US started asking about mask use in the beginning of September (data range from September 8 to October 20, 2020) and included 1,813,000 respondents.

The sample sizes were much smaller in PREMISE compared to those from Facebook, and in 20 states, they were too small to derive a reliable time trend, so before Facebook responses in the US were available (before September 8), we used divisional estimates instead of state-specific estimates. Once US Facebook data were incorporated, we compared estimates on mask use from Facebook and PREMISE to other available sources. National level information on mask use is also being collected by YouGov. We found that the Facebook data were consistent with the YouGov data, while PREMISE data suggested a lower level of use. We also found a state-level survey in Hawaii that was conducted by the Public Policy Center of the University of Hawaii. The estimates from the statewide survey suggest a similar level of mask use as seen in Facebook data; both of these sources suggest a higher level of use than what is reported through the PREMISE surveys. We also considered data in Italy, where both PREMISE and Facebook asked users about mask use. In Italy, also, we found that mask use as reported in Facebook surveys was on average 54% higher than in PREMISE surveys.

Following these checks, we decided to incorporate the Facebook data into our model for the US. We used both sources of data: we used PREMISE data to derive a time trend of mask use and Facebook data to derive the level of mask use. On average, respondents in the Facebook survey were about 30% more likely to report always wearing a mask compared to PREMISE respondents. We adjusted the PREMISE data to the level of Facebook data. Given that we used Facebook data for the level of mask use in all locations outside the US, now that data for the US are available from the same source, using them gives our mask use estimates internal consistency across different locations.

From the Facebook Global Symptom surveys, we used the item, “In the last 7 days, how often did you wear a mask when in public?” to which there are the following possible responses: “All of the time; Most of the time; About half of the time; Sometimes; Never; I have not been in public during the last 7 days”. Respondents for “All of the time” were the numerator in our proportion. From the Facebook COVID-19 Symptom Survey (USA only), we used the item, “In the past 5 days, how often did you wear a mask when in public?” to which the responses are: “All of the time; Most of the time; Some of the time; A little of the time; None of the time; I have not been in public the past 5 days”. Respondents for “All of the time” were the numerator in our proportion.

From the PREMISE surveys, we used the following question: “When you leave your home do you typically wear a face mask (SELECT_ONE)” with responses, “Yes, always; Yes, sometimes; No never”. Respondents for “Yes, always” were the numerator in our proportion. From YouGov, we used the following question: “Thinking about the last 7 days, have you worn a face mask outside your home (e.g., when on public transport, going to a supermarket, going to a main road),” with responses, “Always; Frequently; Sometimes; Rarely; Not at all”. Respondents for “Always” were the numerator in our proportion.

Mask use for each location was estimated using a spline-based smoothing process. We allowed locations with larger sample sizes from survey data to be more flexible. Locations in the bottom quartile of sample size were the smoothest and averaged over 10 neighboring data points; locations in the interquartile range of sample size were smoothed with five neighbors, and locations in the upper quartile of sample size were allowed to be very flexible, smoothed over three neighbors We did not attempt to project changes in mask use into the future. To arrive at smooth, flat values at the ends of the observed data, we computed the average of the change in mask use over the three following days (left tail) and three preceding days (right tail). For locations without data on mask use, we used, in order of preference, national level estimates (for subnational locations), regional estimates, and super-regional estimates based on the regional groupings used by the Global Burden of Diseases, Injuries, and Risk Factors Study (GBD).^55^ The only exception was for countries in Oceania, a region where no data were available through any of the three survey platforms. In the GBD hierarchy, these countries are part of the southeast Asia, east Asia, and Oceania super-region; however, due to cultural differences around the routine seasonal use of masks, we assumed that mask use in Oceania is likely to be more similar to mask use in Australia and New Zealand and so the mask use from the Australasia region was used for countries in Oceania.

### Section 3. IHME’s COVID-19 Projections Modeling Framework

The IHME COVID-19 Projections Modeling Framework have been described in detail previously.^1^ In this section, we provide information on the components of the model that are most related to the analysis presented in this manuscript.

#### Section 3.1. Modeling past deaths using random knot combination splines (RKCS)^1^

##### Data and model overview

To derive infections from deaths and the infection fatality rate for use in the transmission model, we first performed a series of spline regressions using MR-BRT (see Section 1.2). We used an MR-BRT functionality that allowed the user to specify a number of potential knot combinations to be randomly generated and ran separate models for each combination, which were then evaluated for performance and combined using those scores to create a weighted composite of the sub-models. We used 40 combinations in each of the subsequently described model stages, which were run separately by location. The estimates obtained from MR-BRT smoothed the trend in reported deaths and leveraged patterns in reported case and hospital admissions data where available, to make short term forecasts of deaths. Deaths and cases by day were available for every location; hospital admissions data were also available for US 39 states. Before merging with deaths for modeling, we accounted for the lag between hospital admissions or reporting of cases and death based on the Global Line List (<https://github.com/beoutbreakprepared/nCoV2019>)^56^ by shifting dates for these measures forward in time eight days.

##### Deaths as a function of reported cases and hospitalizations

In the first stage, we modeled the cumulative death rate with either the cumulative case rate or the cumulative hospital admission rate as the independent variable. Where data for both of these variables were available, a separate model was run for each. We used a cubic spline with one knot per 12 data points, but with the rightmost interval forced to be linear rather than cubic. We also fixed the rightmost interior knot such that the right segment contained four data points, and we constrained the curve such that cumulative deaths monotonically increased along with cumulative cases/hospitalizations. Because of the shift window, we had eight days of case and hospitalization data that extended past the last day of death data used to fit the model—by linearly extrapolating the tail of the fitted curve, we produced projections of deaths that corresponded to the additional eight days of case or hospitalization data, in addition to our in-sample fit. These death estimates captured the trend in cases or hospitalizations while effectively accounting for changing case- and hospitalization-fatality ratios due to variation in exogenous factors such as age pattern of cases and testing rates.

##### Fitting final deaths curve with uncertainty using all epidemiological data inputs

Using deaths estimated as a function of cases and hospitalizations from the model described above, in addition to observed deaths, we then fit a second-stage model using cumulative deaths from all three sources with time (in days) as the independent variable. We inflated the standard error of the first-stage death estimates by a factor of two so that they were not as influential as the observed deaths. Once again, we used a cubic spline with a linear right tail, and a constraint to be monotonically increasing over time. We also fixed the rightmost interior knot in such a manner that the linear rightmost segment contained four days of reported deaths—and thus 12 days of estimated deaths from cases and hospitalizations. With the resultant curve, we calculated the robust standard deviation of residuals in log daily death space, which we used to independently sample death rates by day, resulting in uncorrelated time series draws representative of the observed noise in the data. We refit models to each of these log daily deaths time series, giving us smooth estimates of death with uncertainty for the full range of dates with observed deaths and extending out to an 8-day projection. We used the same knots samples as the cumulative model, once again with a linear right tail. In this model, we added Gaussian priors on the third derivative of the cubic segments—a stronger prior N(0, 1e-4) on the left-tail segment, and a “dampening” prior N(0, 0.01) on the remaining interior segments. The first of these permitted non-linear growth early on in the outbreak while controlling for erratic behaviour in cubic splines at the terminus, and the second served to reduce volatility that would suggest implausible fluctuations in transmission in the downstream model. Additionally, if fewer than 21 deaths occurred in the past week, we included a strong prior N(0, 1e-8) on the slope of the rightmost segment, forcing it to be flatter. This mitigated the phenomenon of subtle changes of the linear death rate trend in settings with small numbers of deaths being projected as exponential growth in the non-linear transmission model.

##### Day-of-week ensemble

In addition to stochasticity in the day-to-day reporting of these indicators, there was also bias that could be traced to the day of week on which the report falls—in general, Sunday and Monday tend to be underreported, with compensating over-reporting Tuesday through Saturday. While this was generally true, the day of week pattern varied by state. This means that a model run on data reported on a Monday can tend to over-emphasize or create the illusion of declining trends, while the opposite can be true of models run on Saturday-reported data. To address this, we ran seven models, each using data up to the most recent reporting for a given day of the week—so, for results based on data reported on Monday, 26 October, 2020, we ran a model using data up through Tuesday, 20 October, 2020, and additional models for each day up through 26 October, 2020. The predictions of the linear right tail for each model extended to the most recent day predicted in the final model, 3 November, 2020. We used 142 samples from each of the past days models and 148 from the most recent day, resulting in 1000 draws for each location.

#### Section 3.2. Estimating infections from deaths^1^

Conditioning on the death draws, the infection fatality ratio (IFR), and the age-specific mortality rate (MR) (see Sections 4.1 and 4.2), daily infections were inferred by stratifying all-age deaths into age-specific deaths, using the age-specific IFR to determine the number of infections that would have led to this quantity of age-deaths, and then backshifting the infections in time to account for the lag between infection and deaths.

For each of the $j\in1, \ldots,$1000 cumulative death draws time-series, ${CD}^{j}$, one infection-to-death lag, $l^{j}$ was randomly sampled from a discrete uniform distribution on 17 to 21 days.

For each lowest-level location, $loc$:

1. Daily deaths time-series, $DD^{j}(loc)$, were generated by differencing the cumulative deaths time-series, $CD^{j}(loc).$
2. The mortality probabilities, $MP_{ageBin_{i}}\left( loc \right),$ for an individual in this location belonging to each 5-year age bins, $ageBin_{i}$, was calculated:

$MP_{ageBin_{i}}\left( loc \right)= \frac{MR_{ageBin_{i}}\left( loc \right) \times{Pop}_{ageBin_{i}}\left( loc \right)}{\Sigma_{i}(MR_{ageBin_{i}}\left( loc \right) \times{Pop}_{ageBin_{i}}\left( loc \right)}$,

where ${Pop}_{ageBin_{i}}\left( loc \right)$, is the total population for that $ageBin_{i}$ at $loc$. If this was not available, we resorted to using the parent location’s population.

3. The expected age-specific daily deaths time-series, ${DD}_{ageBin_{i}}^{j}\left( loc \right),$ was calculated by stratifying the all-age deaths using the age-specific mortality probabilities, ${MP}_{ageBin_{i}}\left( loc \right)$:

${DD}_{ageBin_{i}}^{j}\left( loc \right)= {MP}_{ageBin_{i}}\left( loc \right)\times DD^{j}\left( loc \right)$.

4. The expected age-specific daily infections time-series, ${DI}_{ageBin_{i}}^{j}\left( loc \right)$, were calculated from the age-specific IFR and daily deaths:

$${DI}_{ageBin_{i}}^{j}\left( loc \right)= {DD}_{ageBin_{i}}^{j}\left( loc \right)/IFR_{ageBin_{i}}\left( loc \right)$$

5. The date of the infection time-series was taken to be the date of the death time series shifted back by $l^{j}$ days.
6. The all-age daily infection time-series was prepared for the SEIR model by summing the infections across all age groups:

$$DI^{j}\left( loc \right)= {\Sigma_{i}DI}_{ageBin_{i}}^{j}\left( loc \right).$$

This process yielded 1000 draws of daily new infections across all modeled locations.

$$MR_{ageBin_{i}}\left( loc \right)/\Sigma_{i}MR_{ageBin_{i}}\left( loc \right)A_{D}{H:D}_{A_{D}}$$

### Section 4. Intermediate quantity modeling

#### Section 4.1. Mortality rate by age estimation^1^

To determine the age pattern of mortality for each location, we assembled available data from multiple global locations and fit a hierarchical meta-regression model. The dependent variable was logit-transformed deaths divided by population. We employed a cascading spline structure to capture the non-linear effect of age, borrowing information from levels higher in the cascade to inform the shape of the age effect in relatively data-sparse regions. The first stage of the cascade was a model fit on all data, with random intercepts by location. The estimated spline coefficients from this global model were passed as Bayesian priors to the subsequent region-specific models, and the region-specific coefficients were passed as priors to location-specific models. For a given in-sample or out-of-sample location, the model from the most detailed geographical level was used to make predictions. Finally, we divided predictions by the minimum location-specific value to obtain age-specific relative mortality ratios.

#### Section 4.2. Infection fatality ratio^1^

We estimated IFRs using random effects meta-analysis, modeling the dependent variable as logit-transformed deaths divided by infections. To calculate the dependent variable, age-specific observations from seroprevalence studies were multiplied by population to obtain an estimate of infections. For each population represented by a seroprevalence observation, a corresponding estimate of deaths was obtained by splitting all-age deaths into age group-specific deaths based on the population’s age distribution and predicted age pattern of mortality. The model included 24 random intercepts by study and a spline to estimate the non-linear effect of age. The spline method allows for the estimation of a continuous age effect from observations recorded as age groups.

#### Section 4.3. Infection to death duration^1^

To estimate the time from infection to death, we brought together two distinct sources of information: published studies of time from infection to symptoms and individual patient data on time from symptom onset to death. Due to a paucity of data on the time from infection to symptom onset, we used the median time reported from a single source (5.1 days) for the first part of this duration and added it to a distribution for the second derived by pooling data from the Global Line List (<https://github.com/beoutbreakprepared/nCoV2019>); Ohio, USA (<https://coronavirus.ohio.gov/wps/portal/gov/covid-19/dashboards>); Rio de Janeiro State, Brazil (<http://painel.saude.rj.gov.br/monitoramento/covid19.html>); Ceara State, Brazil (<https://indicadores.integrasus.saude.ce.gov.br/indicadores/indicadores-coronavirus/coronavirus-ceara>); and Mexico. This pooled dataset included data on 5,125 individuals, with a median time from onset of symptoms to death of 11 days. Informed by this, we used a uniform distribution over 17 to 21 days of lag between infection and death.

#### Section 4.4. Hospitalizations to death ratio^1^

To determine hospitalization, we used cumulative hospital to cumulative deaths ratios estimated directly from hospitalization and mortality data in the US and Europe up to July 10, 2020. We assembled data on COVID-19 hospitalizations from a number of countries and US states. We analyzed hospitalization to death ratios using random effects meta-analysis. We used the location-specific random effect in the estimate for locations with data. In the absence of data, we used the corresponding pooled effect for other countries.

As the hospitalization to death ratios were for all-ages only, to estimate the age-pattern of the hospitalization-to-death ratio, we used the age distribution of hospitalization to death ($H:D$) in the US to estimate the age-distribution for other countries and states:

$$H:D_{ageBin}\left( \mathrm{loc} \right)= \frac{{H:D}_{ageBin}\left( US \right) *{H:D}_{allAge}(loc)\text{ }}{({H:D}_{ageBin}\left( US \right)*D_{ageBin}\left( loc \right))/D_{allAge}(loc)}$$

### Section 5. COVID-19 SEIR model construction for each location

#### Section 5.1. Overview^1^

The primary model for estimating future infections and deaths is a mechanistic compartmental model. Specifically, the fraction of each location’s population that is susceptible ($S$), infected but not infectious (exposed, $E$), infectious ($I_{1}$, $I_{2}$), and recovered ($R$), forming an SEIR model. Temporal variations in past transmission intensity were captured through the time-varying parameter $\beta(t)$. The association between the time-varying transmission intensity and a number of covariates was assessed in a multivariate mixed effects regression across all locations simultaneously. Each of the covariates was then forecast into the future, with certain covariates forecast multiple times corresponding to unique future scenarios. The forecast covariate values and the fitted regression model were then used to estimate future transmission intensity; the future transmission intensity was then used in the SEIR framework to estimate future infections. Finally, reversing the process that estimated past infections from past deaths, future deaths were estimated from future infections.

#### Section 5.2. Covariates

The following covariates were included in the model: population density measured as the share of the population living in areas with more than 2,500 individuals per square kilometer, the fraction of the population living below 100 meters above sea level, smoking prevalence, particulate matter air pollution (PM_2.5_ population-weighted annual average concentration), mobility measured using cell phone apps, mask use, COVID-19 testing per capita, and pneumonia seasonality. Data sources, standardization, and modeling methods for each of these covariates is described in detail in the supplementary information to our published^1^ SEIR model description for the United States.

#### Section 5.3. SEIR-fit^1^

##### Model formulation

To project the full time-series of deaths and infections to the future, we used a transmission model with the following compartments: susceptible, exposed, infected, and removed (SEIR). In particular, each location’s population was tracked through the following system of differential equations:

$$\frac{dS}{dt}=-\beta\left( t \right)\frac{S\left( I_{1}+I_{2} \right)^{\alpha}}{N}$$

$$\frac{dE}{dt}=\beta\left( t \right)\frac{S\left( I_{1}+I_{2} \right)^{\alpha}}{N}-\sigma E$$

$$\frac{dI_{1}}{dt}=\sigma E- \gamma_{1}I_{1}$$

$$\frac{dI_{2}}{dt}=\gamma_{1}I_{1}-\gamma_{2}I_{2}$$

$$\frac{dR}{dt}=\gamma_{2}I_{2}$$

where $\alpha$ represents a mixing coefficient to account for imperfect mixing within each location, $\sigma$ is the rate at which infected individuals become infectious, $\gamma_{1}$ is the rate at which infectious people transition out of the pre-symptomatic phase, and $\gamma_{2}$ is the rate at which individuals recover. This model does not distinguish between symptomatic and asymptomatic infections but has two infectious compartments ($I_{1}$ and $I_{2}$) to allow for interventions that would avoid focus on those who could not be symptomatic. $I_{1}$ is thus the pre-symptomatic compartment.

##### $R_{c}$ *and the effective reproductive number* $R_{eff}\left( t \right)$

We then derived the time-varying basic reproductive number under control, $R_{c}(t)$, and the time-varying effective reproductive number, $R_{eff}(t)$. For a compartmental model with static coefficients, we calculated the basic reproductive number as the largest singular value of the next generation operator

$$R_{c}=\lambda_{max}\left( FV^{-1} \right)$$

where $F$ is the Jacobian of the vector of appearance rates for compartments that actively possess the virus ($E$, $I_{1}$, and $I_{2}$ in our case), and $V=V^{-}+V^{+}$ is the Jacobian of the vector of transport rates of the individuals between these compartments. Both Jacobians were evaluated at the state of disease-free equilibrium (i.e., when $S=N$). The appearance and transport rate vectors for our SEIR model formulation are:

$$f=\left( \begin{matrix} \beta\frac{S\left( I_{1}+I_{2} \right)^{\alpha}}{N} \\ 0 \\ 0 \end{matrix} \right), v=\left( \begin{matrix} \sigma E \\ \gamma_{1}I_{1}-\sigma E \\ \gamma_{2}I_{2}-\gamma_{1}I_{2} \end{matrix} \right).$$

We could then directly calculate the Jacobians at disease-free equilibrium:

$$F=\left( \begin{matrix} 0 & \alpha\beta\left( I_{1}+I_{2} \right)^{\alpha-1} & \alpha\beta\left( I_{1}+I_{2} \right)^{\alpha-1} \\ 0 & 0 & 0 \\ 0 & 0 & 0 \end{matrix} \right)$$

$$V=\left( \begin{matrix} \sigma& 0 & 0 \\ -\sigma& \gamma_{1} & 0 \\ 0 & -\gamma_{1} & \gamma_{2} \end{matrix} \right)\Rightarrow V^{-1}=\left( \begin{matrix} \frac{1}{\sigma} & 0 & 0 \\ \frac{1}{\gamma_{1}} & \frac{1}{\gamma_{1}} & 0 \\ \frac{1}{\gamma_{2}} & \frac{1}{\gamma_{2}} & \frac{1}{\gamma_{2}} \end{matrix} \right)$$

Thus, the next generation operator was

$FV^{-1}=$ $\left( \begin{matrix} \alpha\beta\left( I_{1}+I_{2} \right)^{\alpha-1}\left( \frac{1}{\gamma_{1}}+\frac{1}{\gamma_{2}} \right) & \alpha\beta\left( I_{1}+I_{2} \right)^{\alpha-1}\left( \frac{1}{\gamma_{1}}+\frac{1}{\gamma_{2}} \right) & \alpha\beta\left( I_{1}+I_{2} \right)^{\alpha-1}\cdot\frac{1}{\gamma_{2}} \\ 0 & 0 & 0 \\ 0 & 0 & 0 \end{matrix} \right)$

which yielded

$$R_{c}= \alpha\beta\left( I_{1}+I_{2} \right)^{\alpha-1}\left( \frac{1}{\gamma_{1}}+\frac{1}{\gamma_{2}} \right)$$

##### Fitting $\beta(t)$

We denoted the new daily infections output from the previous step as:

$$f\left( t \right)\approx\beta\left( t \right)S\left( I_{1}+I_{2} \right)^{\alpha}$$

For each draw, we took the parameters governing the transmission dynamics as constant, other than $\beta\left( t \right)$ (i.e., $\alpha$, $\sigma$, $\gamma_{1}$, and $\gamma_{2}$). These parameter values were drawn from distributions based on existing literature.

With a known $f(t)$, we solved a single simple linear ODE to get $E(t)$:

$$\frac{dE}{dt}=-f\left( t \right)-\sigma E$$

This ODE could be solved in closed form using integrating factors, or numerically. In practice, we used the 4^th^ order Runge-Kutta method (RK-4). However, it was useful to solve it in ‘closed form’ using the integration factor approach. Defining

$$v\left( t \right)=\int\sigma dt=\sigma t,$$

we had the closed form solution

$$E\left( t \right)=\exp\left( -\sigma t \right)\int_{0}^{t} -f\left( \tau\right)\exp\left( \sigma\tau\right)d\tau+C\exp(-\sigma t), C=E(0)$$

Having obtained $E\left( t \right)$, we repeated the process, solving for $I_{1}(t)$ and $I_{2}\left( t \right)$:

$$\frac{dI_{1}}{dt}+\gamma_{1}I_{1}=\sigma E\left( t \right)$$

$$I_{1}\left( t \right)=\exp\left( -\gamma_{1}t \right)\left( \int_{0}^{t} -f\left( \tau\right)\exp\left( \sigma\tau\right)d\tau\right)+I_{1}^{0}\exp\left( -\gamma_{1}t \right)$$

$$=F_{1}\left( t \right)+I_{1}^{0}\exp\left( -\gamma_{1}t \right)$$

$$\frac{dI_{2}}{dt}+\gamma_{2}I_{2}=\gamma_{1}I_{1}\left( t \right)$$

$$I_{2}\left( t \right)=\exp\left( -\gamma_{2}t \right)\left( \int_{0}^{t} \gamma_{1}I_{1}\left( \tau\right)\exp\left( \gamma_{2}\tau\right)d\tau\right)$$

$$=\exp\left( -\gamma_{2}t \right)\left( \int_{0}^{t} \gamma_{1}\left( F_{1}\left( \tau\right)+I_{1}^{0}\exp\left( -\gamma_{1}\tau\right) \right) \exp\left( \gamma_{2}\tau\right)d\tau\right)$$

$$=exp \left( -\gamma_{2}t \right)\left( \int_{0}^{t} \gamma_{1} F_{1}\left( \tau\right)\exp\left( \gamma_{2}\tau\right)d\tau\right)+\frac{I_{1}^{0}}{\gamma_{2}-\gamma_{1}}\left( \exp\left( -\gamma_{1}t \right)-\exp\left( -\gamma_{2}t \right) \right)$$

$$=F_{2}\left( t \right)+\frac{I_{1}^{0}}{\gamma_{2}-\gamma_{1}}\left( \exp\left( -\gamma_{1}t \right)-\exp\left( -\gamma_{2}t \right) \right)$$

where $E(t)$ is known when solving $I_{1}$, and then $I_{1}(t)$ is known when solving for $I_{2}$. While useful for formulation to think of the exact solutions, the integrals still had to be solved numerically. We therefore solved all the differential equations using RK-4. With $f(t)$ in hand, we also obtained $S(t)$ by simple integration and subtraction. Having solved for $S(t)$, $I_{1}(t)$, and $I_{2}(t)$, we then had:

$$\beta\left( t \right)=\frac{Nf(t)}{S\left( t \right)\left( I_{1}\left( t \right)+I_{2}\left( t \right) \right)^{\alpha}}$$

#### Section 5.4. $\beta$ regression^1^

Pneumonia seasonality was constructed as an index using medical certification of cause of death data on pneumonia deaths by week and normalized annually. In locations without 4- or 5-star quality cause of death data,^50^ we used latitude as a predictor of the pattern of pneumonia seasonality. With $\beta_{f}(t)$ fit to the data, we next performed a linear regression using the open source mixed effects solver SLIME (<https://github.com/zhengp0/SLIME>) to determine the strength of the relationship between $\beta_{f}(t)$ and the various covariates. All covariates were assumed to have fixed effects, while the intercept was allowed to vary by location. For location $l$, the regression was calculated as:

$$\ln\left( \beta_{p,l} \right)=\alpha_{0,l}+\boldsymbol{X}_{l}\boldsymbol{\alpha}$$

such that the mean squared error between $\beta_{p,l}$ and $\beta_{f,l}$ (our fit from the previous stage) was minimized by location $l$. $\alpha_{0,l}$ is the random intercept for location $l$, $\boldsymbol{X}_{l}$ is a matrix with a column for each covariate in the regression and a row for each day, and $\boldsymbol{\alpha}$ is the coefficient indicating the strength of the relationship between $\log\beta$ and the covariate. Several coefficients in the model were bounded as described in the Supplementary Information supplied to Reiner et al.,^1^ while others were only constrained by directional bounds. Not all covariates were time varying. These non-time varying covariates were used to explain some of the location specific variance otherwise absorbed into the random intercept. Using the fitted $\alpha$ and the forecasted covariates, we produced, by draw, estimates of future transmission intensity $\beta_{p}(t)$.

#### Section 5.5. $\beta$ adjustments^1^

To ensure continuity from our fitted $\beta_{f}$ from SEIR-fit to the predicted $\beta_{F}$ into the future, we shifted the predicted $\beta_{p}$. Generally speaking, we shifted $\beta_{p}$ towards $\beta_{f}$ by first ensuring that on the day of transition ($T$), $\beta_{F}\left( T \right)=\beta_{f}(T)$. We then calculate the average residual over the past $n$ days and assume that are future predictions should be adjusted by this quantity. From the day of transition to $M$ days in the future we linearly interpolate our adjustment from that on the day of transition to the average residual and then apply the average residual to all days past $M$ days in the future. More specifically, we defined $r(t)$ as

$$r\left( t \right)=\log\left( \frac{\beta_{f}\left( t \right)}{\beta_{p}\left( t \right)} \right), t\leq T$$

and $\beta_{F_{1}}(t)$ and $\beta_{F_{2}}(t)$ as

$$\beta_{F_{1}}\left( t \right)=\exp\left( r\left( T \right) \right)\beta_{p}\left( t \right), t\geq T$$

$$\beta_{F_{2}}\left( t \right)=\exp\left( \frac{1}{n}\sum_{i=1}^{n} r\left( T-i+1 \right) \right)\beta_{p}\left( t \right), t\geq T$$

and transition weights $w(t)$ as

$$w\left( t \right)=\left\{ \begin{matrix} \frac{M-\left( t-T \right)}{M}, T\leq t\leq T+M \\ 0, otherwise \end{matrix} \right.$$

Then, for a given $n$ and $M$, we defined $\beta_{F}(t)$ as

$$\beta_{F}\left( t \right)=w\left( t \right)\beta_{F_{1}}\left( t \right)+\left( 1-w\left( t \right) \right)\beta_{F_{2}}(t)$$

Based on out-of-sample tests similar to the sensitivity analyses for optimal values of $M$ and $n$ described in the Supplementary Information for Reiner at al.,^1^ we found that the optimal $n$ was 42 and the optimal $M$ was for $M$ to be, by draw, drawn from a uniform distribution of windows from 7 to 28 days.

#### Section 5.6. SEIR-predict^1^

The general format of our predictions was relatively simple: we took the final predicted $\beta_{F}$ and ran our system of ODEs forward in time using our fitted compartment values at time $T$ as the initial conditions of the second SEIR model.

There were, however, a number of simplifications made within our modeling formulation. First, we ignored the potential for importation, which may be more likely in larger, denser locations. Second, we assumed a well-mixed population, which may be more egregious in smaller, less dense locations. As two intermediate solutions for this, we introduced two correction factors. In each location, we only used one or the other correction factor, and the use and magnitude of the correction was based on OOS predictive validity, by dropping eight weeks of data and comparing the predicted outbreak to the observed one. The first correction factor allowed for the addition of a small number of additional infections above and beyond those from the interaction between $I_{1}$ and $I_{2}$ and $S$. These can be envisaged as individuals traveling outside the location, becoming infected, and returning as exposed individuals. The second correction factor removed a small fraction of exposed individuals from the $E$ compartment and moved them directly to the recovered compartment. Our model acted on the fraction of individuals who were infectious, exposed, etc., and the results of allowing for fractional infectious individuals (and no possibility for truly ‘zero’ infections) could alter the dynamics for small locations. These corrections can be mathematically described using $\theta^{+}$ and $\theta^{-}$ for the importation correction and the small location correction, respectively. Again, each location received only one of these, and they altered the SEIR model formulation for prediction as

$$\frac{dS}{dt}=-\beta\left( t \right)\frac{S\left( I_{1}+I_{2} \right)^{\alpha}}{N}-\theta^{+}S$$

$$\frac{dE}{dt}=\beta\left( t \right)\frac{S\left( I_{1}+I_{2} \right)^{\alpha}}{N}-\sigma E+\theta^{+}S-\theta^{-}E$$

$$\frac{dI_{1}}{dt}=\sigma E- \gamma_{1}I_{1}$$

$$\frac{dI_{2}}{dt}=\gamma_{1}I_{1}-\gamma_{2}I_{2}$$

$$\frac{dR}{dt}=\gamma_{2}I_{2}+\theta^{-}E$$

With these correction factors identified, we could then run our ODEs forward (again using the RK-4 algorithm), to have a complete time-series of infections through the end of the year.

### Section 6. Final data combination and summarization

The transmission model produced 1,000 full time series (including projections) of COVID-19 cases and deaths. We summarized draws into means with 95% UIs for reporting. To control for extreme values, the top 2.5% and bottom 2.5% of draws were dropped and replaced through random resampling of the remaining 950 draws.

#### Section 6.1. Scenarios

We estimated the trajectory of the epidemic by location (countries and subnational locations) under a “mandates easing” scenario that models what would happen in each location if the current pattern of lifting social distancing mandates continues and new mandates are not imposed.

To model a more plausible scenario, we predicted likely responses of national and local governments (in subnational locations) during the second phase of the pandemic using observed data from the first phase. For this plausible reference scenario, the model assumes that in each location, an easing social distancing measures (SDMs) will continue on the same trajectory until the daily death rate reaches a threshold of 8 deaths per million. If the daily death rate in a location exceeds that threshold at any point in the forecasted study period, we assume that there will be a reintroduction of SDMs for a six-week period. The chosen threshold (of a rate of daily deaths of 8 per million) represents the 90^th^ percentile of the distribution of the daily death rate at which locations around the world implemented their mandates during the first months of the COVID-19 pandemic. Because of the poor economic impacts of the first set of SDMs, we anticipated increased reluctance from governments to re-impose mandates. To reflect this reluctance, we selected the 90^th^ percentile rather than the 50^th^ percentile. In locations that we do not forecast to exceed the threshold of a daily death rate of 8 per million during the forecasted study period, the projection is based on the covariates in the model and the forecasts for these covariates to March 1, 2021. In this scenario, when the daily projected death rate exceeds 8 per million, we assume that mandates will be introduced within seven days, reducing mobility to either the location-specific sum of the effects of all social distancing mandates or to the lowest observed level, whichever is greater.

The scenario of universal mask wearing models what would happen if 95% of the population in each location always wore a mask when they were in public. This value was chosen to represent the highest observed rate of mask use observed globally during the COVID-19 pandemic through July 2020. In this scenario, we also assume that if the daily death rate in a location exceeds 8 deaths per million, SDMs will be reintroduced for a six-week period.

### Section 7. Limitations of the analysis

First, there is a set of limitations related to the meta-regression, including the following. The number of published studies on the protection provided by cloth masks worn by the general public is limited. With rapid development of the COVID-19 literature, new data on the effectiveness of masks can quickly be incorporated into our meta-regression model. Future studies could change our pooled estimate of the effect size and/or the large uncertainty interval around it in the meta-regression. As we looked at multiple observations per study, it was not really feasible to account for all possible clustering. In our previous research, we performed sensitivity analyses (found in the supplementary information in Reiner et al.,^1^ and found minimal differences in investigating the role of clustering in our results. Further, the studies in our meta-regression had different endpoints and while we controlled for that, it would be ideal to have more studies that focus on COVID-19 as an endpoint.

Second, related to the modeling framework, we use an SEIR model to predict the course of the epidemic with and without universal mask use. In general, SEIR models have tended to overestimate the infections and deaths associated with COVID-19 in their forecasts. Over-estimation is likely due to the fact that individuals change their behaviors as the epidemic gets worse around them and governments tend to react when hospital systems are nearing capacity. We built the government response into our reference scenario and tried to use empirically observed data on mobility and current mask use to reflect individual behavioral responses. Further, our model makes a number of simplifying assumptions associated with mixing and transmission heterogeneity and as such, our conclusions must be considered with these assumptions in mind.^1^ Also, we used a log-linear mixed regression, which does not take into account the potential for non-linear relationships. We acknowledge that there are likely non-linear relationships between some of the drivers of transmission and transmission intensity. Moreover, we expect there to also be complex lagged relationships between covariates and transmission (e.g., fatigue related to duration of mandate altering its impact on transmission). Improving how our model uses covariates to capture temporal variation in new infections is an open avenue of research.

Third, our out-of-sample predictive validity testing has shown that errors tend to progressively get larger the longer the forecast but has also shown that errors are much larger in settings where there are fewer than 50 total deaths to date. For example, predictions from publicly available models for sub-Saharan Africa have been particularly inaccurate.^57^

Fourth, we assume that cases and deaths used in our analysis are accurately reported by Johns Hopkins University or location-specific Ministries of Health except in Ecuador and Peru, where excess mortality analyses have indicated substantial underreporting of COVID-19 deaths. Other countries may not be detecting or reporting deaths and cases due to lack of testing or other considerations. Overall, though, we believe that our prediction model performs well and where antibody tests have been conducted at the population level, we have found our model based on deaths has matched these results.^1^

Fifth, our models are sensitive to the trends in the last 7–14 days in deaths and somewhat sensitive to the trend in cases. In settings where deaths and cases are steadily rising, the model will tend to have large estimates of β_t._ If the rise in transmission is not captured by trends in mobility or other covariates, the unexplained residual in the model increases and this is then reflected in the forecasts by day through to January 1, 2021. The reverse relationship also holds true for when there is a consistent downward trend. The sensitivity of our model to data trends is a strength in that it makes our models reflect the on-the-ground realities; it is also a challenge in the sense that our model results will change when there are changes in recent transmission that are not captured by the covariates.

Sixth, we rely on self-reported data that is collected via mobile phone app-based surveys on use of masks. In addition to the usual biases that accompany self-reported data, in this case we do not know whether respondents may be reporting their behavior differently in settings with mask mandates in place compared to settings without mask mandates. The respondents to app-based surveys are also likely not to be a truly representative sample of the populations in each location. The degree to which the respondents represent the general population varies across locations and depends on the prevalence of Facebook use and other app use in each country. While this is a limitation of the data that are currently available, we believe that given the samples tend to be biased towards more educated, urban, and younger populations, the reported mask use is likely to be an over-estimate in these locations. If this is true, then the estimates of the impact of expanding mask use to reach 95% coverage in these populations would be an underestimate of the true potential effect of the intervention.

### Section 8. Figures and tables

#### Figure S1. Relative risk of respiratory virus infection by publication first author and year of publication

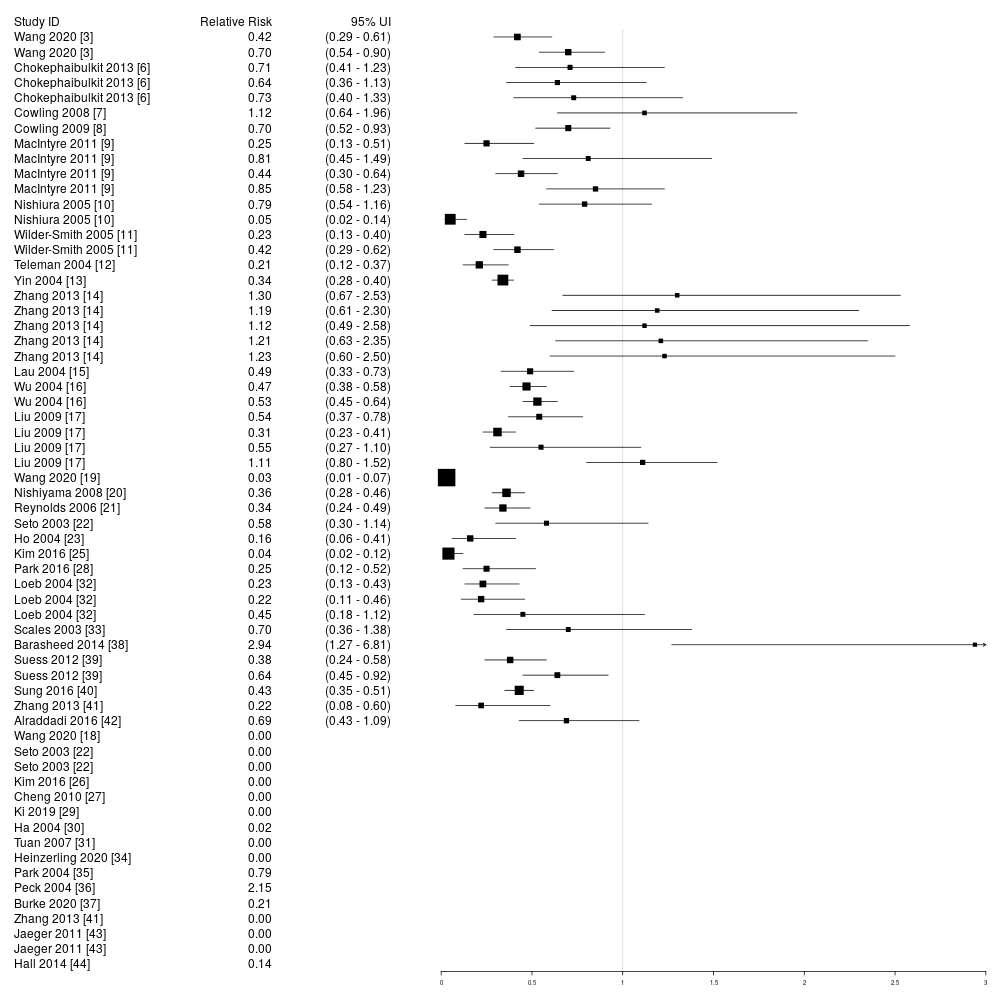

This figure demonstrates the relative risk (RR) of respiratory virus infection, recalculated for use in this meta-regression based on numbers reported in the publication. We present a RR of zero where there were zero counts in the numerator; confidence intervals were not presented for these studies in the figure. These studies were included in the analysis with a continuity correction.

#### Figure S2. Univariate MR-BRT model results

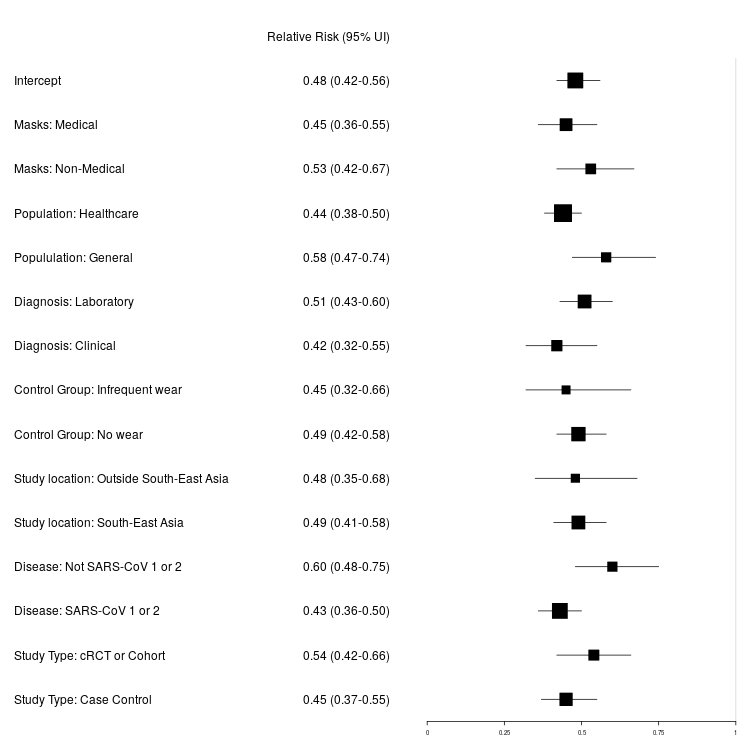

This figure depicts the relative risk (and corresponding 95% uncertainty interval [UI]) for each variable of interest, separately, using meta-regression techniques; the first row presents the results of an intercept-only model. The size of the box is proportional to the precision of the estimate, based on number of observations, with more precise studies having larger boxes. Medical mask=medical, surgical, or N95 masks.

#### Figure S3. Sub analyses: (A) estimates for non-medical mask usage among those in a healthcare and non-healthcare setting; (B) estimates for general population non-medical and medical mask usage in non-healthcare settings

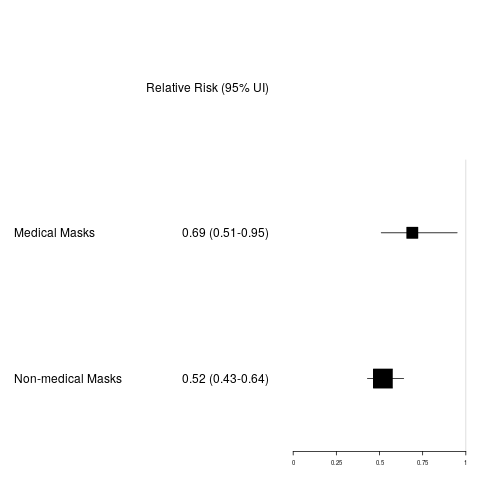

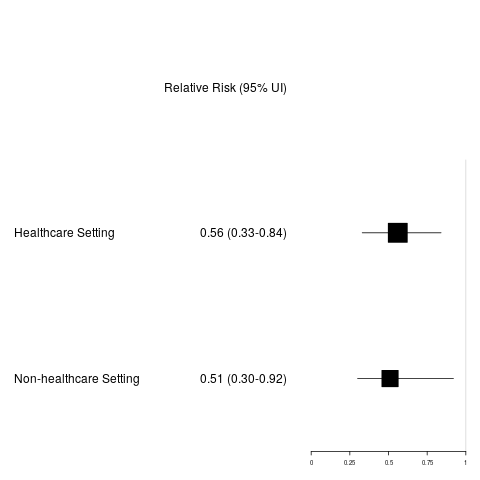
**A**	**B**

This figure depicts the relative risk (and corresponding 95% uncertainty interval [UI]) for each variable of interest, separately, using meta-regression techniques. The size of the box is proportional to the precision of the estimate, based on number of observations, with more precise studies having larger boxes. Medical mask=medical, surgical, or N95 masks.

#### Figure S4. Sensitivity analysis using a continuity correction of 0.5

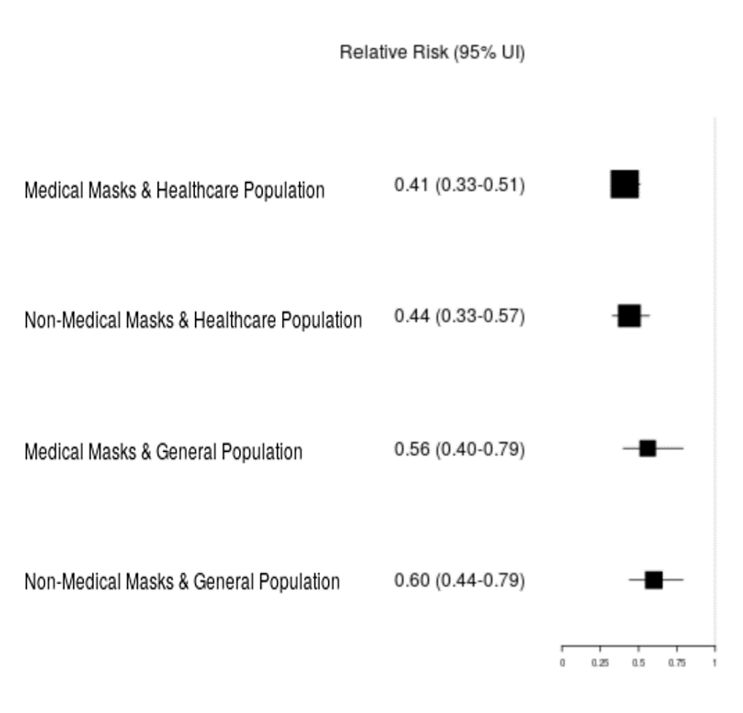

The continuity correction of 0.5 is in contrast to the main analysis, which used a continuity correction of 0.01. This figure depicts the relative risk (and corresponding 95% uncertainty interval [UI]) for each variable of interest, separately, using meta-regression techniques. The size of the box is proportional to the precision of the estimate, based on number of observations, with more precise studies having larger boxes. Medical mask=medical, surgical, or N95 masks.

#### Figure S5. Sensitivity analysis using a fixed-effects model

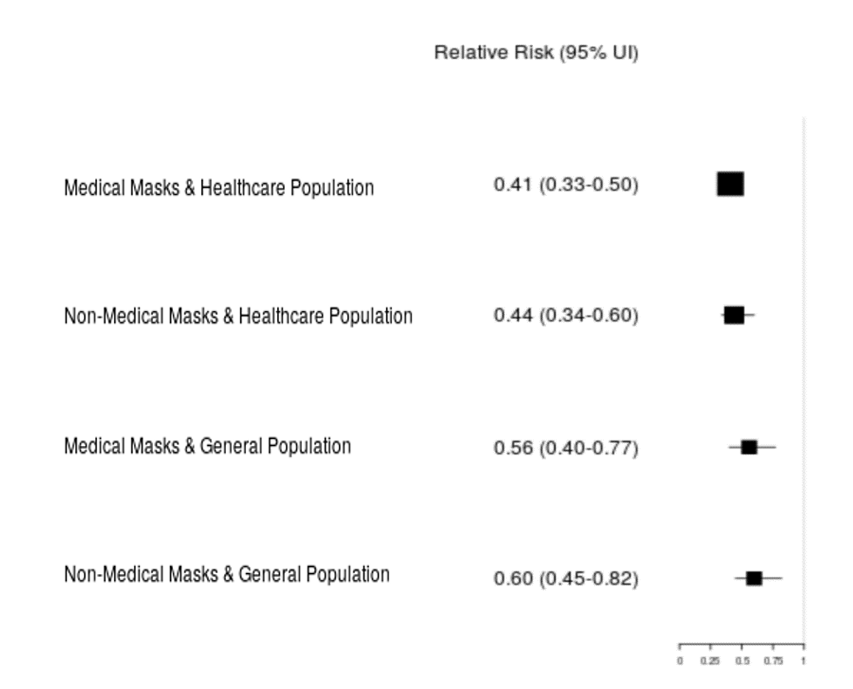

The use of a fixed-effects model is in contrast to the main analysis, which used a random-effects model. This figure depicts the relative risk (and corresponding 95% uncertainty interval [UI]) for each variable of interest, separately, using meta-regression techniques. The size of the box is proportional to the precision of the estimate, based on number of observations, with more precise studies having larger boxes. Medical mask=medical, surgical, or N95 masks.

#### Figure S6. Sensitivity analysis using calculated odds ratios

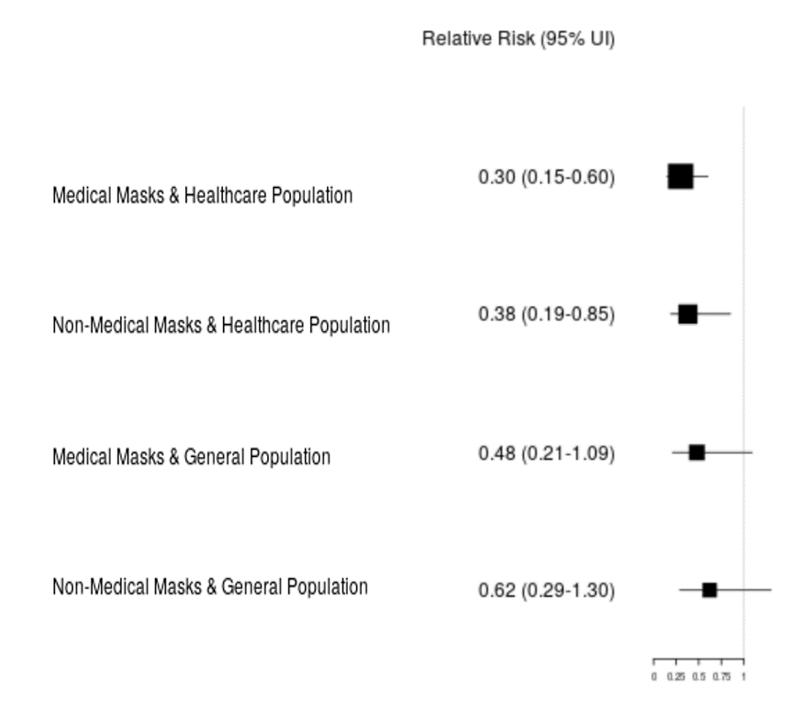

The use of calculated odds ratios is in contrast to the main analysis, which used relative risks. This figure depicts the relative risk (and corresponding 95% uncertainty interval [UI]) for each variable of interest, separately, using meta-regression techniques. The size of the box is proportional to the precision of the estimate, based on number of observations, with more precise studies having larger boxes. Medical mask=medical, surgical, or N95 masks.

#### Figure S7. Sensitivity analysis looking at an intercept-only model with all 63 observations versus only those with full data (n=62)

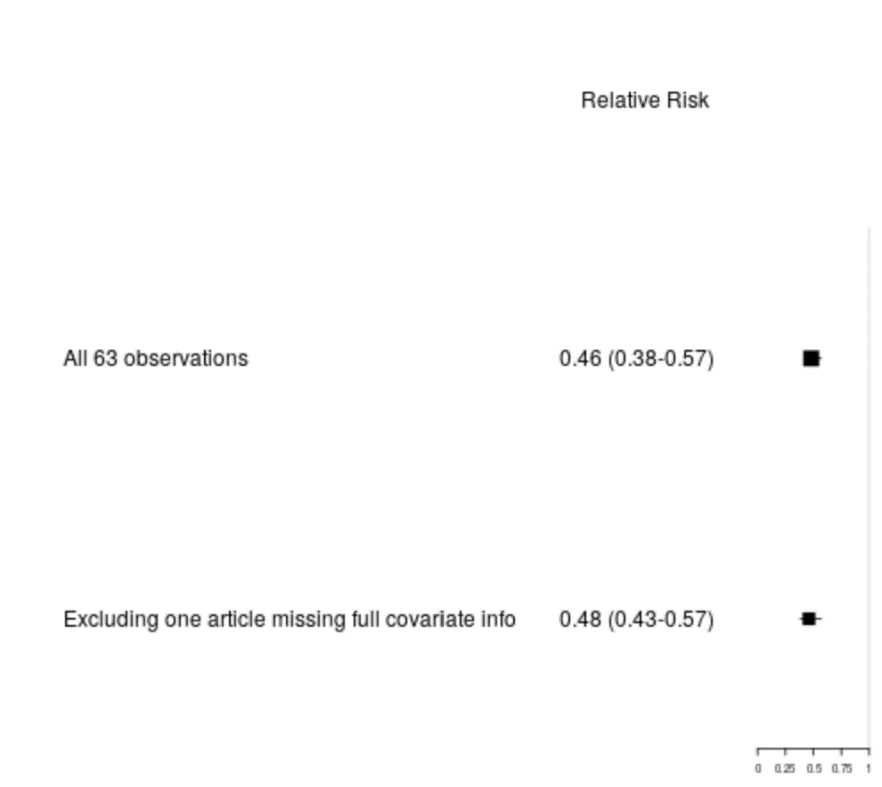

This figure depicts the relative risk (and corresponding 95% uncertainty interval [UI]) for each variable of interest, separately, using meta-regression techniques. The size of the box is proportional to the precision of the estimate, based on number of observations, with more precise studies having larger boxes. The “all 63 observations” variable includes estimates from one study without full data, which was excluded from all other analyses. The “excluding one article” variable excludes the estimates from this study with incomplete data.

#### Figure S8. Sensitivity analysis using reported confidence interval, where available, versus calculated standard error for all 63 observations

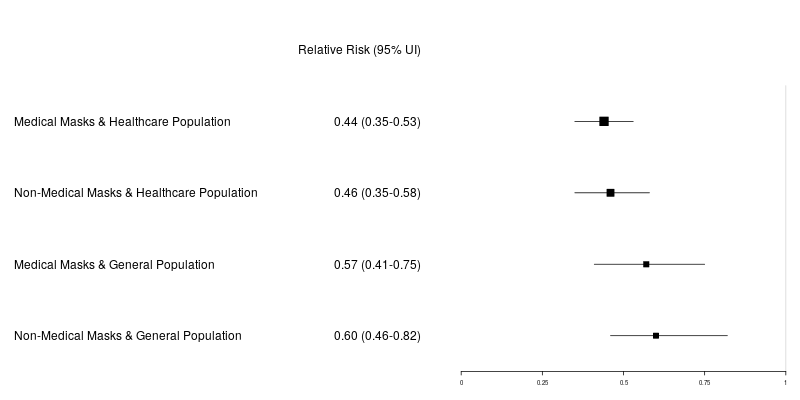

The use of reported confidence intervals, where available is in contrast to the main analysis, which used calculated standard errors. This figure depicts the relative risk (and corresponding 95% uncertainty interval [UI]) for each variable of interest, separately, using meta-regression techniques. The size of the box is proportional to the precision of the estimate, based on number of observations, with more precise studies having larger boxes. Medical mask=medical, surgical, or N95 masks.

#### Figure S9. Sensitivity analysis omitting cRCTs (n=53 observations)

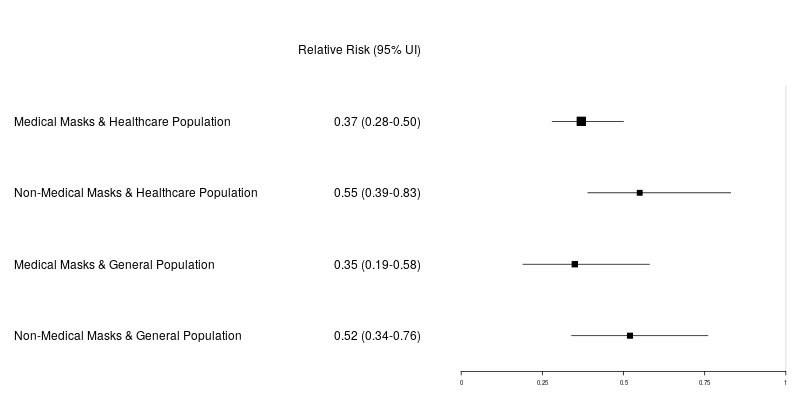

This figure depicts the relative risk (and corresponding 95% uncertainty interval [UI]) for each variable of interest, separately, using meta-regression techniques. The size of the box is proportional to the precision of the estimate, based on number of observations, with more precise studies having larger boxes. cRCTs=cluster randomized controlled trials. Medical mask=medical, surgical, or N95 masks.

Figure S10. Mask use by gender, globally and by Global Burden of Disease (GBD) super-region, 2020
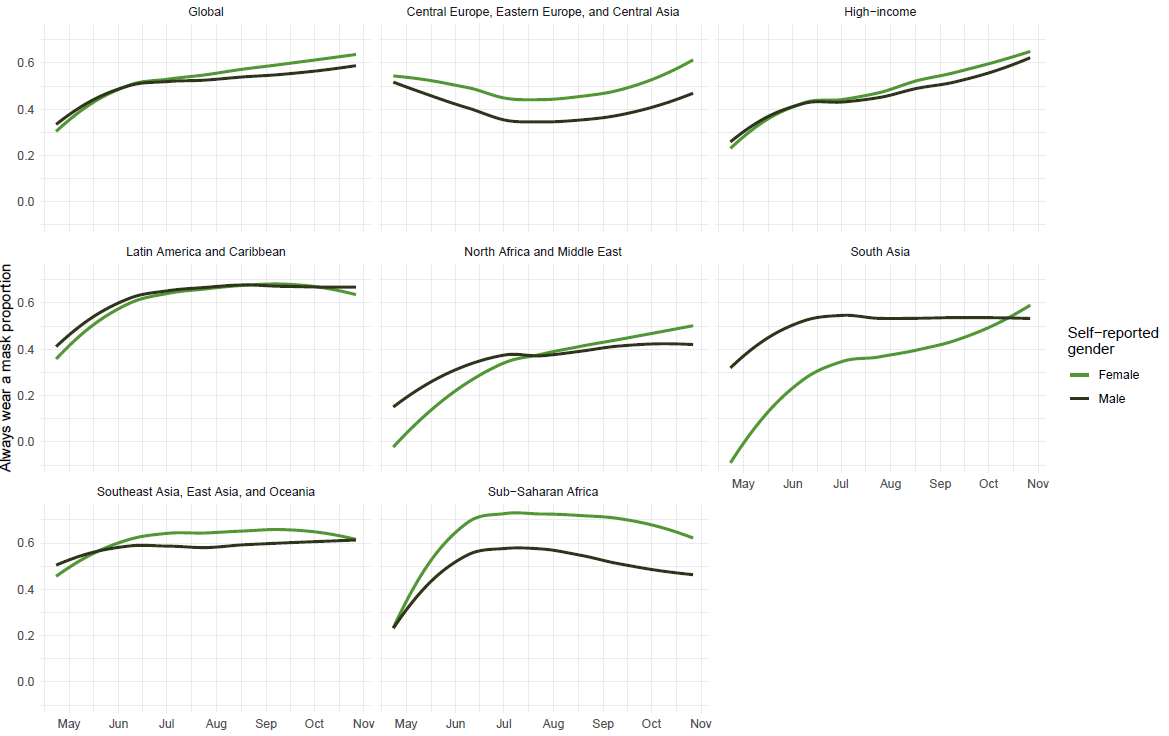

Figure S11. Mask use by adult age group globally and by GBD super-region, 2020
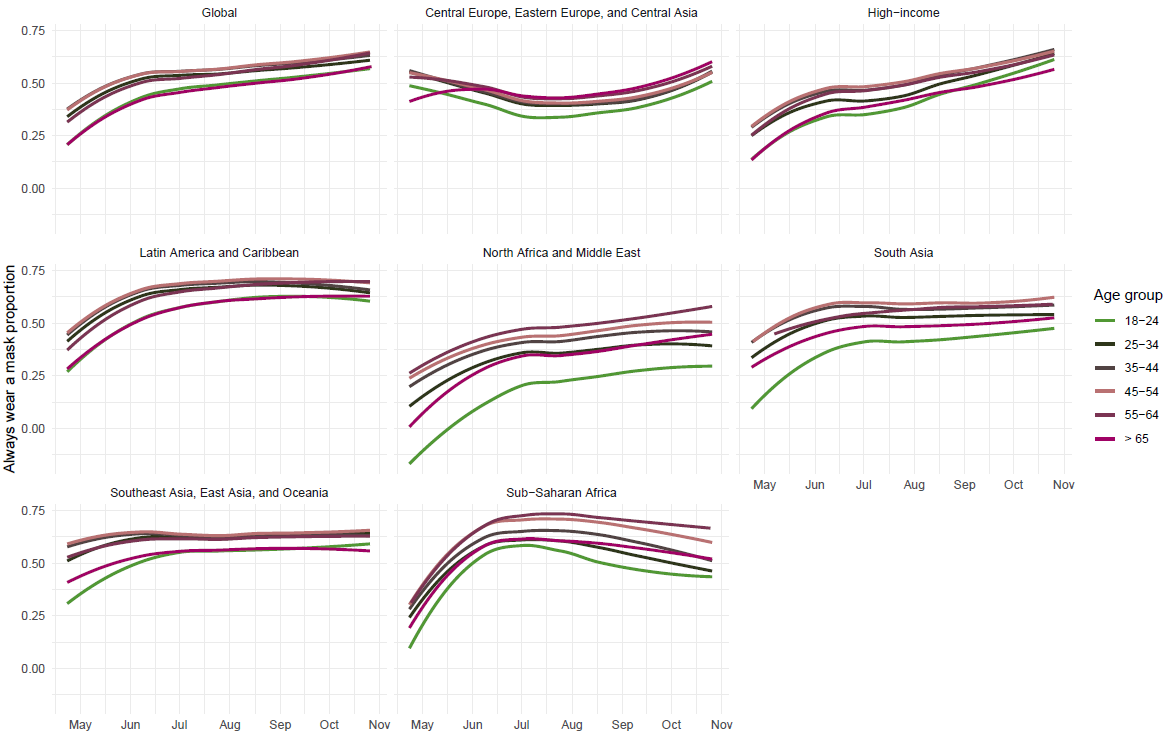

Figure S12. Mask use by urbanicity globally and by GBD super-region, 2020
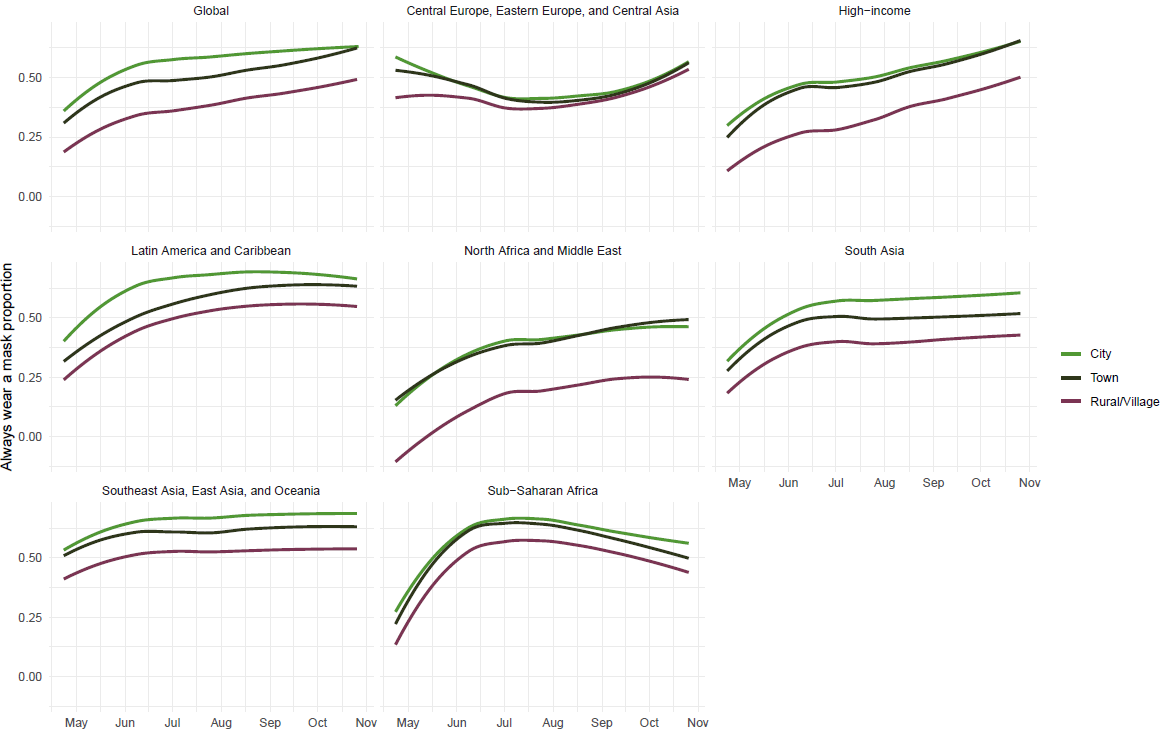

Figure S13. Mask use by self-reported number of contacts in the previous 24 hours globally and by GBD super-region, 2020
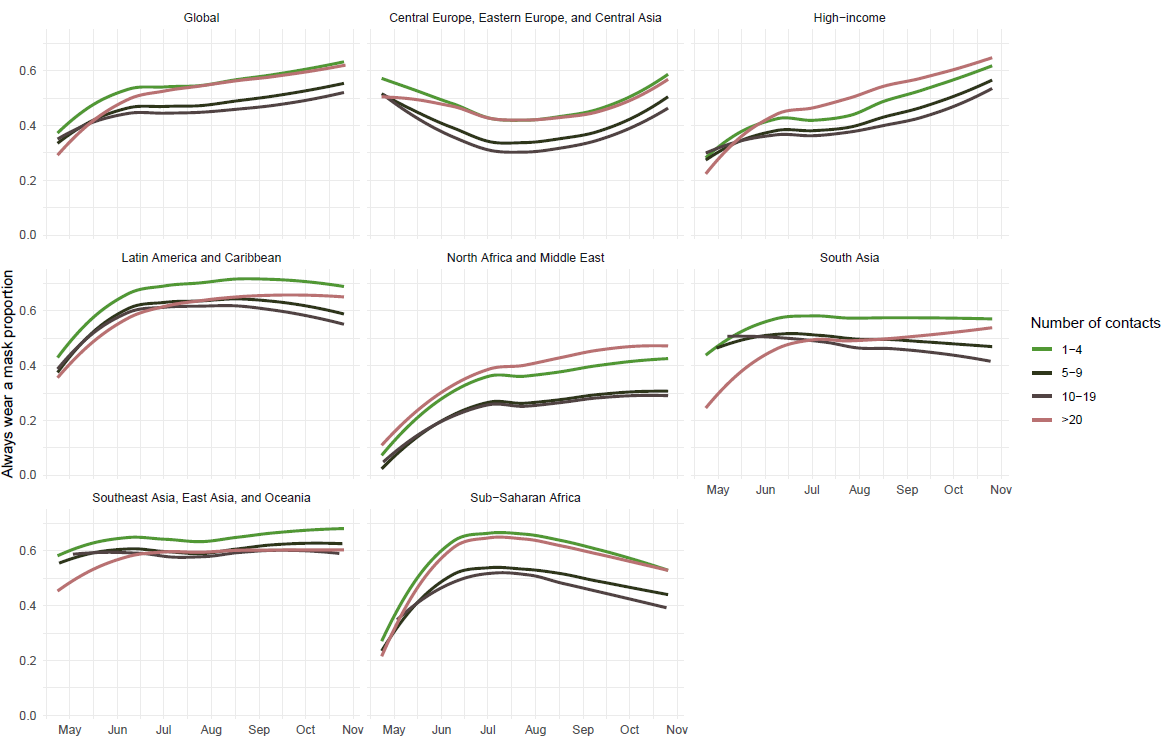

#### Figure S14. Estimates of mask use at the national level, organized by GBD region, and for countries with sub-national units, between January 1, 2020 and January 1, 2021

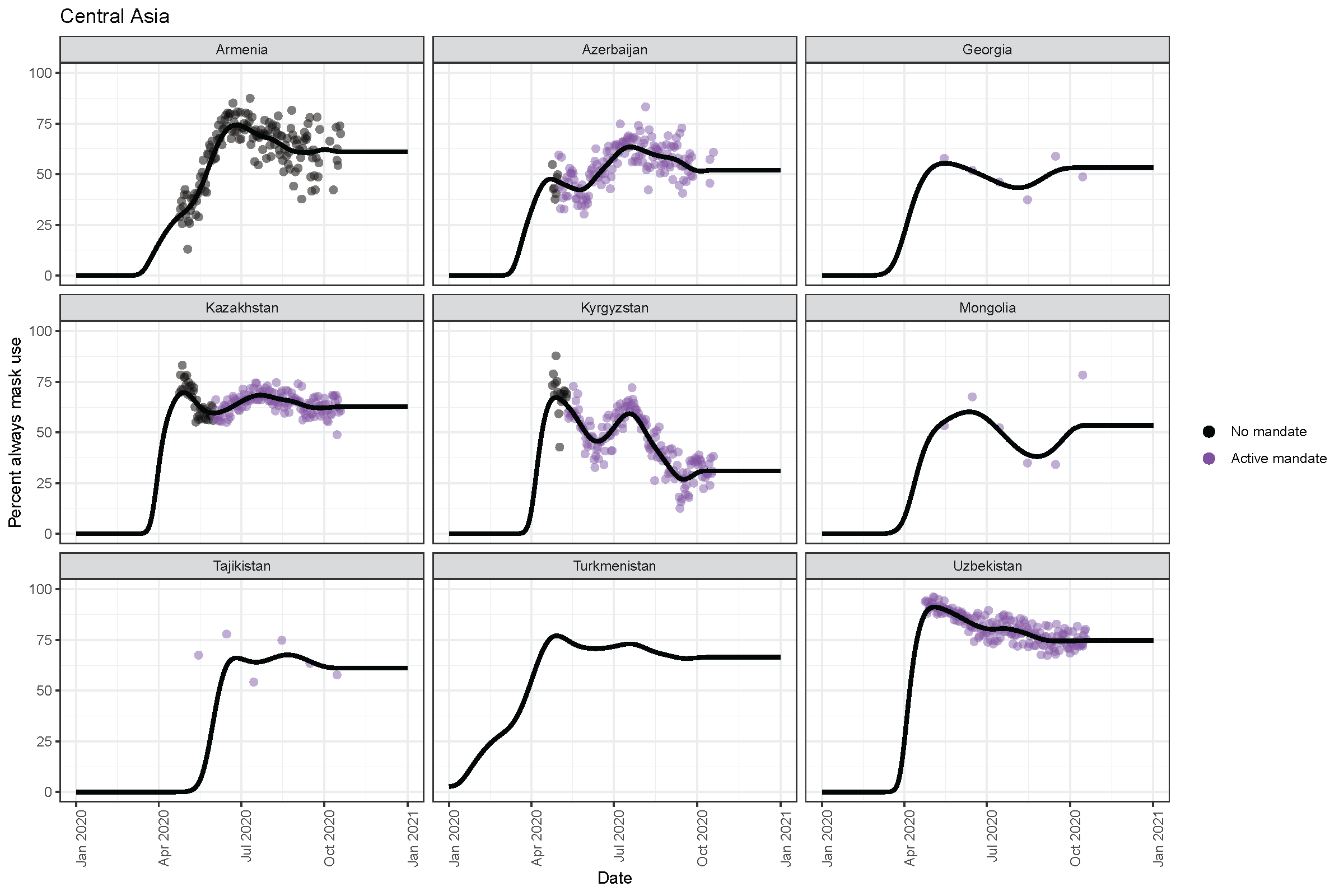

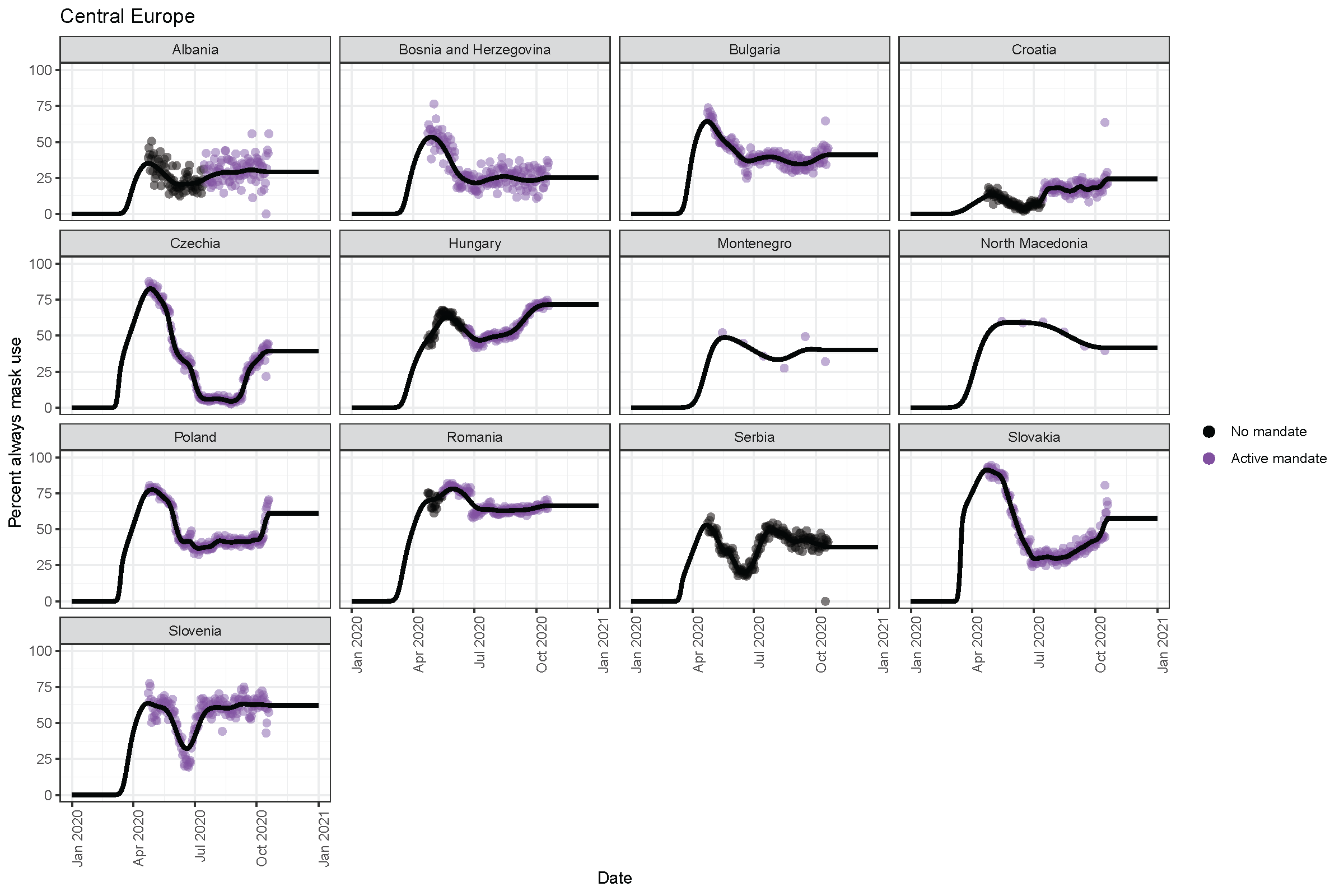

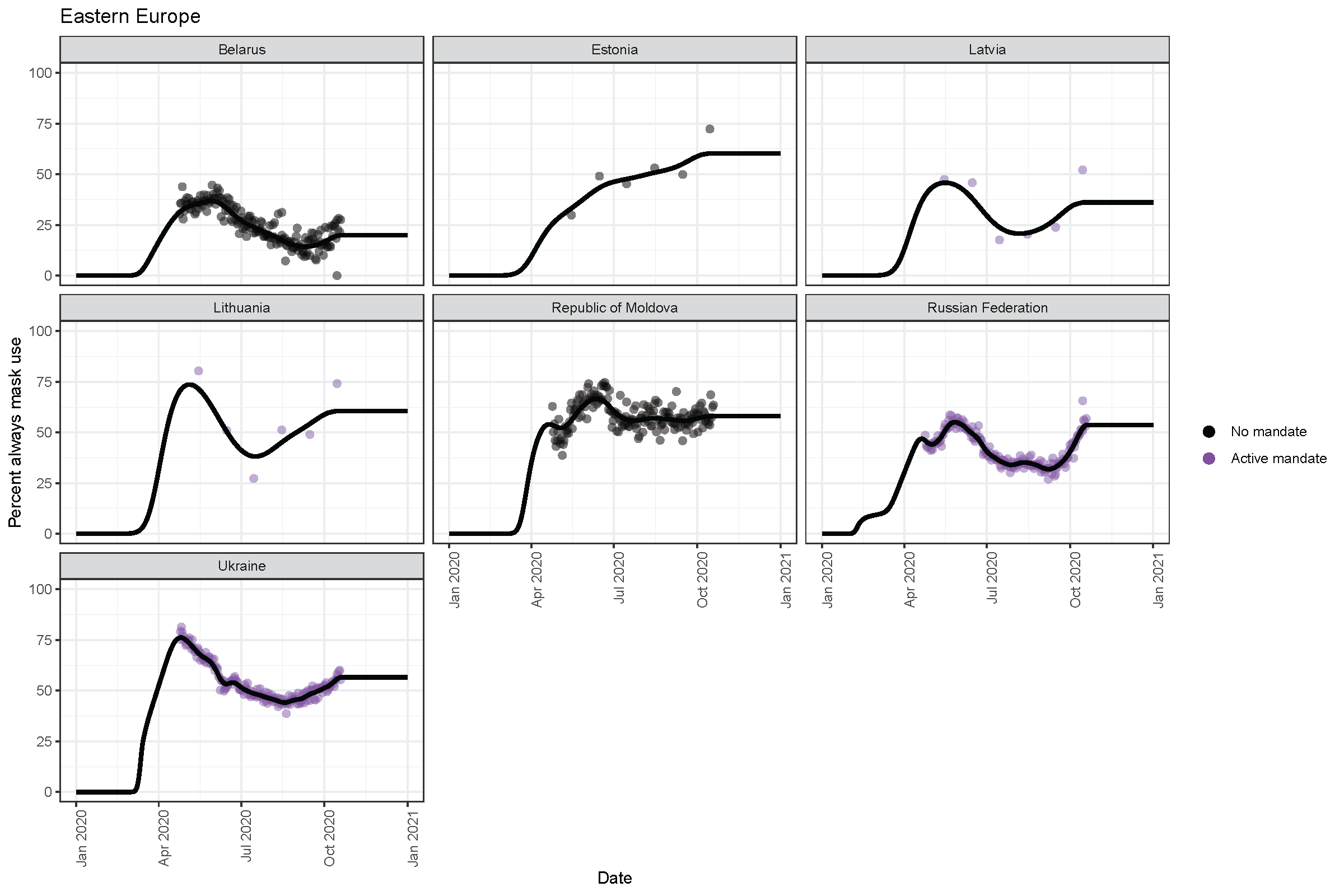

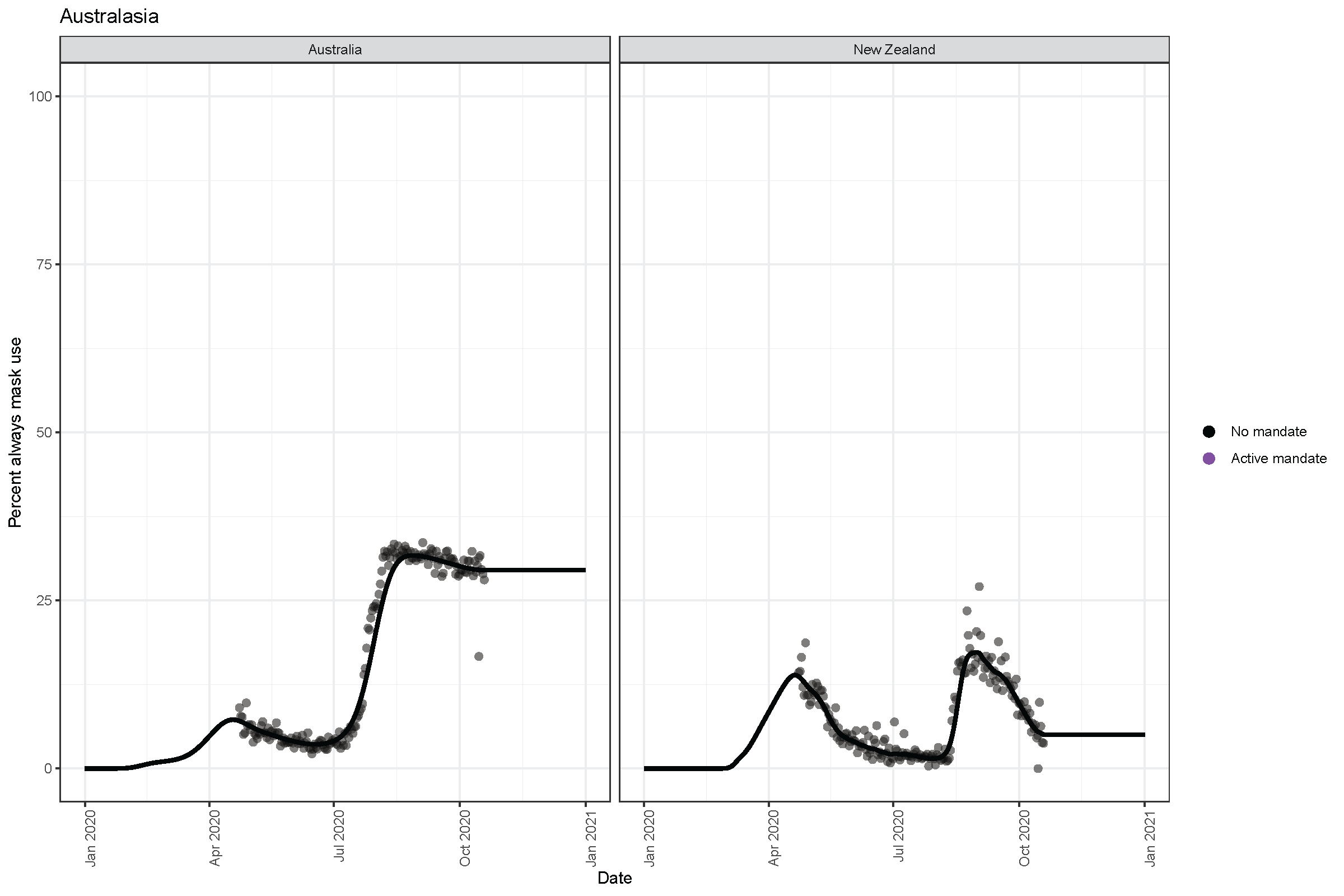

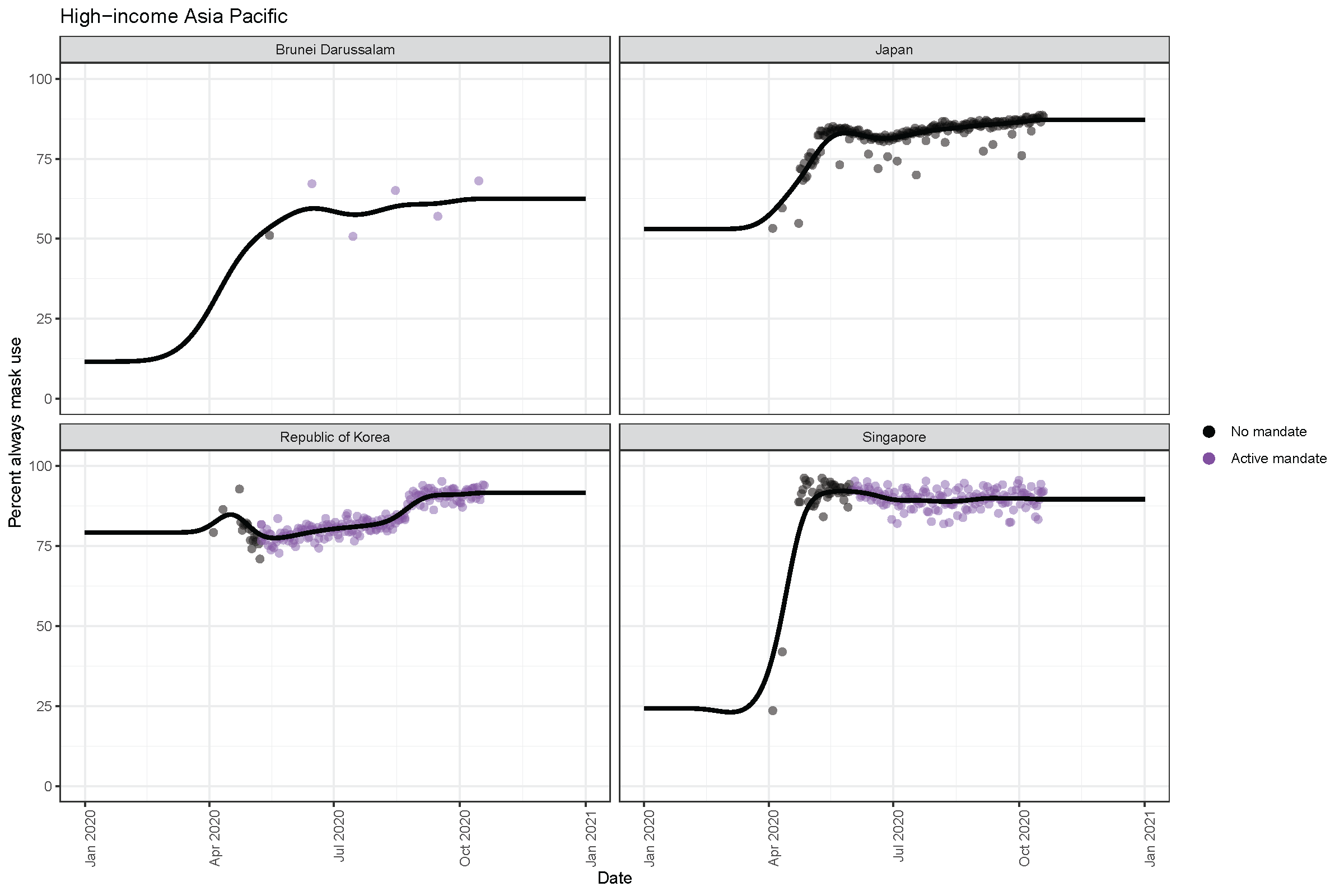

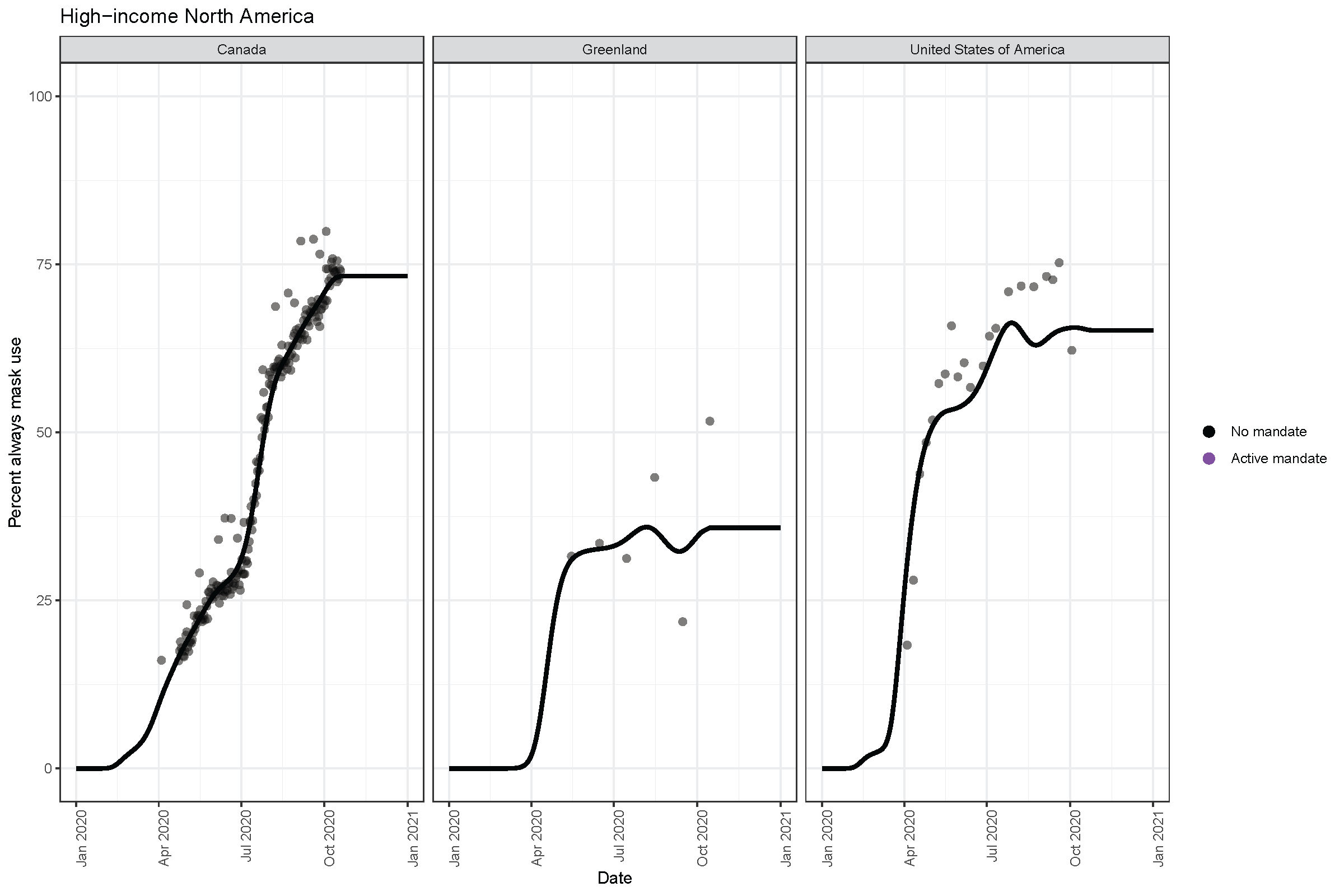

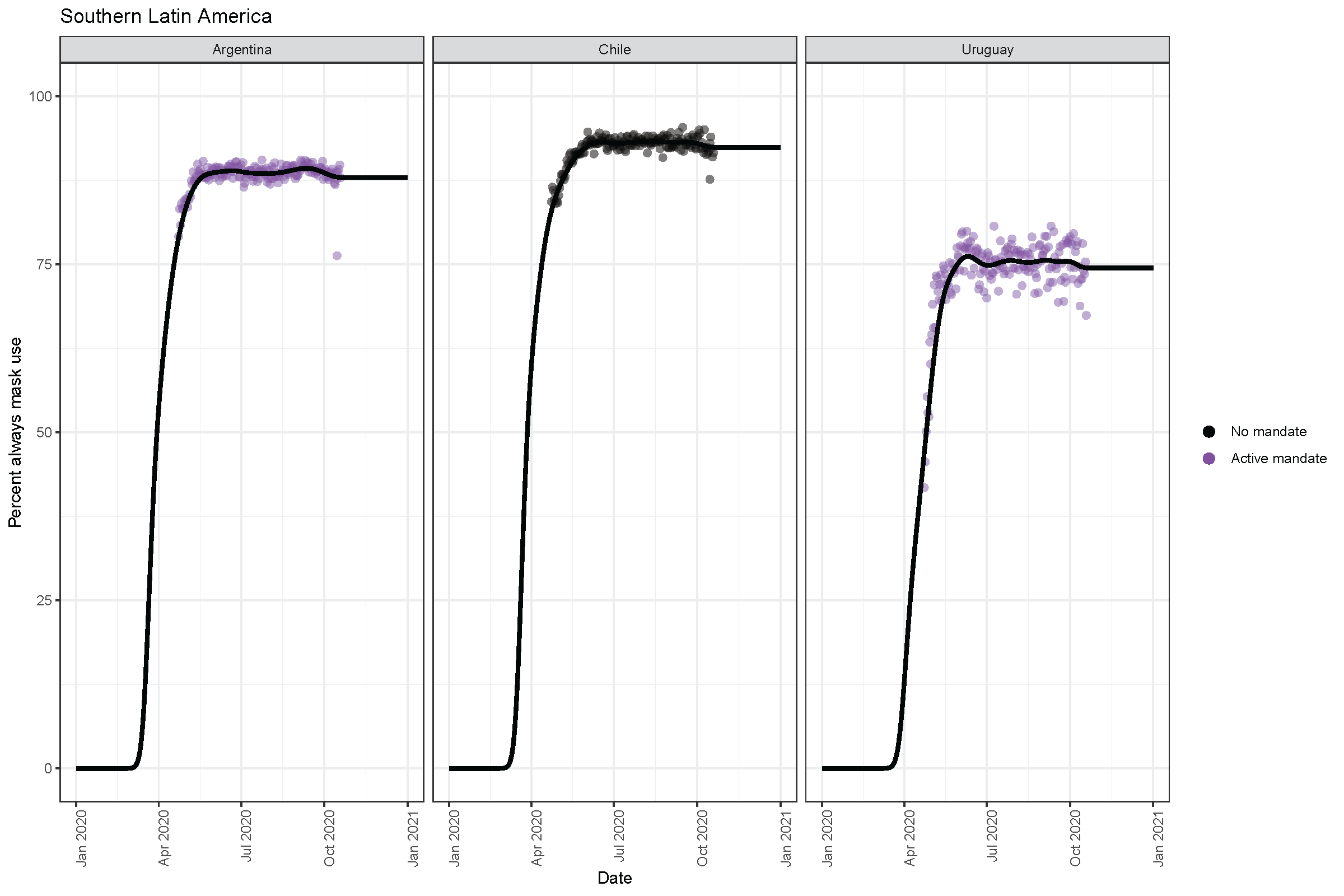

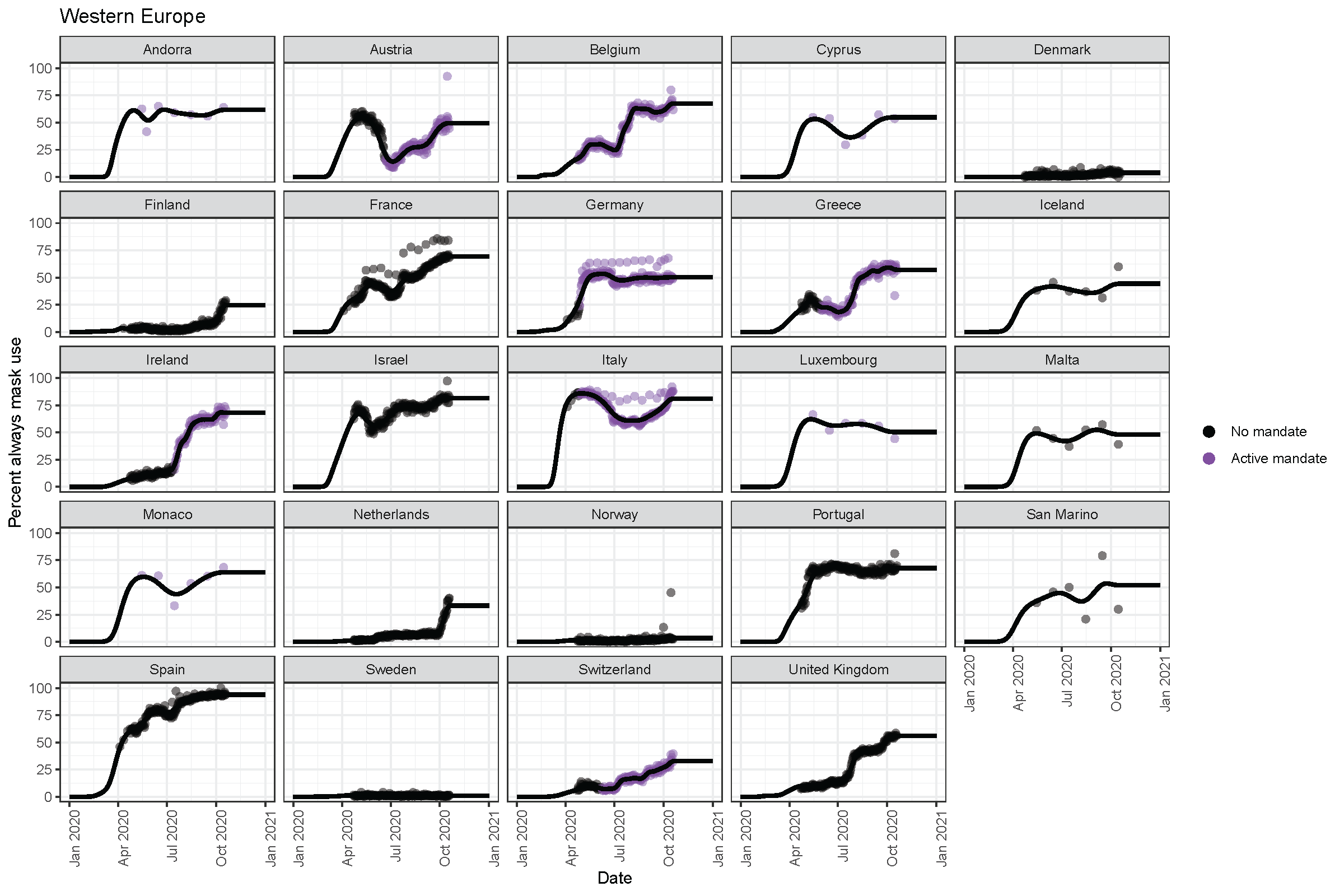

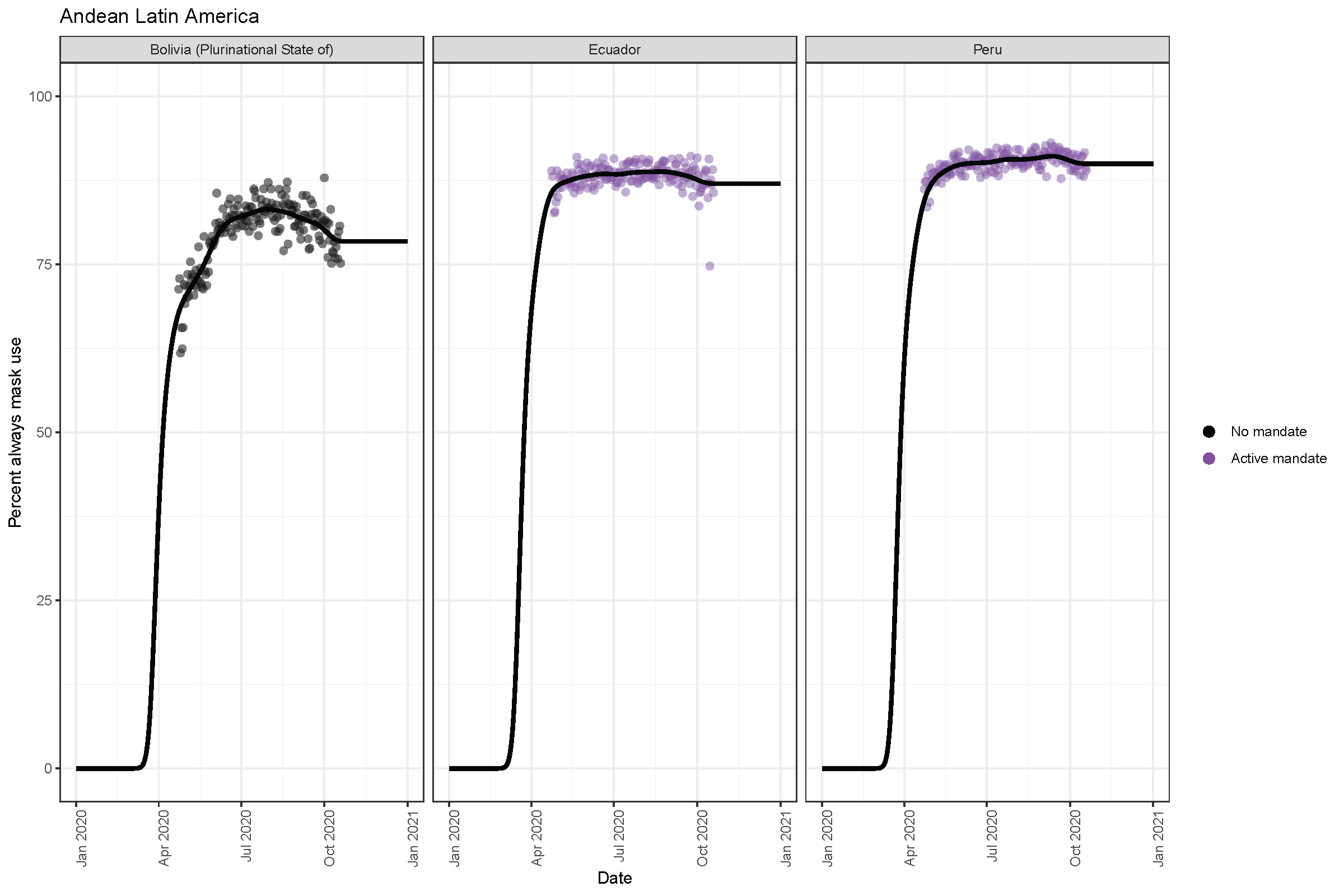

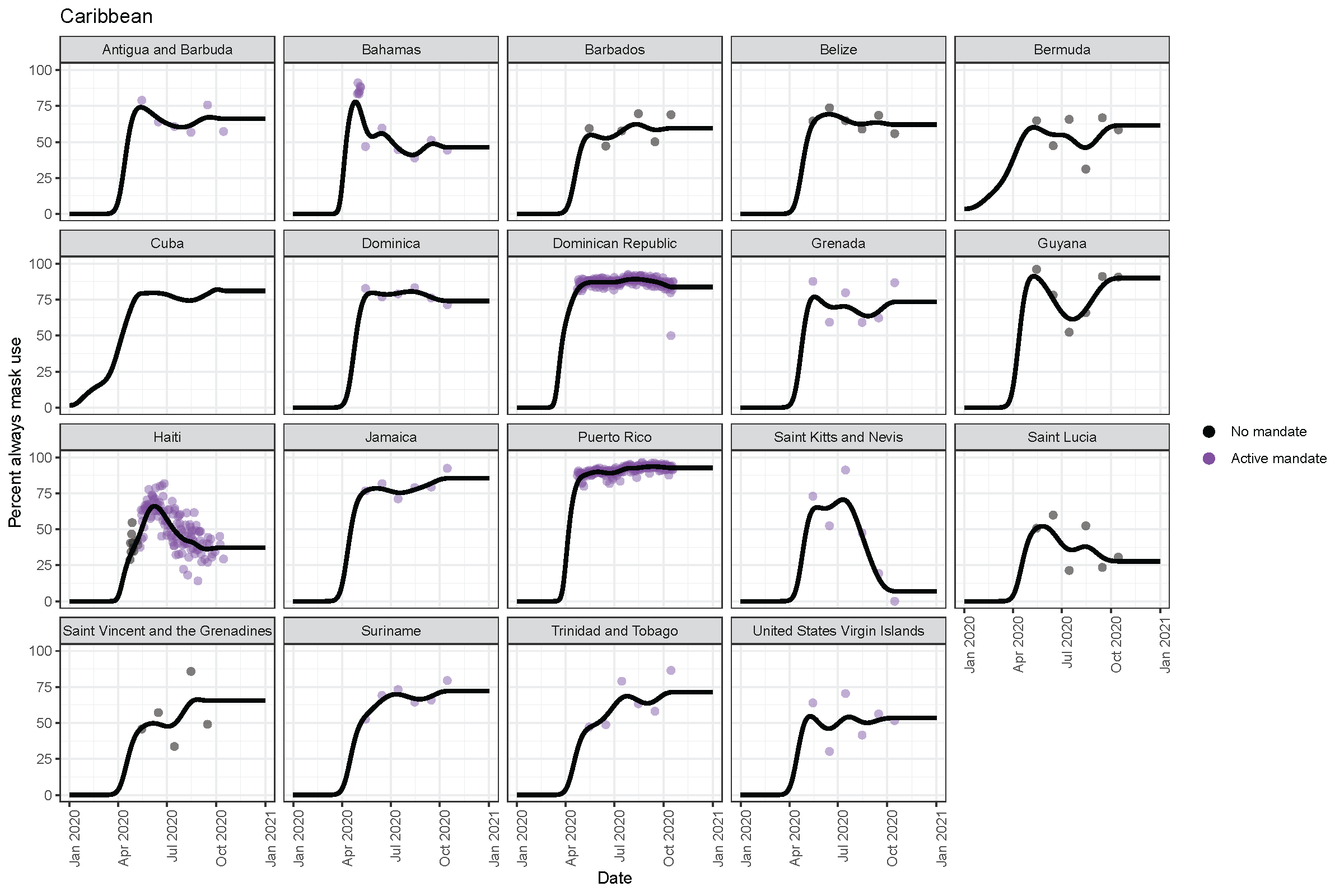

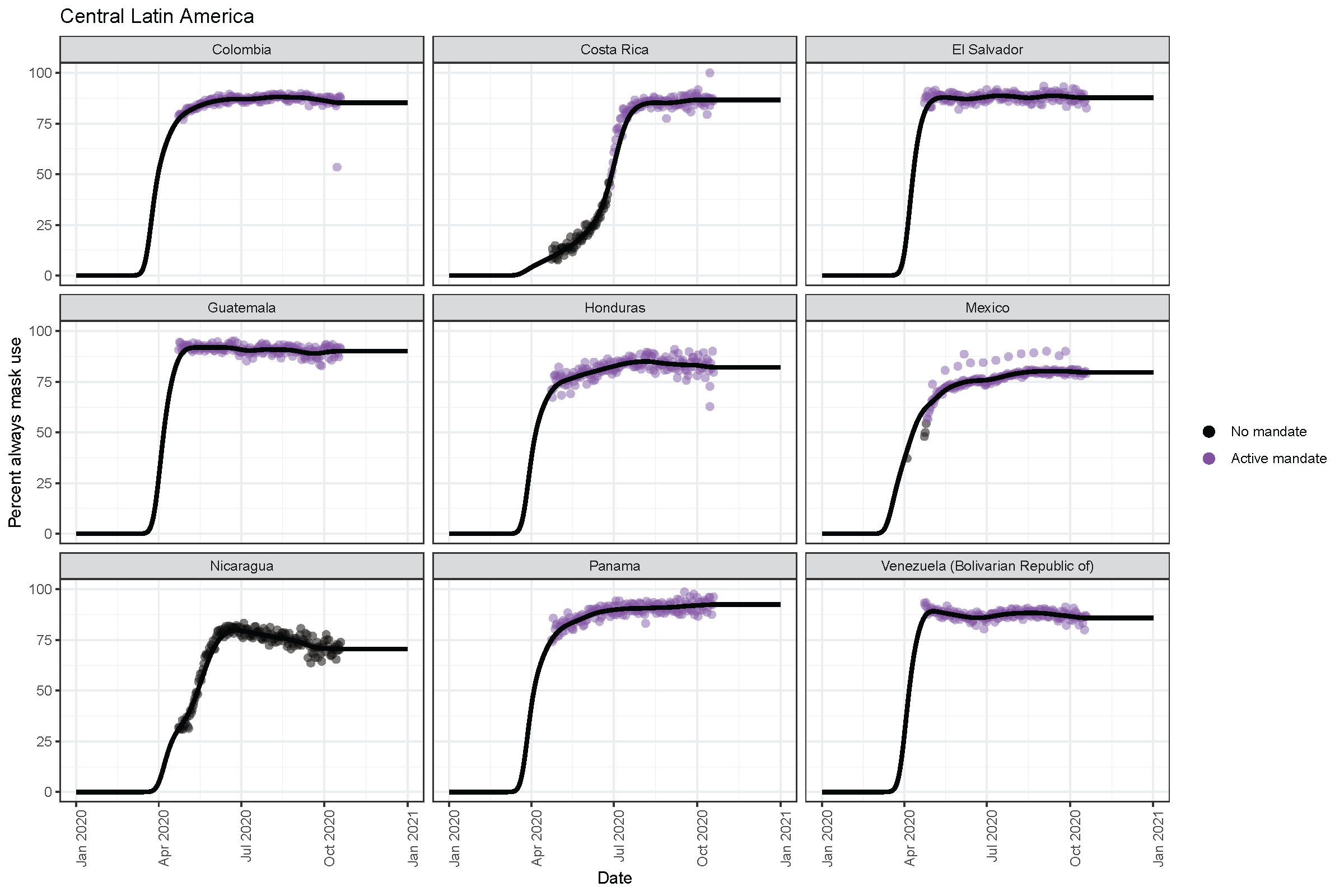

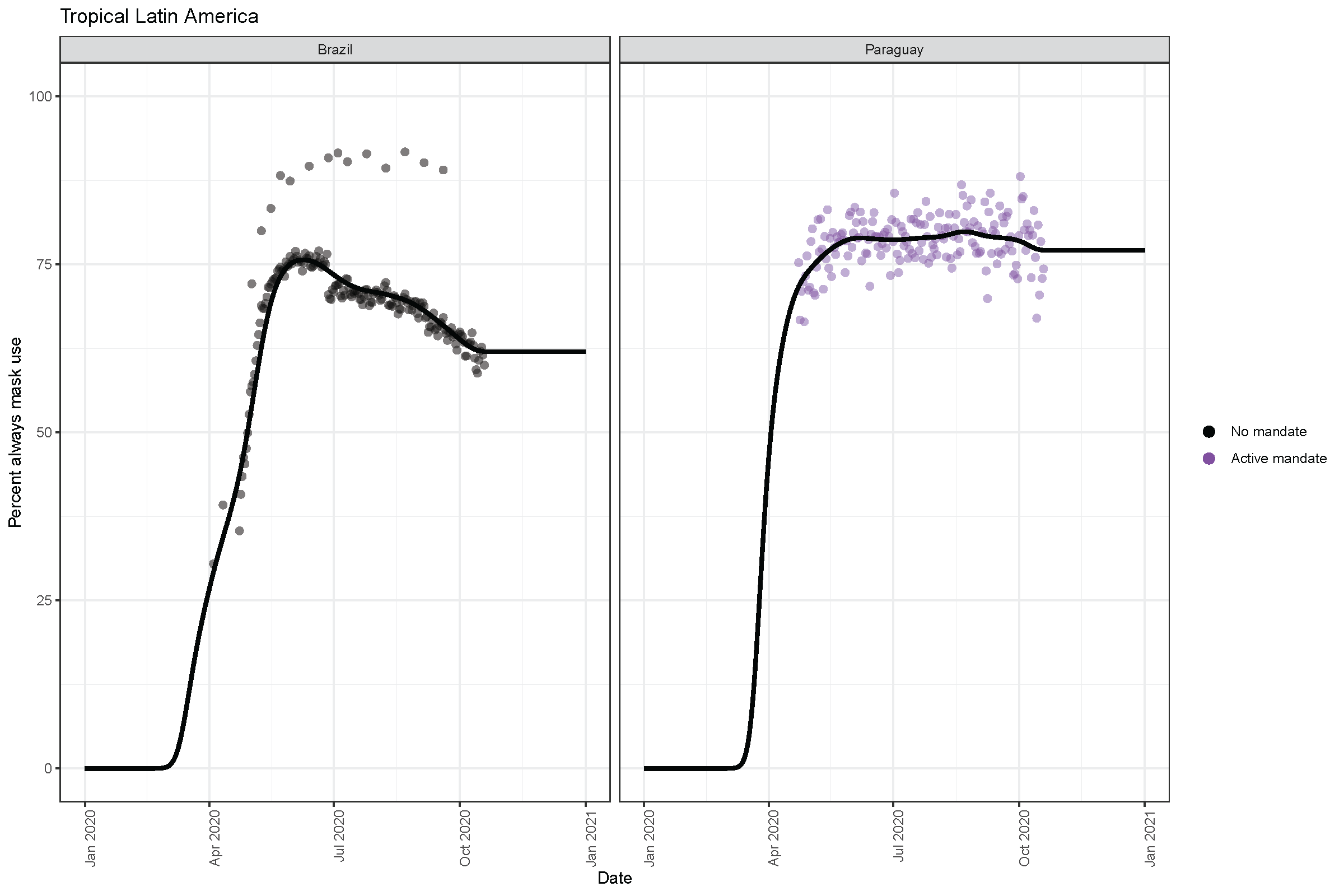

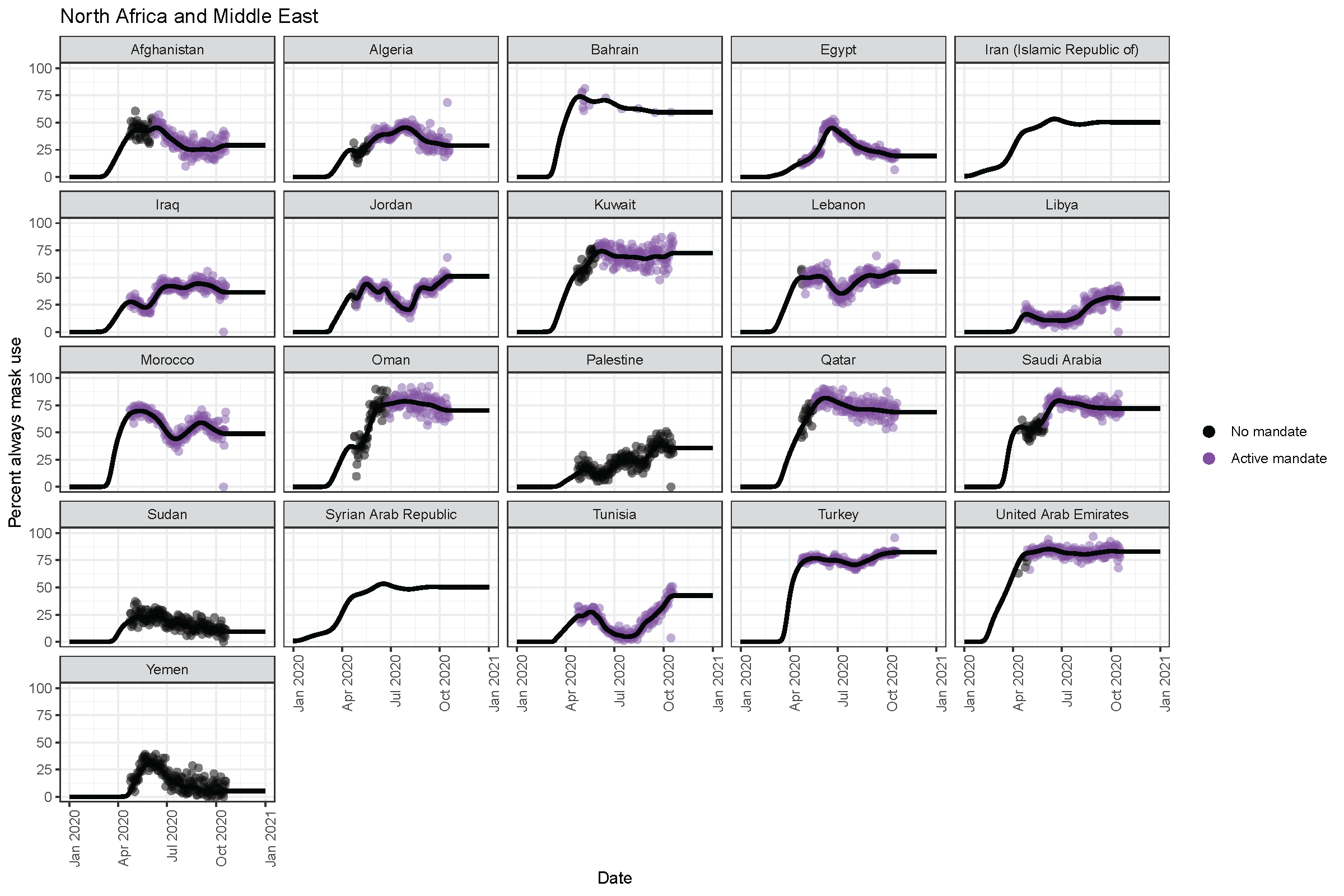

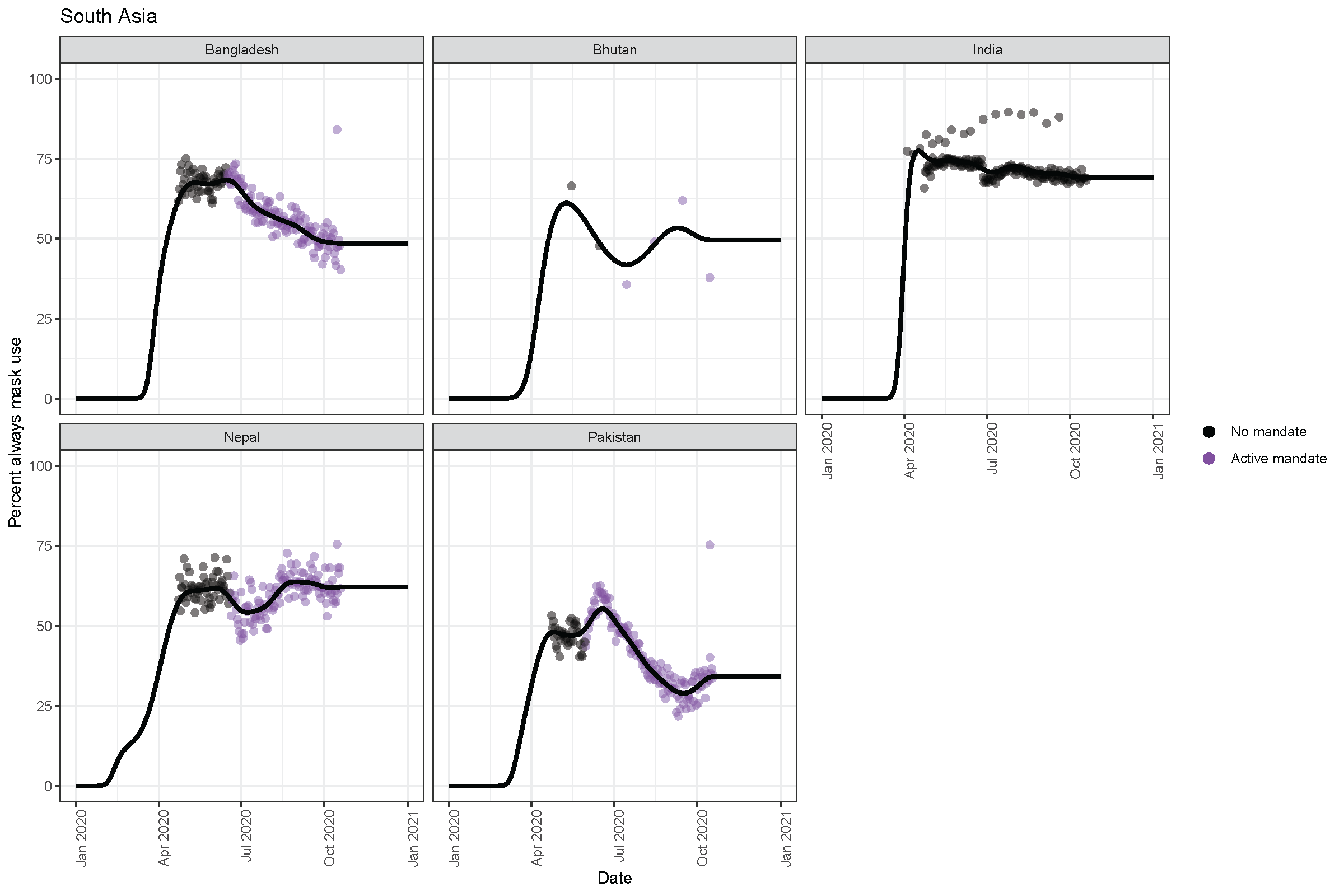

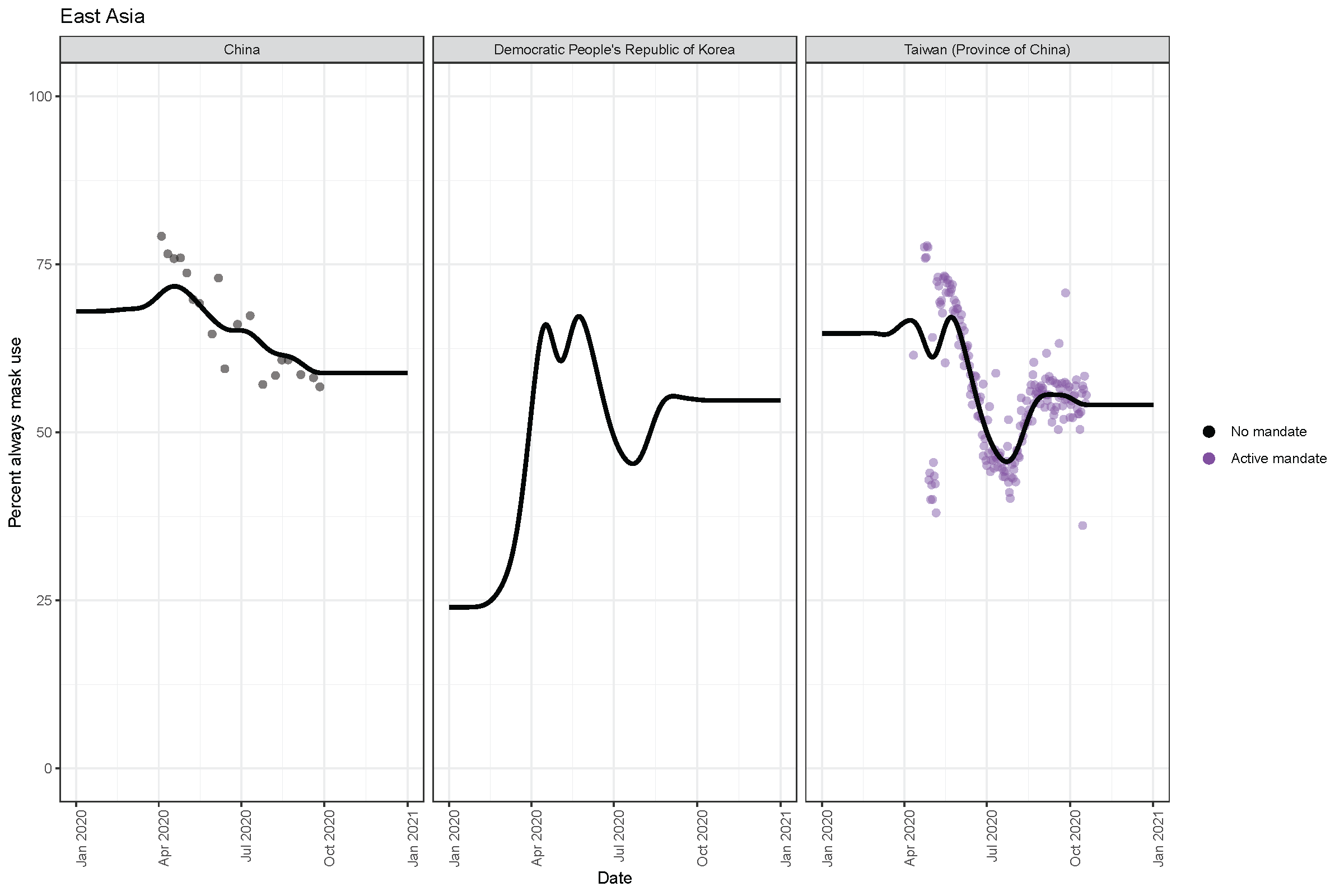

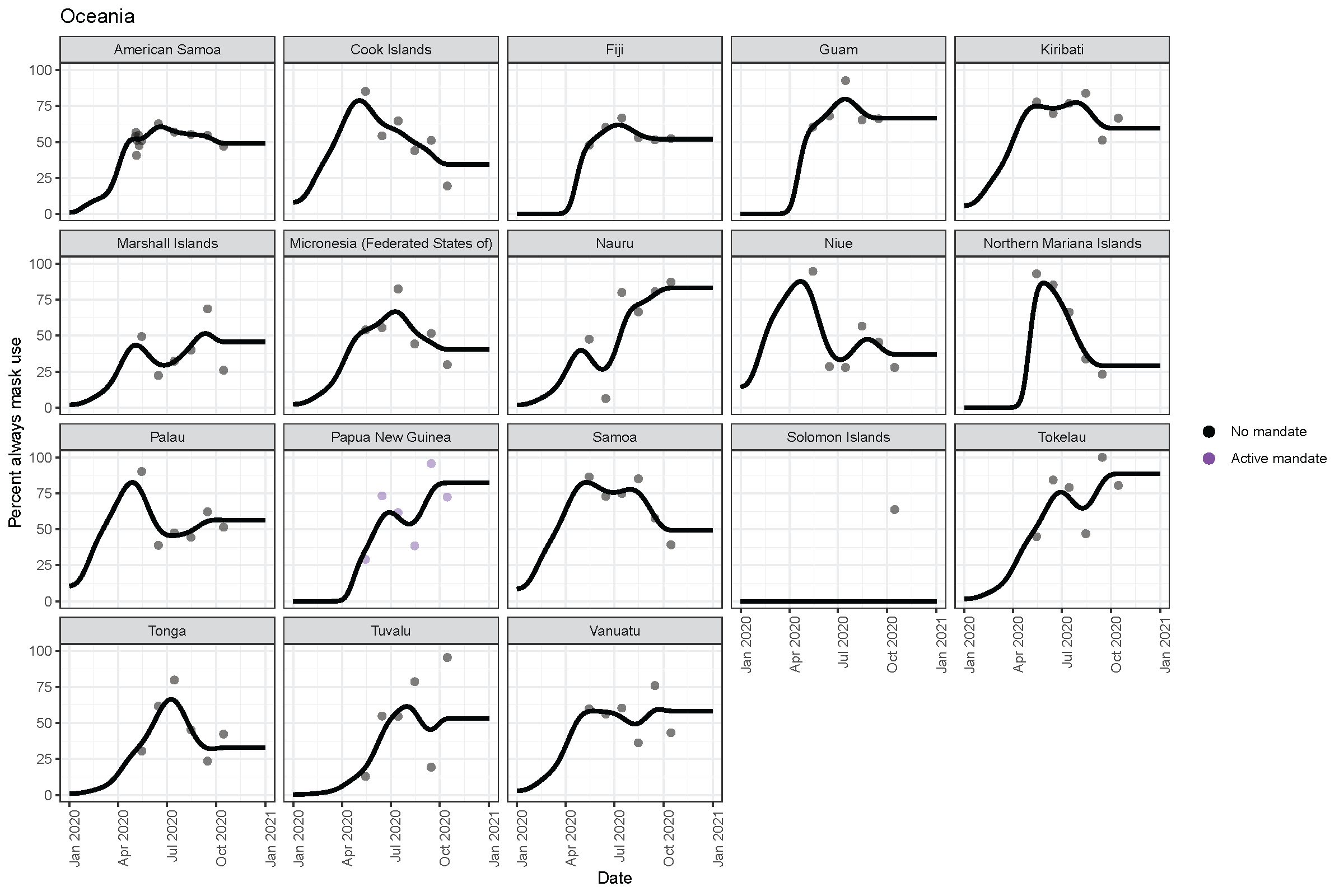

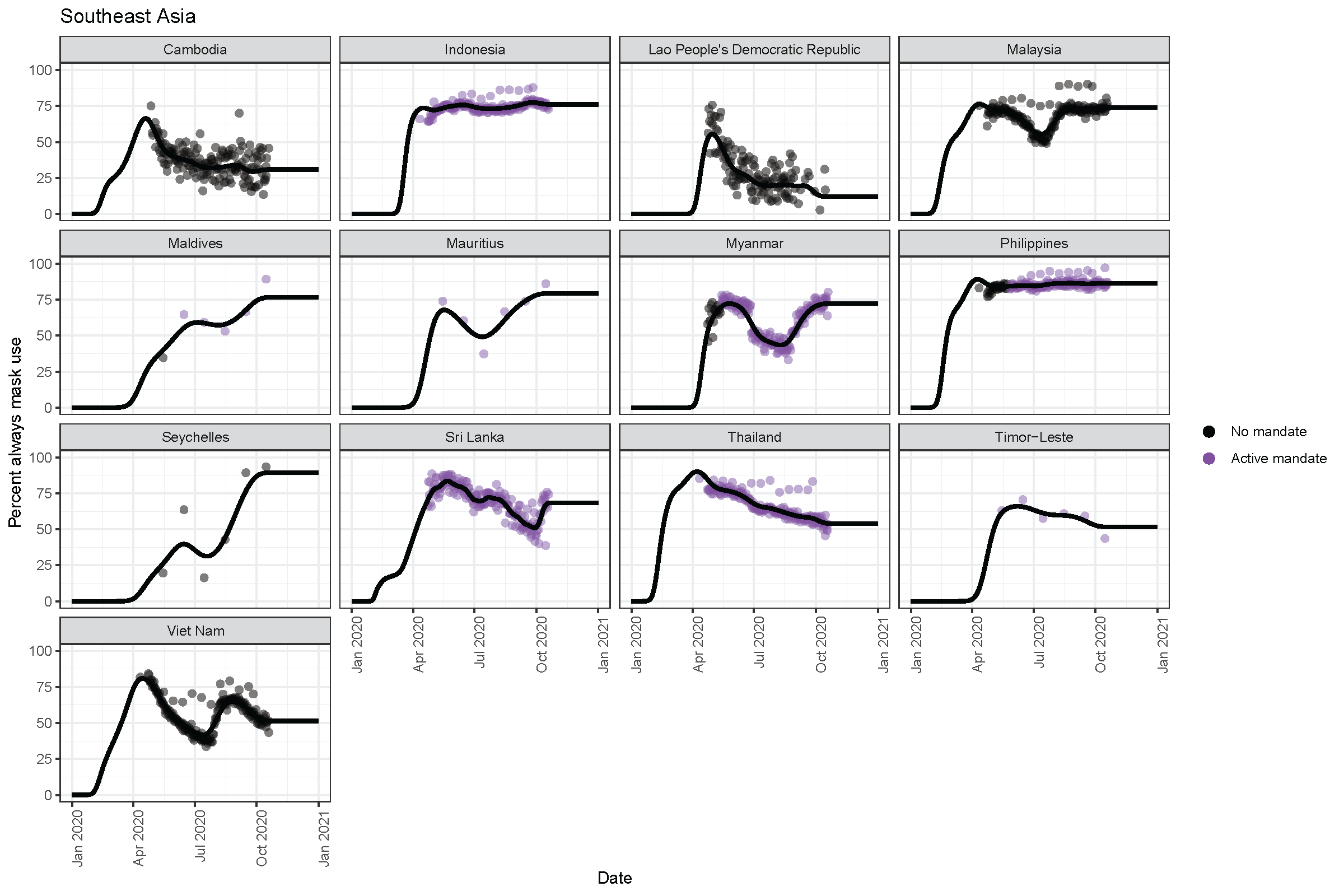

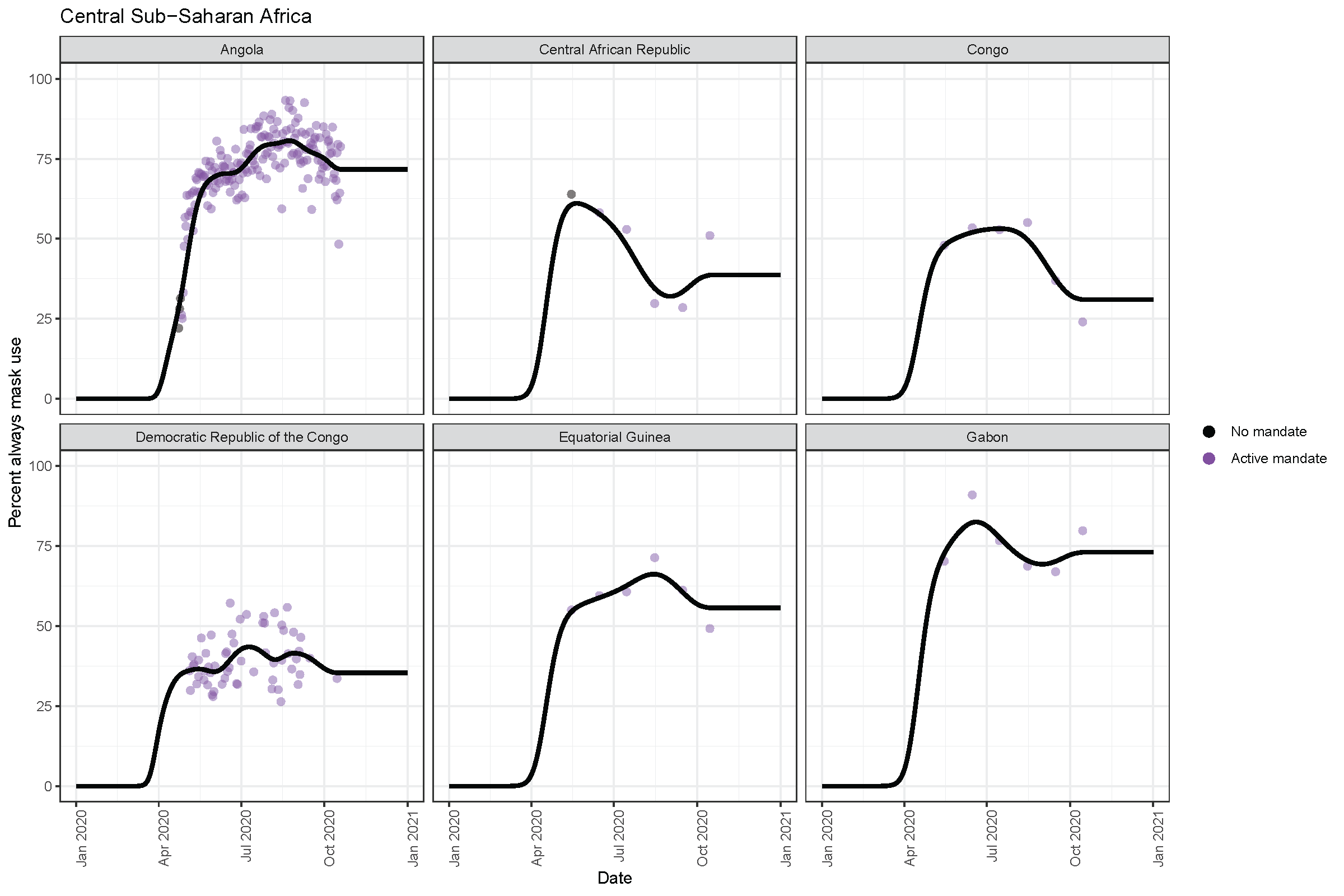

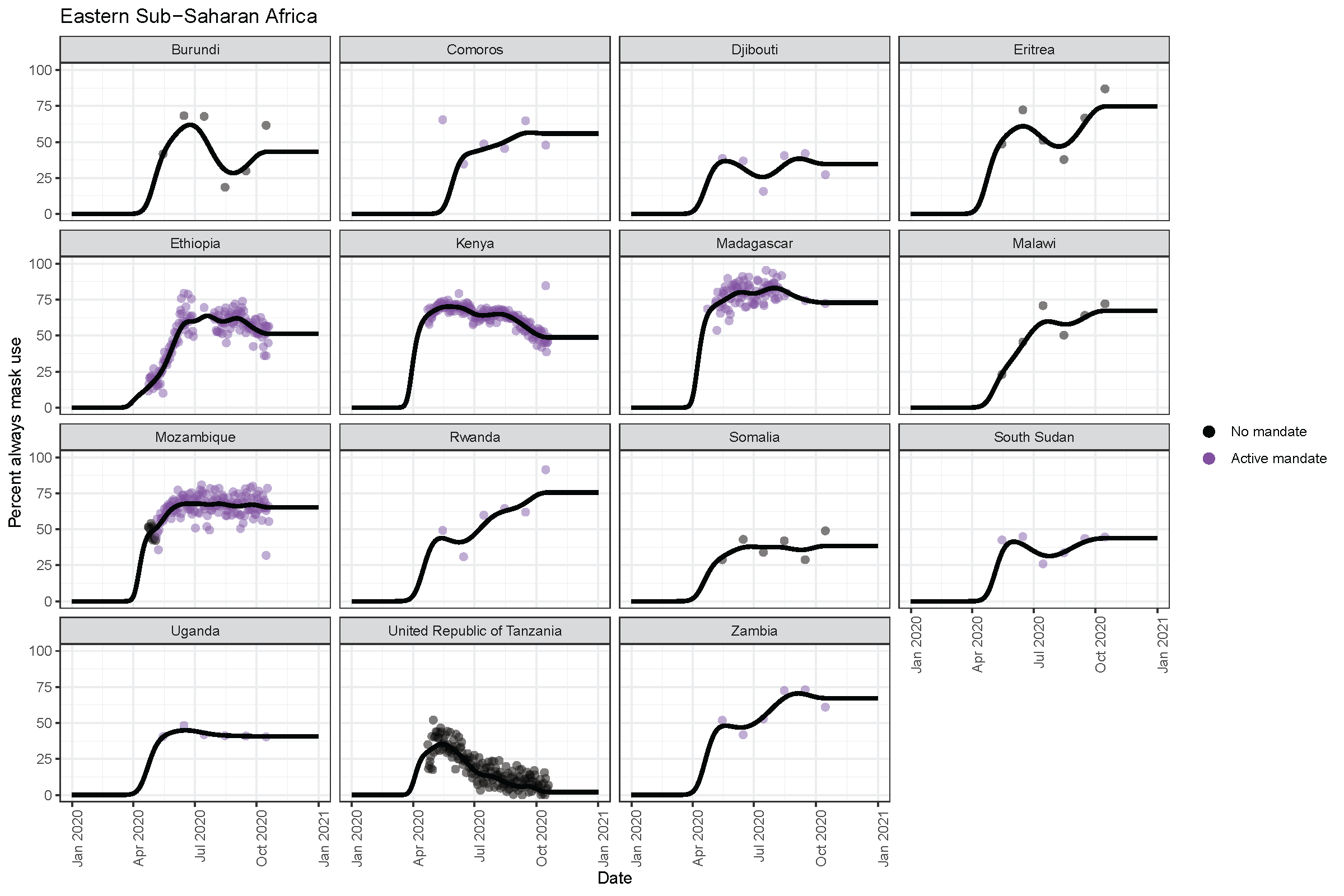

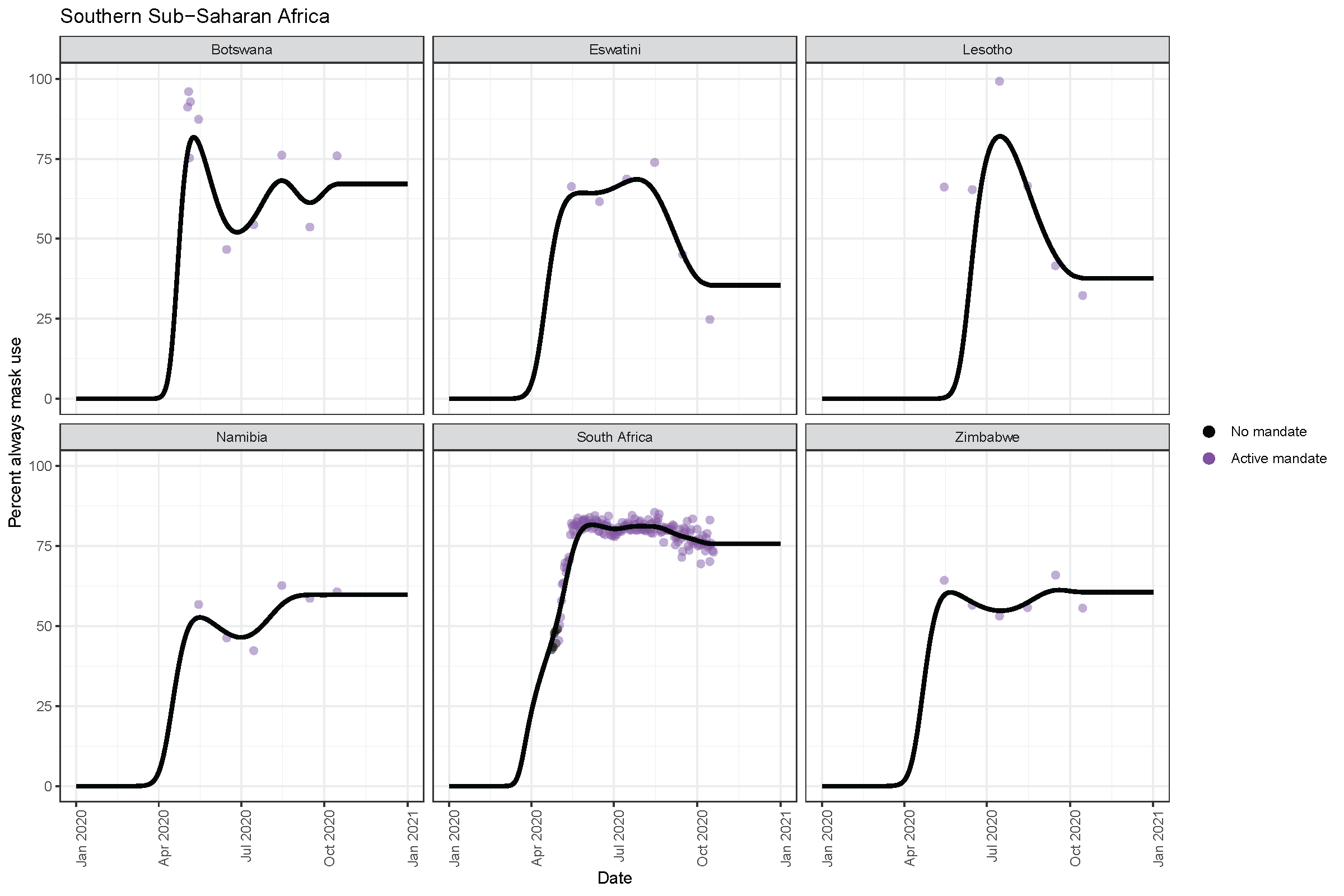

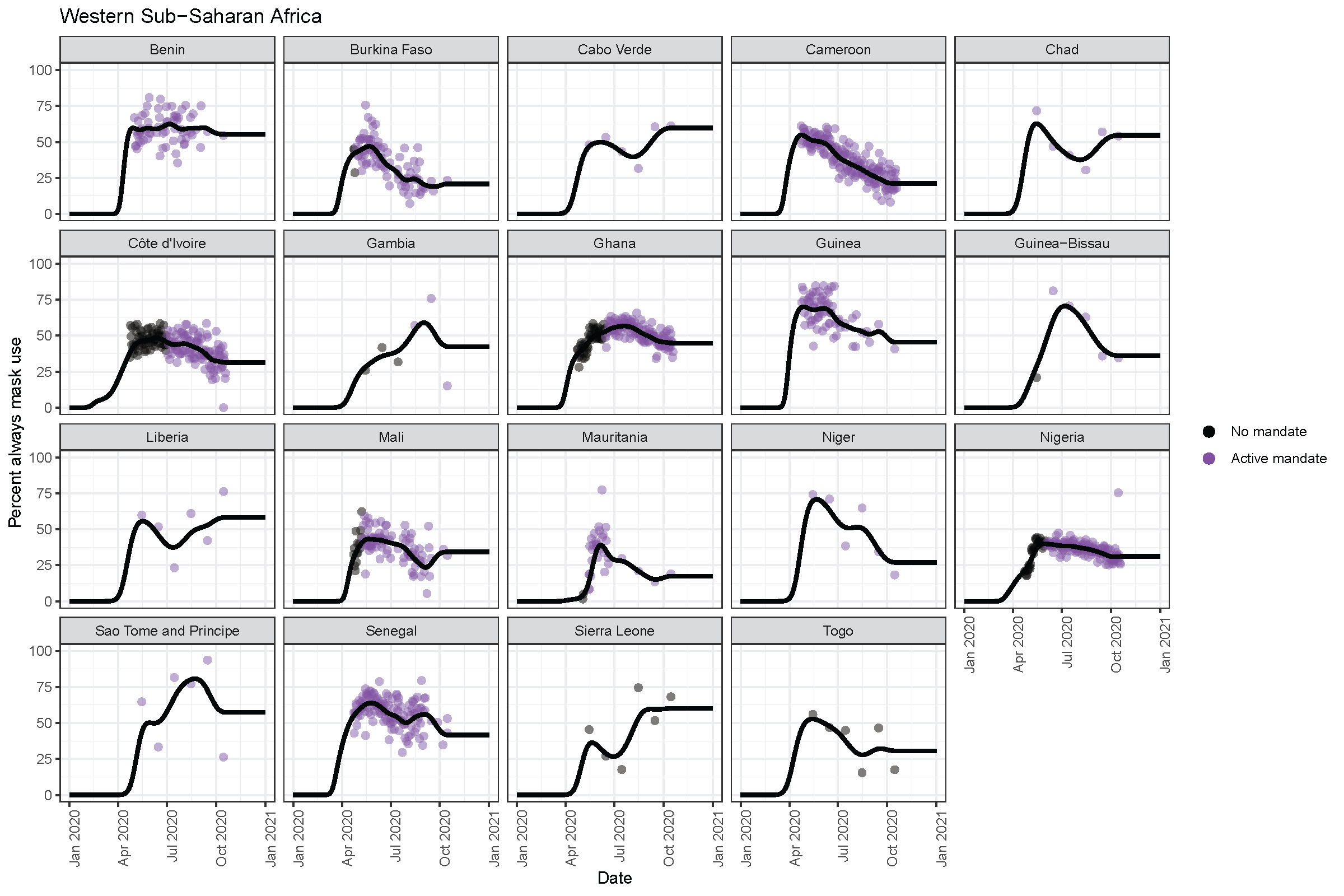

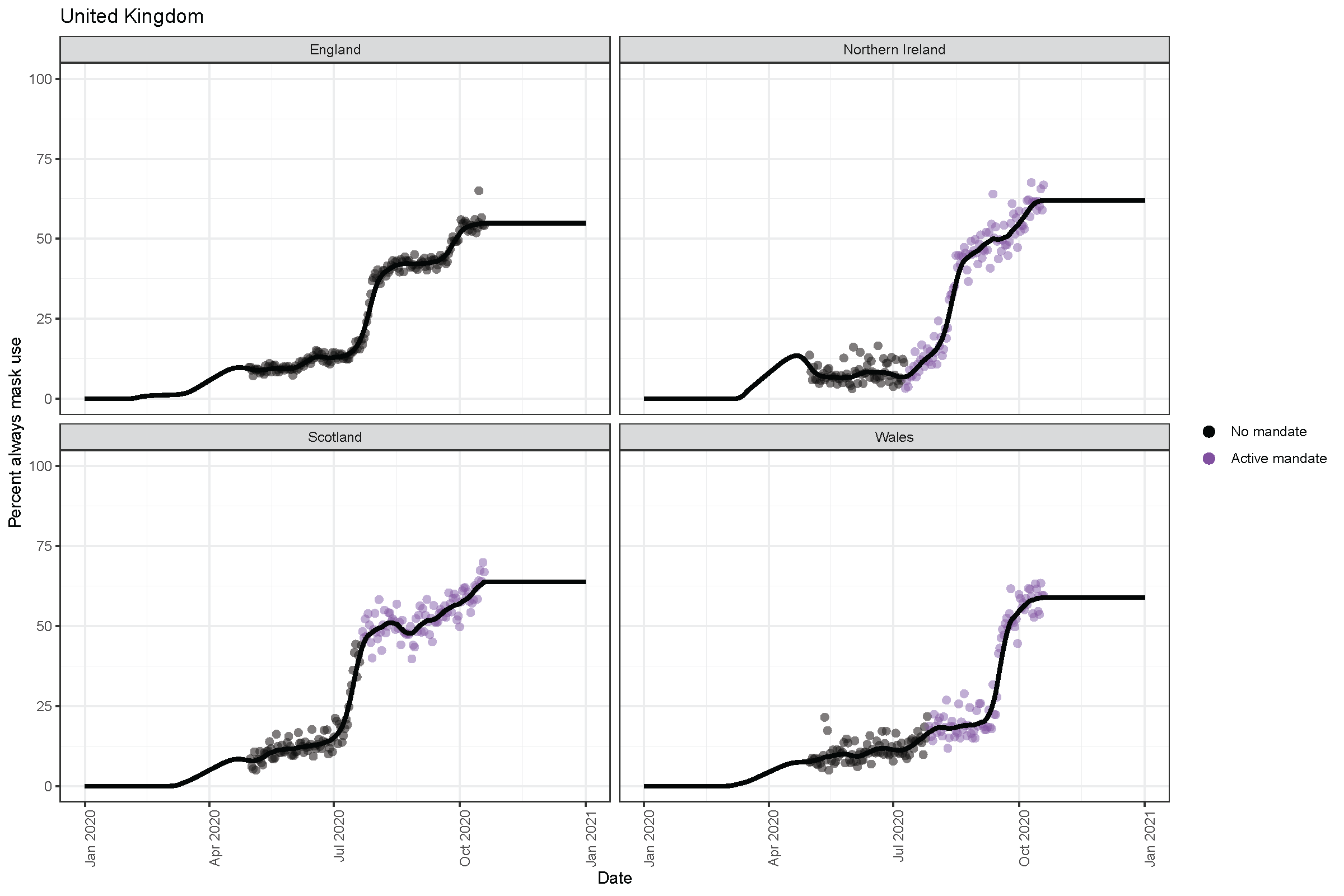

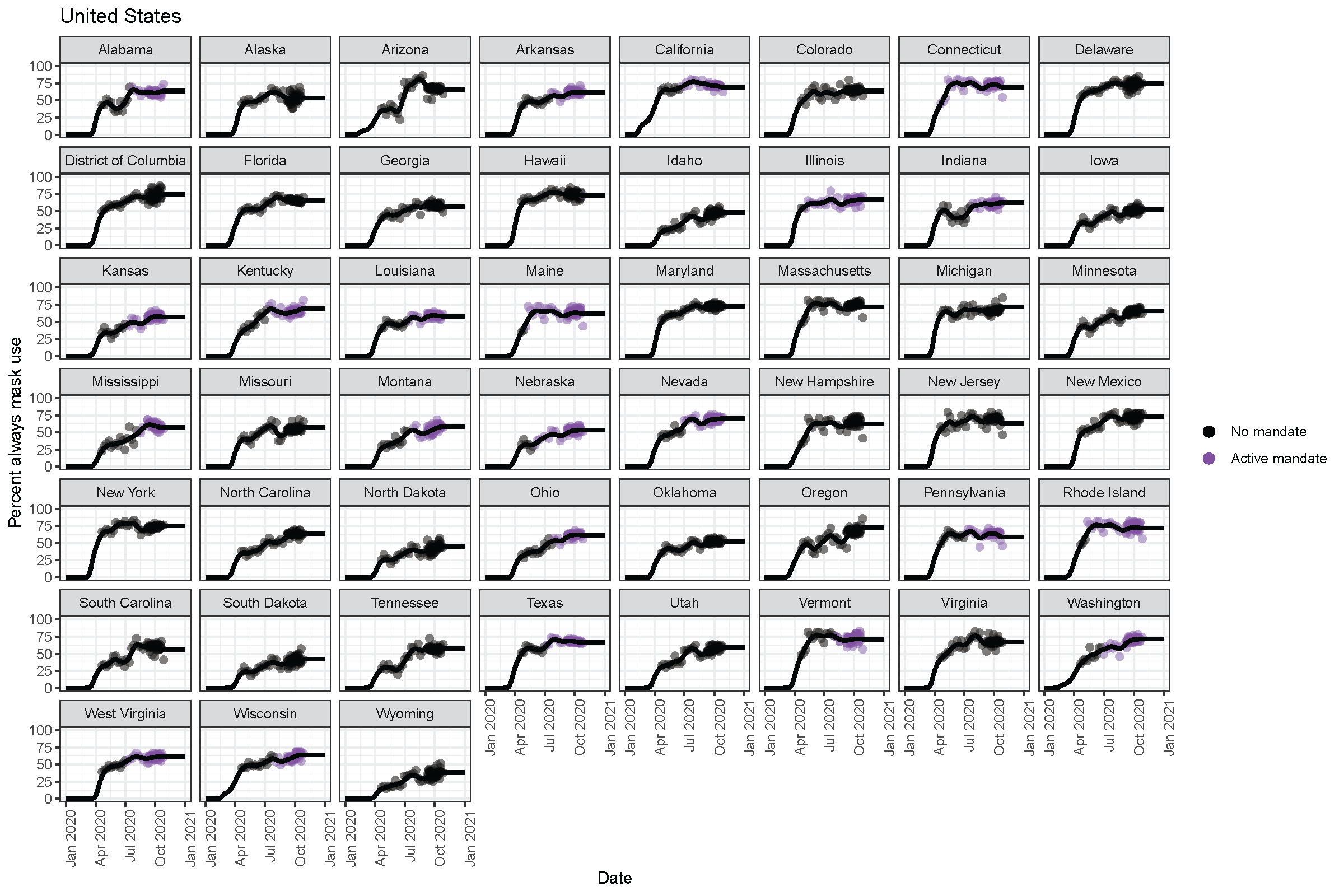

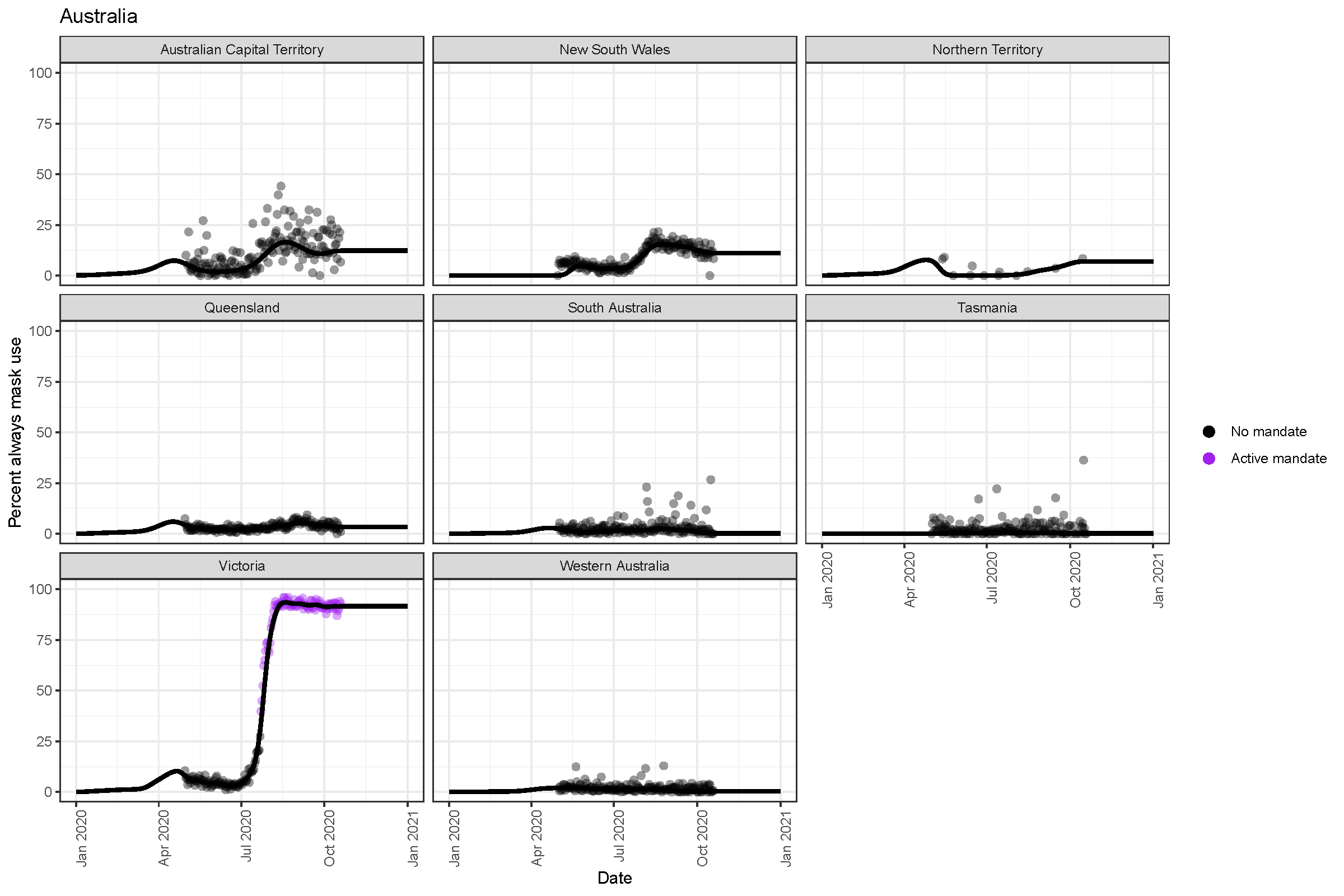

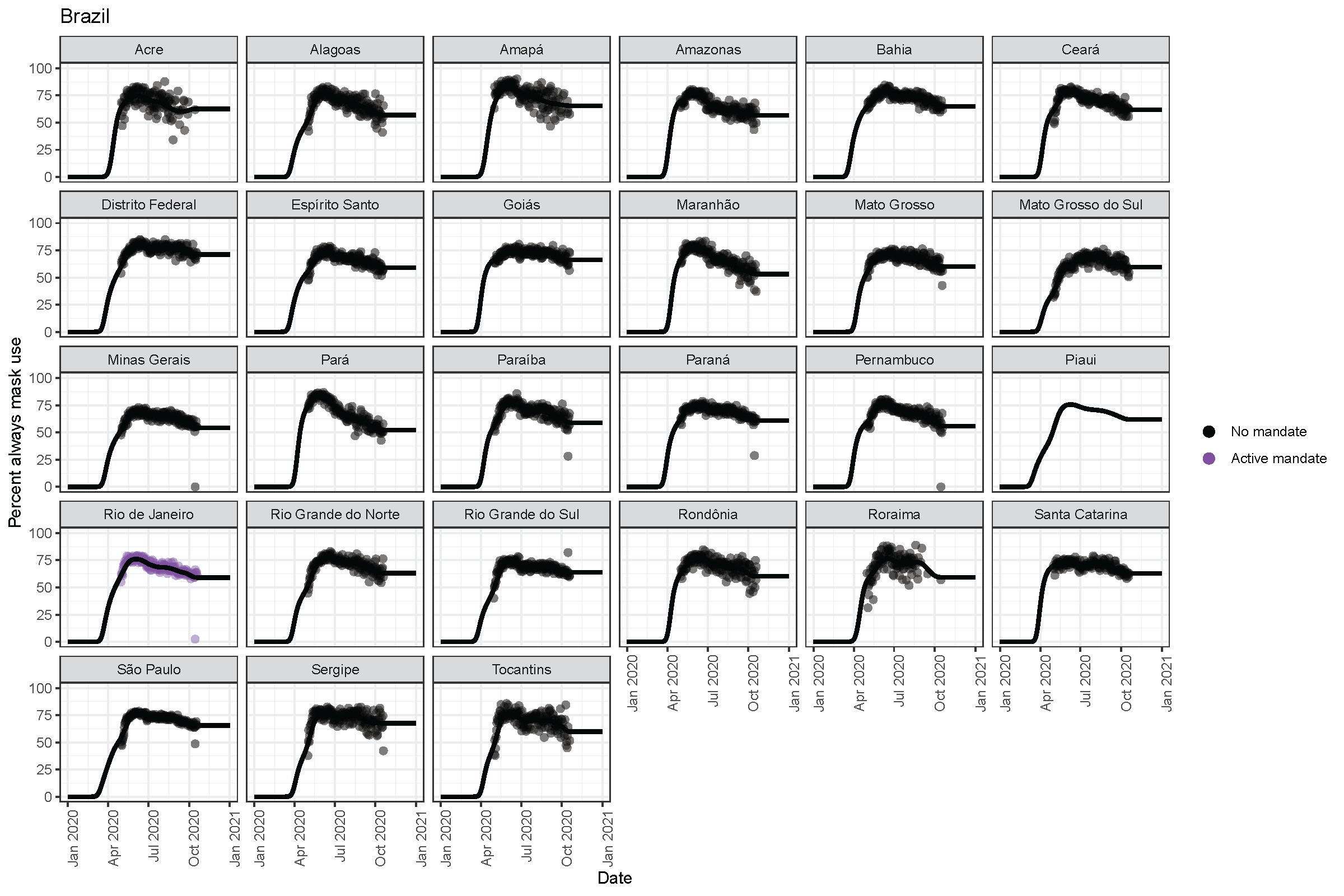

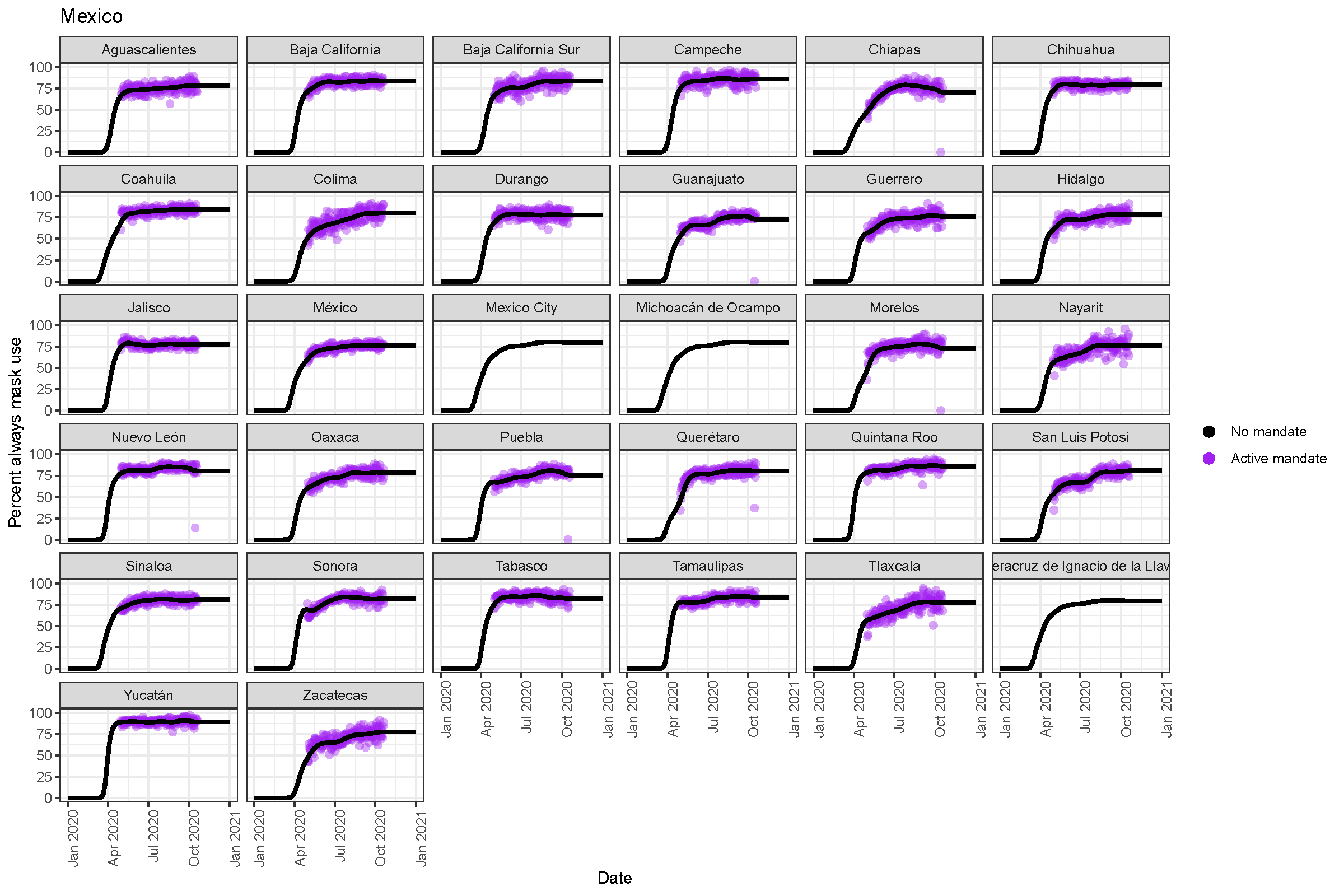

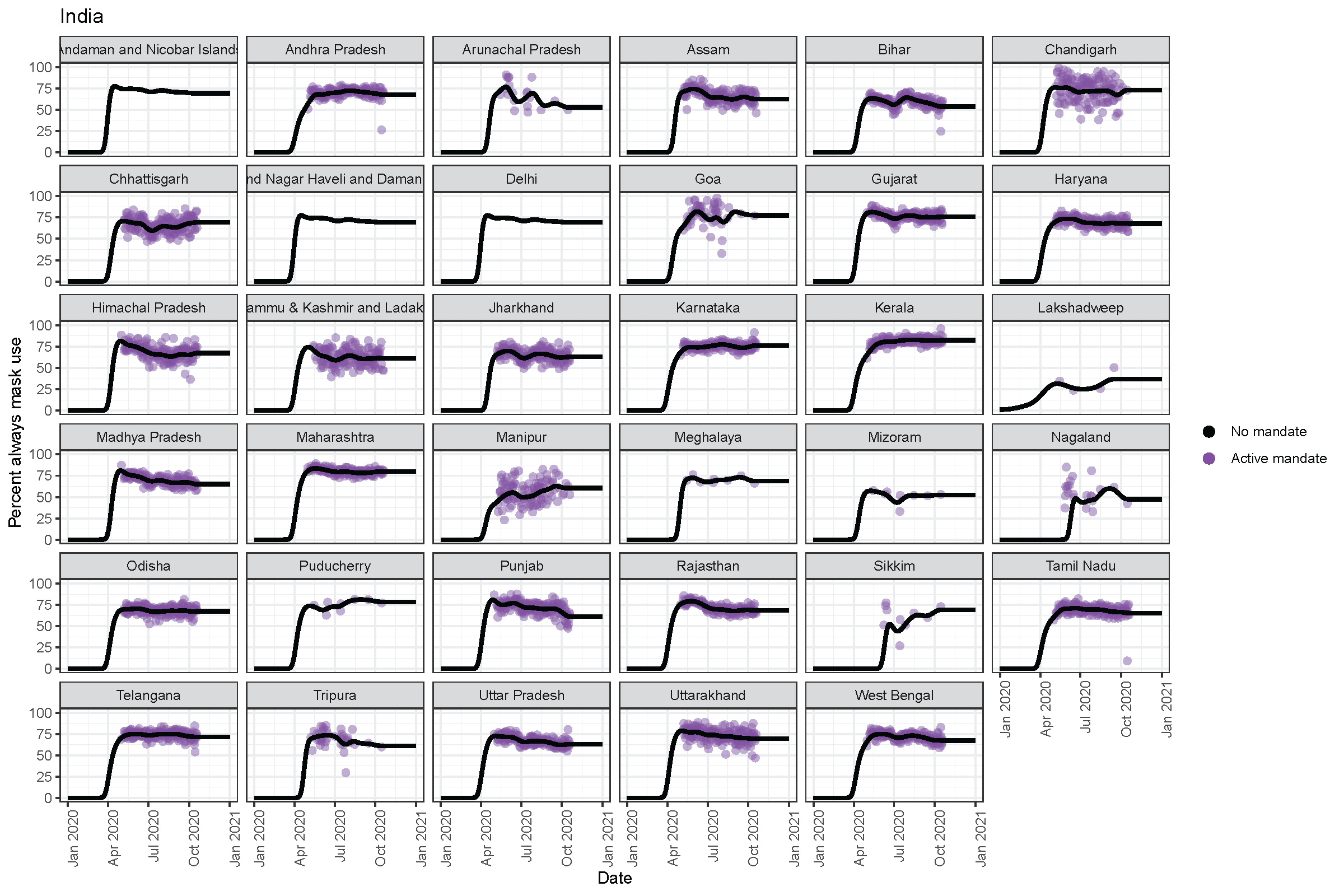

#### Figure S15. Active mask mandates as of October 26, 2020

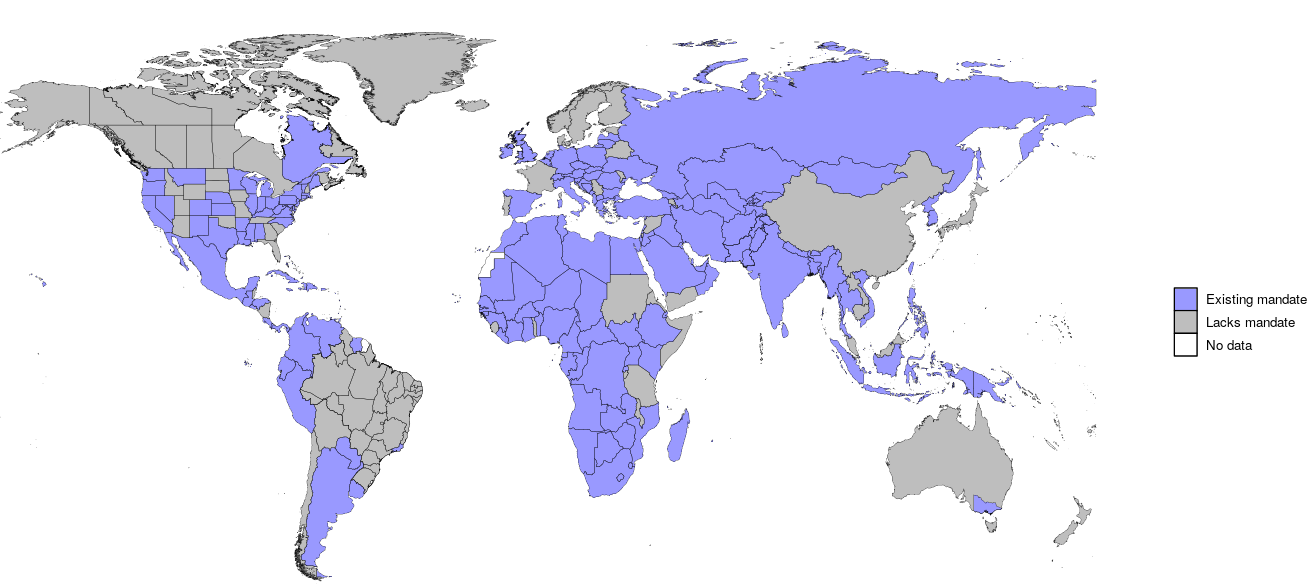

Mandates represent those targeted to the general public or to businesses. Canada has a nationwide mandate for mask use among travelers and Mississippi rescinded a state-wide mask mandate but re-imposed one later on high-transmission areas of the state.

Figure S16. Cumulative deaths from January 1, 2020 to January 1, 2021 in the reference, universal mask use scenario (95% coverage), and an alternative 85% mask use coverage scenario, by GBD super-region
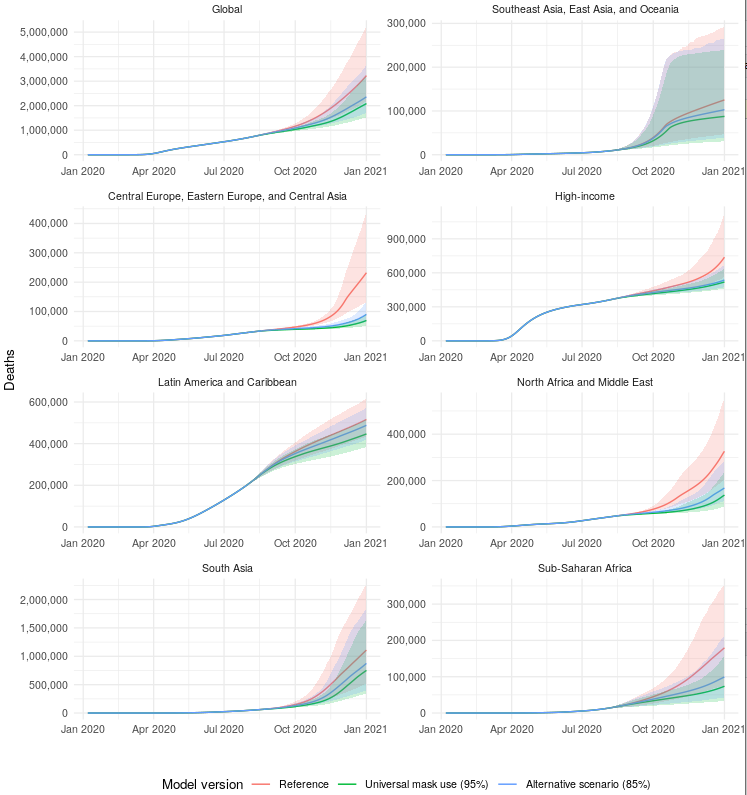

#### Table S1. Meta-regression publication characteristics

| **Characteristic** | | **N (%) of observations** | **Studies** |
| --- | --- | --- | --- |
| **Study location*** | **Southeast Asia** | 27 (67.5%) | 3, 6-31 |
|  | **Outside southeast Asia** | 13 (32.5%) | 32-44 |
| **Disease studied** | **SARS-CoV 1 or 2** | 34 (54%) | 3, 10-13, 15-24, 30-37 |
|  | **Not SARS-CoV 1 or 2** | 29 (56%) | 6-9, 14, 25-29, 38-44 |
| **Mask type studied** | **Cloth masks** | 2 (3%) | 14,24 |
|  | **Non-descript masks** | 17 (27%) | 3, 13-17, 20, 22, 30, 31, 41, 43 |
|  | **Medical, surgical, or N95 masks** | 44 (70%) | 6–12, 14, 17–19, 21–23, 25–29, 32–40, 42, 44 |
| **Diagnosis** | **Laboratory** | 53 (84%) | 3, 6-11, 14, 17–21, 23–27, 29–32, 34–44 |
|  | **Clinical** | 10 (16%) | 12, 13, 15, 16, 22, 28, 33 |
| **Population and setting** | **General** | 13 (21%) | 3, 7, 8, 15, 16, 31, 38, 39, 41 |
|  | **Healthcare** | 50 (79%) | 6, 9–14, 17–30, 32–37, 42–44 |
| **Type of control group** | **Infrequent mask use** | 14 (22%) | 10, 14, 21, 25, 30, 32, 34, 36, 42, 43 |
|  | **No mask use** | 47 (75%) | 3, 6–9, 11–20, 22, 23, 26–29, 31, 33, 35, 37–39, 41, 43, 44 |
|  | **Pre- and post-intervention mask wear** | 1 (2%) | 40 |
|  | **Missing** | 1 (2%) | 24 |

*This row presents the results at the study-level (N=40) rather than the observation level (N=63) as the study location did not vary within an individual study

#### Table S2. Individual publication characteristics

| **Author** | **Mask Wearers, Infection +** | **Mask Wearers** | **Mask Non-wearers, Infection +** | **Mask Non-wearers** | **Study Country** | **Disease** | **Diagnosis** | **Mask Type** | **Study Design** | **Healthcare Setting** | **Intervention Group Type** | **Control Group Type** | **Citation** |
| --- | --- | --- | --- | --- | --- | --- | --- | --- | --- | --- | --- | --- | --- |
| Wang | 8 | 46 | 17 | 41 | China | SARS-CoV-2 | Laboratory | Non-medical mask | Cohort | No | Consistent Use | No mask use | 3 |
| Wang | 24 | 83 | 17 | 41 | China | SARS-CoV-2 | Laboratory | Non-medical mask | Cohort | No | Any Use | No mask use | 3 |
| Chokephaibulkit | 30 | 239 | 3 | 17 | Thailand | H1N1 | Laboratory | Medical mask | Case Control | Yes | Non-medical mask | No mask use | 6 |
| Chokephaibulkit | 16 | 142 | 3 | 17 | Thailand | H1N1 | Laboratory | Medical mask (N95) | Case Control | Yes | Medical mask (N95) | No mask use | 6 |
| Chokephaibulkit | 10 | 78 | 3 | 17 | Thailand | H1N1 | Laboratory | Medical mask | Case Control | Yes | Medical mask | No mask use | 6 |
| Cowling | 4 | 61 | 12 | 205 | China (Hong Kong) | Influenza | Laboratory | Medical mask | cRCT | No | Facemask | No mask use | 7 |
| Cowling | 18 | 258 | 28 | 279 | China (Hong Kong) | Influenza | Laboratory | Medical mask | cRCT | No | Facemask + Hygiene | No mask use | 8 |
| MacIntyre | 3 | 949 | 6 | 481 | China | Influenza | Laboratory | Medical mask (N95) | cRCT | Yes | Medical mask (N95) | No mask use | 9 |
| MacIntyre | 5 | 492 | 6 | 481 | China | Influenza | Laboratory | Medical mask | cRCT | Yes | Medical mask | No mask use | 9 |
| MacIntyre | 13 | 949 | 15 | 481 | China | Respiratory Virus | Laboratory | Medical mask (N95) | cRCT | Yes | Medical mask (N95) | No mask use | 9 |
| MacIntyre | 13 | 492 | 15 | 481 | China | Respiratory Virus | Laboratory | Medical mask | cRCT | Yes | Medical mask | No mask use | 9 |
| Nishiura | 8 | 43 | 17 | 72 | Vietnam | SARS | Laboratory | Medical mask | Case Control | Yes | Consistent Use | Inconsistent use | 10 |
| Nishiura | 1 | 26 | 3 | 4 | Vietnam | SARS | Laboratory | Medical mask | Case Control | Yes | Consistent Use | Inconsistent use | 10 |
| Wilder-Smith | 3 | 24 | 37 | 68 | Singapore | SARS | Laboratory | Medical mask (N95) | Case Control | Yes | Any Use | No mask use | 11 |
| Wilder-Smith | 6 | 27 | 34 | 65 | Singapore | SARS | Laboratory | Medical mask (N95) | Case Control | Yes | Any Use | No mask use | 11 |
| Teleman | 3 | 26 | 33 | 60 | Singapore | SARS | Clinical | Medical mask (N95) | Case Control | Yes | Medical mask (N95) | No mask use | 12 |
| Yin | 68 | 246 | 9 | 11 | China | SARS | Clinical | Non-medical mask | Case Control | Yes | Any Use | No mask use | 13 |
| Zhang | 33 | 152 | 2 | 12 | China | H1N1 | Laboratory | Non-medical mask | Case Control | Yes | Consistent Use | Inconsistent use | 14 |
| Zhang | 44 | 222 | 2 | 12 | China | H1N1 | Laboratory | Non-medical mask | Case Control | Yes | Any Use | Inconsistent use | 14 |
| Zhang | 3 | 16 | 2 | 12 | China | H1N1 | Laboratory | Medical mask (N95) | Case Control | Yes | Medical mask (N95) | No mask use | 14 |
| Zhang | 37 | 183 | 2 | 12 | China | H1N1 | Laboratory | Medical mask | Case Control | Yes | Medical mask | No mask use | 14 |
| Zhang | 9 | 44 | 2 | 12 | China | H1N1 | Laboratory | Non-medical mask | Case Control | Yes | Non-medical mask | No mask use | 14 |
| Lau | 8 | 94 | 17 | 98 | Hong Kong | SARS | Clinical | Non-medical mask | Case Control | No | Any Use | No mask use | 15 |
| Wu | 25 | 146 | 44 | 120 | China | SARS | Clinical | Non-medical mask | Case Control | No | Consistent Use | No mask use | 16 |
| Wu | 50 | 255 | 44 | 120 | China | SARS | Clinical | Non-medical mask | Case Control | No | Any Use | No mask use | 16 |
| Liu | 8 | 123 | 43 | 354 | China | SARS | Laboratory | Medical mask | Case Control | Yes | Any Use | No mask use | 17 |
| Liu | 15 | 274 | 36 | 203 | China | SARS | Laboratory | Medical mask | Case Control | Yes | Any Use | No mask use | 17 |
| Liu | 2 | 33 | 49 | 444 | China | SARS | Laboratory | Medical mask (N95) | Case Control | Yes | Any Use | No mask use | 17 |
| Liu | 11 | 95 | 40 | 382 | China | SARS | Laboratory | Non-medical mask | Case Control | Yes | Any Use | No mask use | 17 |
| Wang | 0 | 278 | 10 | 215 | China | SARS-CoV-2 | Laboratory | Medical mask (N95) | Case Control | Yes | Any Use | No mask use | 18 |
| Wang | 1 | 1286 | 119 | 4036 | China | SARS-CoV-2 | Laboratory | Medical mask | Case Control | Yes | Any Use | No mask use | 19 |
| Nishiyama | 17 | 61 | 14 | 18 | Vietnam | SARS | Laboratory | Non-medical mask | Cohort | Yes | Consistent Use | No mask use | 20 |
| Reynolds | 8 | 42 | 14 | 25 | Vietnam | SARS | Laboratory | Medical mask | Case Control | Yes | Consistent Use | Inconsistent use | 21 |
| Seto | 2 | 13 | 26 | 98 | China | SARS | Clinical | Non-medical mask | Case Control | Yes | Any Use | No mask use | 22 |
| Seto | 0 | 11 | 51 | 123 | China | SARS | Clinical | Medical mask | Case Control | Yes | Any Use | No mask use | 22 |
| Seto | 0 | 11 | 92 | 164 | China | SARS | Clinical | Medical mask (N95) | Case Control | Yes | Any Use | No mask use | 22 |
| Ho | 2 | 62 | 2 | 10 | China | SARS | Laboratory | Medical mask (N95) | Cohort | Yes | Any Use | No mask use | 23 |
| Pei | 11 | 98 | 61 | 115 | China | SARS | Laboratory | Non-medical mask | Case Control | Yes | MISSING | MISSING | 24 |
| Kim | 1 | 444 | 16 | 308 | South Korea | MERS | Laboratory | Medical mask (N95) | Case Control | Yes | Consistent Use | Inconsistent use | 25 |
| Kim | 0 | 7 | 1 | 2 | South Korea | MERS | Laboratory | Non-medical mask | Case Control | Yes | Any Use | No mask use | 26 |
| Cheng | 0 | 568 | 4 | 268 | China (Hong Kong) | H1N1 | Laboratory | Medical mask | Case Control | Yes | Any Use | No mask use | 27 |
| Park | 3 | 24 | 2 | 4 | South Korea | MERS | Clinical | Medical mask | Case Control | Yes | Any Use | No mask use | 28 |
| Ki | 0 | 218 | 6 | 230 | South Korea | MERS | Laboratory | Medical mask | Case Control | Yes | Any Use | No mask use | 29 |
| Ha | 0 | 61 | 0 | 1 | Vietnam | SARS | Laboratory | Non-medical mask | Cohort | Yes | Consistent Use | Inconsistent use | 30 |
| Tuan | 0 | 9 | 7 | 154 | Vietnam | SARS | Laboratory | Non-medical mask | Cohort | No | Consistent Use | No mask use | 31 |
| Loeb | 3 | 23 | 5 | 9 | Canada | SARS | Laboratory | Medical mask | Case Control | Yes | Consistent Use | Inconsistent use | 32 |
| Loeb | 2 | 16 | 5 | 9 | Canada | SARS | Laboratory | Medical mask (N95) | Case Control | Yes | Consistent Use | Inconsistent use | 32 |
| Loeb | 1 | 4 | 5 | 9 | Canada | SARS | Laboratory | Medical mask | Case Control | Yes | Consistent Use | Inconsistent use | 32 |
| Scales | 3 | 16 | 4 | 15 | Canada | SARS | Clinical | Medical mask | Case Control | Yes | Any Use | No mask use | 33 |
| Heinzerling | 0 | 3 | 3 | 34 | USA | SARS-CoV-2 | Laboratory | Medical mask | Case Control | Yes | Consistent Use | Inconsistent use | 34 |
| Park | 0 | 57 | 0 | 45 | USA | SARS | Laboratory | Medical mask | Cohort | Yes | Any Use | No mask use | 35 |
| Peck | 0 | 13 | 0 | 28 | USA | SARS | Laboratory | Medical mask (N95) | Cohort | Yes | Consistent Use | Inconsistent use | 36 |
| Burke | 0 | 63 | 0 | 13 | USA | SARS-CoV-2 | Laboratory | Medical mask | Cohort | Yes | Any Use | No mask use | 37 |
| Barasheed | 4 | 36 | 2 | 53 | KSA | Respiratory Virus | Laboratory | Medical mask | cRCT | No | Medical mask | No mask use | 38 |
| Suess | 6 | 69 | 19 | 82 | Germany | Influenza | Laboratory | Medical mask | cRCT | No | Facemask | No mask use | 39 |
| Suess | 10 | 67 | 19 | 82 | Germany | Influenza | Laboratory | Medical mask | cRCT | No | Facemask + Hygiene | No mask use | 39 |
| Sung | 40 | 911 | 95 | 920 | USA | Respiratory Virus | Laboratory | Medical mask | Prospective Trial | Yes | Post-intervention mask period | Pre-intervention control period | 40 |
| Zhang | 1 | 16 | 8 | 28 | Airline Flight | H1N1 | Laboratory | Non-medical mask | Case Control | No | Any Use | No mask use | 41 |
| Zhang | 0 | 15 | 9 | 26 | Airline Flight | H1N1 | Laboratory | Non-medical mask | Case Control | No | Any Use | No mask use | 41 |
| Alraddadi | 11 | 151 | 7 | 66 | KSA | MERS | Laboratory | Medical mask | Cohort | Yes | Consistent Use | Inconsistent use | 42 |
| Jaeger | 0 | 20 | 9 | 43 | USA | H1N1 | Laboratory | Non-medical mask | Cohort | Yes | Any Use | No mask use | 43 |
| Jaeger | 0 | 12 | 9 | 51 | USA | H1N1 | Laboratory | Non-medical mask | Cohort | Yes | Consistent Use | Inconsistent use | 43 |
| Hall | 0 | 42 | 0 | 6 | KSA | MERS | Laboratory | Medical mask | Case Control | Yes | Any Use | No mask use | 44 |

Table S3. Results (cumulative deaths on January 1, 2021) from the two main scenarios presented in the main text and a third alternative scenario where mask use achieved 85% coverage within 7 days of the model projection

| **Region** | **Reference (95% UI)** | **Universal mask use (95% UI)** | **Alternative mask use (95% UI)** |
| --- | --- | --- | --- |
| Global | 3226003 (2183429–5221802) | 2085710 (1529000–3183895) | 2359967 (1697510–3673747) |
| Southeast Asia, East Asia, and Oceania | 125200 (46949–292524) | 87591 (30757–238976) | 102870 (38844–264347) |
| Central Europe, Eastern Europe, and Central Asia | 232097 (131801–436407) | 69468 (50952–92898) | 90172 (64013–133718) |
| High–income | 738007 (565234–1130053) | 519447 (461394–634322) | 536495 (473086–668933) |
| Latin America and Caribbean | 516094 (434664–614954) | 446680 (383343–518128) | 488043 (419130–571336) |
| North Africa and Middle East | 326138 (203558–552056) | 137336 (88545–235416) | 167892 (112826–286478) |
| South Asia | 1108806 (519804–2256104) | 751373 (345092–1638252) | 875027 (403383–1847628) |
| Sub–Saharan Africa | 179661 (75559–352596) | 73816 (33761–157277) | 99469 (42963–213231) |

#### Table S4. GATHER checklist

| Checklist # | Checklist item | Location |
| --- | --- | --- |
| Objectives and funding |  |  |
| 1 | Define the indicators, populations, and time periods for which estimates were made. | Main manuscript |
| 2 | List the funding sources for the work. | Main manuscript |
| Data Inputs |  |  |
| *For all data inputs from multiple sources that are synthesised as part of the study:* |  |  |
| 3 | Describe how the data were identified and how the data were accessed. | Supplementary appendix Sections 1.2, 2.1, 3.1; Supplementary Information previously published in Reiner et al.^1^ |
| 4 | Specify the inclusion and exclusion criteria. Identify all ad-hoc exclusions. | Supplementary appendix Sections 1.2, 2.1, 3.1; Supplementary Information previously published in Reiner et al.^1^ |
| 5 | Provide information on all included data sources and their main characteristics. For each data source used, report reference information or contact name/institution, population represented, data collection method, year(s) of data collection, sex and age range, diagnostic criteria or measurement method, and sample size, as relevant. | Supplementary Information previously published in Reiner et al.^1^ |
| 6 | Identify and describe any categories of input data that have potentially important biases (e.g., based on characteristics listed in item 5). | Supplementary Appendix Section 3.1 |
| *For data inputs that contribute to the analysis but were not synthesised as part of the study:* |  |  |
| 7 | Describe and give sources for any other data inputs. | Supplementary Information previously published in Reiner et al.^1^ |
| *For all data inputs:* |  |  |
| 8 | Provide all data inputs in a file format from which data can be efficiently extracted (e.g., a spreadsheet as opposed to a PDF), including all relevant meta-data listed in item 5. For any data inputs that cannot be shared due to ethical or legal reasons, such as third-party ownership, provide a contact name or the name of the institution that retains the right to the data. | See Supplementary Information previously published in Reiner et al.^1^ |
| Data analysis |  |  |
| 9 | Provide a conceptual overview of the data analysis method. A diagram may be helpful. | Main manuscript, Supplementary Appendix Sections 3–5 and Supplementary Information previously published in Reiner et al.^1^ |
| 10 | Provide a detailed description of all steps of the analysis, including mathematical formulae. This description should cover, as relevant, data cleaning, data pre-processing, data adjustments and weighting of data sources, and mathematical or statistical model(s). | Main manuscript, Supplementary Appendix Sections 3–5 and Supplementary Information previously published in Reiner et al.^1^ |
| 11 | Describe how candidate models were evaluated and how the final model(s) were selected. | Main manuscript, Supplementary Appendix Sections 3–5 and Supplementary Information previously published in Reiner et al.^1^ |
| 12 | Provide the results of an evaluation of model performance, if done, as well as the results of any relevant sensitivity analysis. | Main manuscript, Supplementary Appendix Sections 3–5 and Supplementary Information previously published in Reiner et al.^1^ |
| 13 | Describe methods for calculating uncertainty of the estimates. State which sources of uncertainty were, and were not, accounted for in the uncertainty analysis. | Main manuscript, Supplementary Appendix Sections 3–5 and Supplementary Information previously published in Reiner et al.^1^ |
| 14 | State how analytic or statistical source code used to generate estimates can be accessed. | Supplementary Information previously published in Reiner et al.^1^ |
| Results and Discussion |  |  |
| 15 | Provide published estimates in a file format from which data can be efficiently extracted. | Main manuscript Table 1 |
| 16 | Report a quantitative measure of the uncertainty of the estimates (e.g. uncertainty intervals). | Main manuscript  Online data visualization tool: <https://covid19.healthdata.org> |
| 17 | Interpret results in light of existing evidence. If updating a previous set of estimates, describe the reasons for changes in estimates. | Main manuscript |
| 18 | Discuss limitations of the estimates. Include a discussion of any modelling assumptions or data limitations that affect interpretation of the estimates. | Main manuscript, Supplementary Appendix Section 7 |
